# Specific methylation marks in promoter regions are associated to the pathogenic process of Chronic Chagas disease Cardiomyopathy by modifying transcription factor binding patterns

**DOI:** 10.1101/2021.12.20.21267972

**Authors:** Pauline Brochet, Barbara Ianni, Laurie Laugier, Amanda Farage Frade, João Paulo Silva Nunes, Priscila Camillo Teixeira, Charles Mady, Ludmila Rodrigues Pinto Ferreira, Quentin Ferré, Ronaldo Honorato Barros Santos, Andreia Kuramoto, Sandrine Cabantous, Samuel Steffen, Antonio Noedir Stolf, Pablo Pomerantzeff, Alfredo Inacio Fiorelli, Edimar A Bocchi, Cristina Wide Pissetti, Bruno Saba, Darlan da Silva Cândido, Fabrício Dias, Marcelo Sampaio, Fabio Antônio Gaiotto, José Antonio Marin-Neto, Abílio Fragata, Ricardo Costa Fernandes Zaniratto, Sergio Siqueira, Giselle de lima Peixoto, Vagner Oliveira-Carvalho Rigaud, Fernando Bacal, Paula Buck, Rafael Almeida Ribeiro, Hui Tzu Lin-Wang, José Antonio Marin-Neto, André Schmidt, Martino Martinelli, Mario Hiroyuki Hirata, Eduardo Donadi, Alexandre Costa Pereira, Virmondes Rodrigues, Denis Puthier, Jorge Kalil, Lionel Spinelli, Edecio Cunha-Neto, Christophe Chevillard

## Abstract

Chagas disease, caused by *Trypanosoma cruzi*, is an endemic parasitical disease of Latin America, affecting 7 million people. Although most patients are asymptomatic, 30% develop complications, including Chronic Chagasic Cardiomyopathy (CCC), which ranges from moderate to severe stages depending on the cardiac ejection fraction. The pathogenic process remains poorly understood, although genetic and epigenetic factors have already been proposed.

Based on bulk RNA-seq and EPIC methylation data, we investigated the genetic and epigenetic deregulations present in the moderate and severe stages of CCC. We identified 4 main biological processes associated with the pathology development, including immune response, ion transport, cardiac muscle processes and nervous system. An in-depth study of the transcription factors binding sites in the differentially methylated regions corroborated the importance of these processes. We also conducted a methylation study on blood to identify potential biomarkers for CCC. Our data revealed 198 differentially methylated positions (DMPs) that could serve as biomarkers of the disease, of which 61 are associated with disease severity.

## Introduction

Chagas disease is a neglected disease caused by the protozoan *Trypanosoma cruzi*. This parasite is endemic in 21 Latin America countries, where it affects around 7 million people by means of an insect vector, *Reduviidae*. With migratory flows, this disease can now be found in non-endemic countries and spread by congenital contamination or blood transfusion (1), notably in North America (2) (n>300,000), Europe (3) (n>100,000), Japan (4) (n>4,000) or Australia (5) (n>1,000). The clinical course of the disease comprises an acute and a chronic phase. For the majority of the patients, the acute stage is asymptomatic and lasts 4 to 8 weeks. After, the patients enter in the chronic phase, where 60% of the patients remain asymptomatic and 40% develop symptomatic disease, being 10% megaesophagous/megacolon, and 30% Chagas disease cardiomyopathy (CCC) with varying degrees of severity including refractory heart failure (1). This cardiomyopathy is the main cause of deaths from Chagas disease itself, and is one of the most lethal cardiomyopathies (6). Some drugs are effective on *T. cruzi* during the acute phase, but their effects during chronic phase are questionable and several side effects have been reported (7). The fact that the biological processes leading to CCC are not yet well understood has impaired the development of efficient therapeutical strategies.

The CCC myocardium displays a diffuse myocarditis with signs of inflammatory infiltrate and heart fiber damage, including significant fibrosis. The inflammatory infiltrate of CCC heart lesions is mainly composed of T cells displaying a Th1-like cytokine profile (8–11). This exacerbated Th1 response, characterized by a high secretion of interferon-gamma (IFN-*γ*) and tumor necrosis alpha (TNF-*α*), with lower production of interleukin (IL)-4, IL-6, IL-7, and IL-15, is associated to overexpression of Th1 transcription factors such as TBX21 (T-bet) (12). Interestingly, the intensity of the myocardial infiltrate was shown to be positively correlated with the abundance of CXCL9 mRNA (13). Moreover, our group has previously demonstrated that CCC myocardium presents a unique gene expression profile, distinct from the other dilated cardiomyopathies (12, 14).

Many studies have highlighted the importance of DNA methylation in the regulation of gene expression in dilated cardiomyopathy (15), in particular by the methylation/demethylation of transcription factor binding site (TFBS) located in genes regulatory regions (16). Development of severe CCC is also dependent of epigenetic regulations such as DNA methylation (17), but also involving miRNAs (14, 18) or lncRNAs (19) or DNA methylation processes. Considering these informations, the microarray analyses performed so far are not deep enough to analyze all the dysregulation of non-coding RNAs. Moreover, the Illumina Infinium HumanMethylation450 BeadChip, used to perform most of the previous methylation analyses, has been discontinued and was known to miss important regulatory regions (20). In this study, to strengthen our analysis and get a more complete picture of the epigenomic landscape of CCC myocardium, we performed gene expression analysis using RNA-seq complemented with Illumina Infinium MethylationEPIC array BeadChip, covering 96% of gene loci, including lncRNA. Moreover, blood samples from moderate and severe CCC were also evaluated to further decipher the biological process associated with Chagas dilated cardiomyopathy and to identify possible blood biomarkers useful for diagnosis.

## Methods

### Ethical Considerations

The protocol was approved by the institutional review boards of the University of São Paulo School of Medicine and INSERM (French National Institute of Health and Medical Research). Written informed consent was obtained from all patients or family members. All experimental methods comply with the Helsinki Declaration.

### Patients and Myocardial Tissue Collection

Human left ventricular free wall heart tissue samples were obtained from patients with end-stage heart failure CCC at the time of heart transplantation (n=8). CCC patients underwent a serological diagnosis of *T. cruzi* infection and standard electrocardiography and echocardiography, and tissues were subject to histopathological assessment as previously described (21). Biopsies from controls (n=6) were obtained from healthy hearts of organ donors having no suitable recipient, and biopsies for dilated cardiomyopathy (DCM) from end-stage patients, at the time of heart transplantation (n=8).

### RNA Extraction and sequencing

Heart tissue samples (20–30 mg) were cleared from pericardial fat, crushed with ceramic beads (CK14, diameter 1.4 mm, Bertin) in 350 µL of RLT lysis buffer supplemented with 3.5 µL of β-mercapto-ethanol. Total RNA was extracted from biopsies using the RNeasy Mini Kit adapted with Trizol. RNA quantity and quality were measured with a 2100 BioAnalyzer (Agilent). Ribosomal RNAs were depleted and samples were prepared for sequencing according to the Illumina TruSeq RNA Preparation Kit and subjected to pairwise sequencing (2×150bp) with an Illumina HiSeq sequencer. Sequencer results were obtained as raw fastq file format.

### Quality control and alignment

Raw data quality was verified with *FastQC* (v0.11.5). Low-quality reads, Illumina adapters and reads smaller than 20 nucleotides were removed with *Trimmomatic* (v0.39) (22), using default values for other options. Reads were aligned on GRCh37 (hg19) human reference genome from Ensembl using STAR (v2.5.4b) (23), specifying that the reads are 2×150 nucleotides (paired-end). Alignment quality was checked by *BAMQC* (*qualimap* v2.2.1) (24). Gene quantification was done with *featureCounts* (v2.0.0) (25), using the exons as features and the genes as attributes. All the bioinformatics steps are arranged in a Snakemake (v5.7.4) (26) workflow and full reproductibility is ensure though a Docker image, available at this address (github link).

### Differential expression analysis

Statistical analyses were performed using R version 3.6.2. Because there is a large infiltrate of immune cells in the heart tissue, and therefore in the samples studies, genes related to immunoglobulins and T-cell receptor (TCR) were excluded of the analyses. The *DESeq2* package (v1.26.0) was used for data normalization and differential gene expression analyses (27). Log2 fold change (log2(FC)) were corrected using the DESeq2 shrinkage function. Multi testing correction was performed using the Benjamini-Hochberg method to obtain False Discovery Rate (FDR) for each gene. Genes with an FDR value lower than 0.05 and an absolute log2(FC) greater than 1.5 were considered as differentially expressed (DEG). Subsequent diagrams were produced with *Enhanced Volcano* (v1.4.0) (https://github.com/kevinblighe/EnhancedVolcano) or *ggplot2* packages (28). Moreover, hierarchical clustering (HCA), using Spearman correlation distance as distance metric, was computed using *gplots* (v3.0.4) package (https://rdrr.io/cran/gplots/). Principal component analysis (PCA) were computed with *factoextra* (v1.0.7) and *FactoMineR* (v2.3) packages (http://lib.stat.cmu.edu/R/CRAN/).

### Functional enrichment

Functional enrichment was computed using two strategies: *(i)* using *ClueGO* cytoscape plugin for Gene Ontology Biological Process annotations (GO BP) (29) and using *GAGE* package (v2.36.0) and *pathview* package (v1.26.0) for KEGG pathways annotations (30, 31). The interaction between differentially expressed ncRNA and protein-coding genes was analyzed with LncRNA2Target and LncTarD databases (32, 33). Considering the lack of annotation of lncRNA function, the LncRNADisease database was also used (34).

### Evaluation of cell types in heart tissue

RNAseq deconvolution was performed using ADAPTS R package (35). Cell type signatures coming from microarray and single-cell RNA-seq datasets were used: a first one containing 22 cell types of immune cells from PBMC samples (36), and a second one containing 5 cell types from the left ventricle of healthy heart tissue (37). Signature matrices were generated with ADAPTS, and then the proportion of each cell type was estimated on our total normalized RNA-seq data (with the Ig and TR genes). Finally, for each cell type, a Wilcoxon test (FDR ≤ 0.05) was applied to identify which ones have significantly different amount between cases and controls. To confirm these results, we performed the same analysis using Cybersortx Fractions and we obtained the same qualitative results (data not shown).

### Tissue DNA methylation analysis

Analysis of DNA methylation data was performed with the *ChAMP* package (38). The beta values were normalized with the BMIQ method (39), and the batch effect was corrected with the *ComBat* method (40). The analysis of variation of methylation on genomic positions was done using the *ChAMP* package and multitesting correction was performed using the Benjamini-Hochberg method (FDR, False Discovery Rate). Only genomic positions with an FDR ≤ 0.05 and a |Δβ| ≥ 0.2 (41) were selected. A DMP (Differentially Methylated Position) is associated with a gene when the DMP is inside the gene body or in its promoter region (from gene TSS to 1.5kb upstream), according to Illumina annotations. The functional analysis was done in the same manner as for DEGs (see «Functional enrichment» paragraph).

### Transcription factor characterization

An analysis of differentially methylated regions (DMR) was done with the ChAMP package, using the DMRCate method, with parameters lambda=400 and C=2 (42). A DMR of interest was defined as a region containing at least 1 DMP located in TSS ([TSS + 1500bp; TSS]), 1^st^ Exon or 5’UTR region of DEGs; and having a FDR less than or equal to 0.05. In order to identify transcription factor binding sites (TFBS) affected by a difference in methylation, the ReMap database was used (43). A total of 84 cell lines was selected (**Supplementary Table 1**), containing immune and heart-related cells, and including 151 transcription factors (TF). First, the transcription factors specifically associated with DMRs were identified with the OLOGRAM tool (44) in a pairwise analysis, meaning we identify the individual TFs enriched with the DMRs. Only those with an FDR ≤ 0.05 were retained. In a second step, we studied the combinations of those selected TFs that were observed in the DMR by using the n-wise overlap option of OLOGRAM (option –more-bed-multiple-overlap). In this latter analysis, we studied combinations of up to 4. The idea of the combinations of TFs was that if we have a complex of A+B+C, it means that A, B and C co-occur on the genome and may thus have functional relationship.

### Blood DNA collection

Blood (5 to 15 ml of blood) from 96 CCC patients (48 moderate CCC (Left ventricular ejection fraction > 40%) and 48 severe CCC (left ventricular ejection fraction < 40%)) and 48 asymptomatic Chagas disease controls was also collected in EDTA tubes. Genomic DNA was isolated on a silica-membrane according to the manufacturer’s protocol (QIAamp DNA Blood Max Kit, Qiagen, Hilden, Germany).

### Blood DNA methylation analysis

In order to ensure that no associations were missed in this exploratory analysis, no fold change threshold was chosen. A DMP was only defined by a FDR ≤ 0.05. Two different analyses were done: differentially methylated sites between controls and severe CCC; and differentially methylated sites between moderate and severe CCCs. For both analyses, a functional enrichment analysis was made on associated genes as previously (see «Functional enrichment» paragraph). In order to confirm the relevance of our analysis on blood methylation in moderate patients, the biological processes affected by the differences in methylation between asymptomatic and severe CCC on blood samples were compared to those found between controls and severe CCC for tissue RNAseq and methylation.

### Blood biomarkers search

For this analysis, the cohorts were mixed and divided in two groups: a training group, containing 70% of the data and a validation group containing the remaining samples. Two cases were studied: the differentially methylated sites between controls and all the CCC (indicator of disease presence), and the differentially methylated sites between moderate and severe CCCs (indicator of disease severity). After DMP selection, 3 different machine learning (ML) models were tested with our dataset: a decision tree, a logistic regression, and a random forest. On one hand, for the tree-based models, the following options were fitted: number of estimators, maximal depth, maximal features and minimal samples per leaf. On the other hand, the penalty and the parameter of regularization C (a method which reduce the risk of overfitting by favoring the simplest models) were optimized in logistic regression.

For each case, the analysis was made in two steps: a low number of DMPs were selected according to the statistical test included in ChAMP package, and then different models of ML were applied on this data. The models were adjusted depending on the parameters mentioned above (number of DMP, number of estimators, maximal depth…), and the set of model-parameters providing the better accuracy (TP (True Positive) + TN (True Negative) / All) was kept. If the accuracy was still low, the analysis was restarted and relaunched from the beginning, with a bigger number of DMPs (classified by FDR). All the ML analyses were done on Python3 using the python pandas (https://pandas.pydata.org/) and scikit-learn packages (http://citebay.com/how-to-cite/scikit-learn/). Cross validations were performed using the training group only. Generalization tests were done using the validation group.

## Results

### Understanding biological processes associated with CCC

#### Description of datasets

Gene expression analysis was conducted on left ventricular free wall myocardial tissue from 8 severe CCC patients and 6 healthy organ donors (see workflow: **Figure 1**). Gene expression analysis was done by sequencing total RNA (paired end 2×150bp) after ribosomal RNAs depletion. For each sample, we obtained between 40 and 75 million sequencing reads. Reads were aligned to the human reference genome GRCh37/hg19 using STAR. The average mappable rate of the raw reads reached 90% (± 2%). Tissue sample DNA methylation analysis was conducted on the same samples as gene expression. This analysis was done using Illumina EPIC kit, allowing the study of 850.000 methylation sites. After quality control, 721.803 CpGs were retained for analysis. For blood DNA methylation analysis, the same tool as for tissue DNA methylation was used. After quality control, 138 samples were kept (48 asymptomatic, 47 moderate CCC and 43 severe CCC), and 736.662 CpGs were retained for analysis.

**Figure 1.**
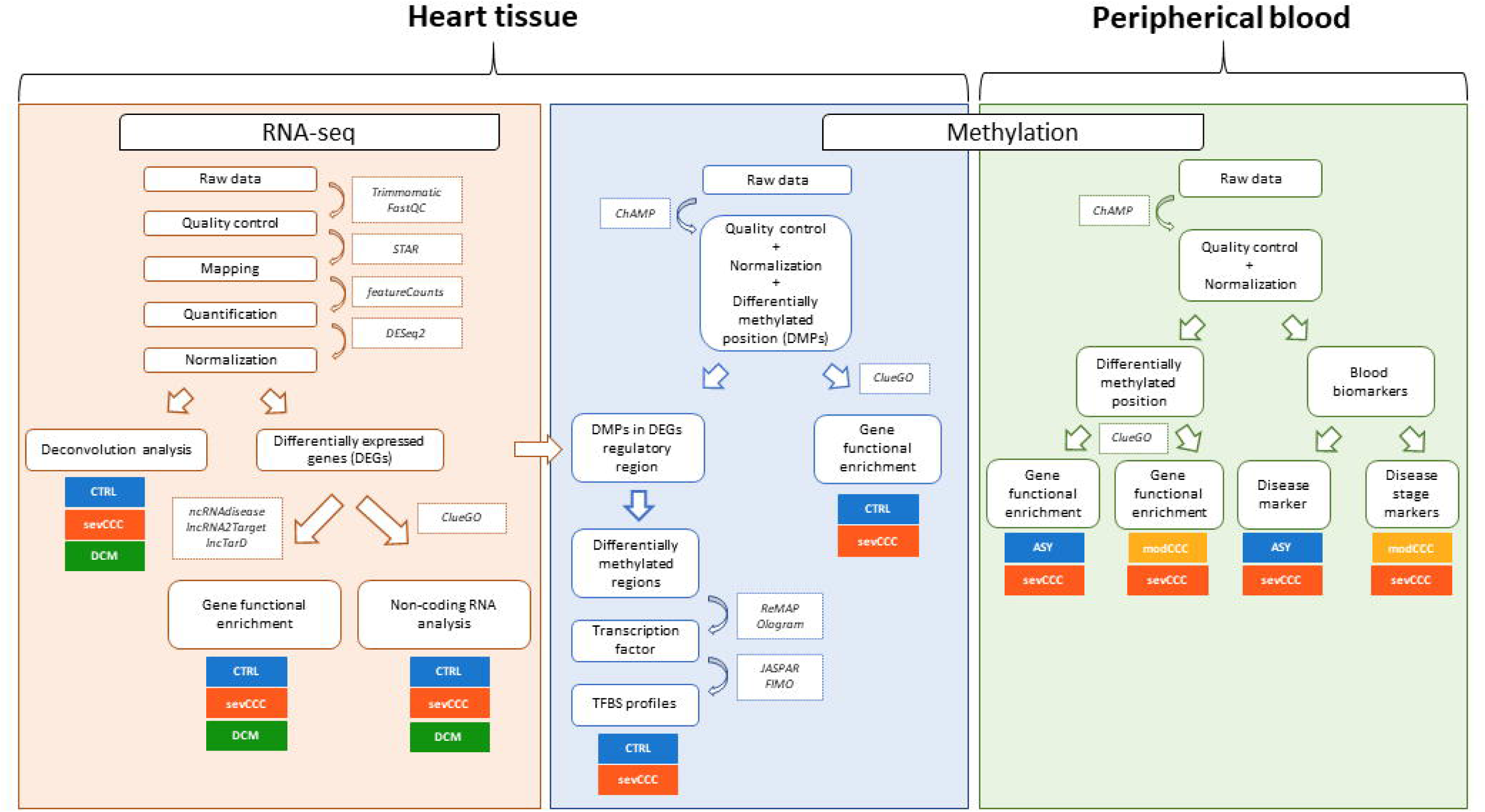
Workflow overview. Heart tissue RNAseq (orange) quality was verified with *fastQC*. Low-quality base and adapters were removed using *Trimmomatic*, and remaining reads were aligned with *STAR* aligner against hg19 human genome. Normalization and differential expression test were performed with *DESeq2*. *ClueGO* and ncRNAseq databases (*ncRNAdisease*, *lncRNA2target*, *lncTarD*) were used to characterized genes functionality. On the other hand, a deconvolution analysis was performed with *ADAPTS* on control and severe CCC samples. Heart tissue (blue) and blood (green) methylation analysis followed the same first steps: quality control, normalization, batch effect correction and differential methylation position (DMP) test were all performed with *ChAMP*. Genes associated with DMP (DMP in promoter region, body or 3’UTR) were functionally annotated with *ClueGO*. In tissue samples, differentially methylated regions located in differentially expressed genes regulatory region (from TSS-1500 to first exon) were searched with *ChAMP*. Transcription factor binding site (TFBS) were predicted with *OLOGRAM* using *ReMap* database, and TFBS profiles were identified with *FIMO*, using *JASPAR* database. In blood samples, two analyses were performed: DMP between asymptomatic and severe CCC, and DMP between moderate and severe CCC. In both cases, gene enrichment was performed in the same way than tissue methylation data. Finally, in blood samples, blood biomarkers were identified, for disease diagnosis and disease stage diagnosis.

### Gene expression profile

To understand the pathogenic processes driving CCC, we have studied the genomic deregulation of patients at several scales. Gene expression data were obtained from 43533 transcripts. These include protein coding units (43.03%); pseudogenes (20.71%); long non-coding RNA (lincRNA) (11.54%); antisense sequences (10.12%); micro RNA (miRNA) (3.64%); miscellaneous RNA (miscRNA) (3.03%); small nuclear RNA (snRNA) (2.45%); small nucleolar RNA (snoRNA) (1.99%). sense intronic sequences (1.54%); among others (**Supplementary Table 2**). A small fraction of these transcripts (1407/43533 (3.23%)) were considered as differentially expressed (FDR ≤ 0.05 and absolute log2FoldChange ≥ 1.5) between the two groups of samples (**Supplementary Table 3**). Most of these differentially expressed transcripts were overexpressed in CCC (1176/1407 (83.5%)). Among the 1407 transcripts, we detected protein coding units (63.54%); and non-coding RNAs (36.46%) pseudogene (11.02%); lincRNA (10.31%); antisense sequences (9.38%); miRNA (1.35%); misc RNA (0.57%); snRNA (0.64%); snoRNA (0.28%); sense intronic sequences (1.00%)). A specific enrichment occurs in protein coding (Chi squared corrected p-value (FDR) = 3.36E-20, OR = 90) and non-coding genes, as miRNAs (Chi square corrected p-value (FDR) = 1.96E-05, OR = 21) or pseudogenes (Chi square corrected p-value (FDR) = 4.96E-14, OR = 61) (**Supplementary Table 2**).

A PCA analysis was performed with all differentially expressed genes (DEGs) (**Supplementary figure 1A**). The first component was sufficient to explain 62.9% of the whole variance. This main component separates the samples between CCC and control, confirming that CCC myocardial gene expression patterns were substantially different from controls. A Hierarchical Clustering Analysis (HCA) computed with the normalized count matrix confirmed the importance of those features in the disease (**Figure 2A**). The sex and the age of the patients have no impact on this clustering (**Supplementary Figure 2A-2B**).

**Figure 2.**
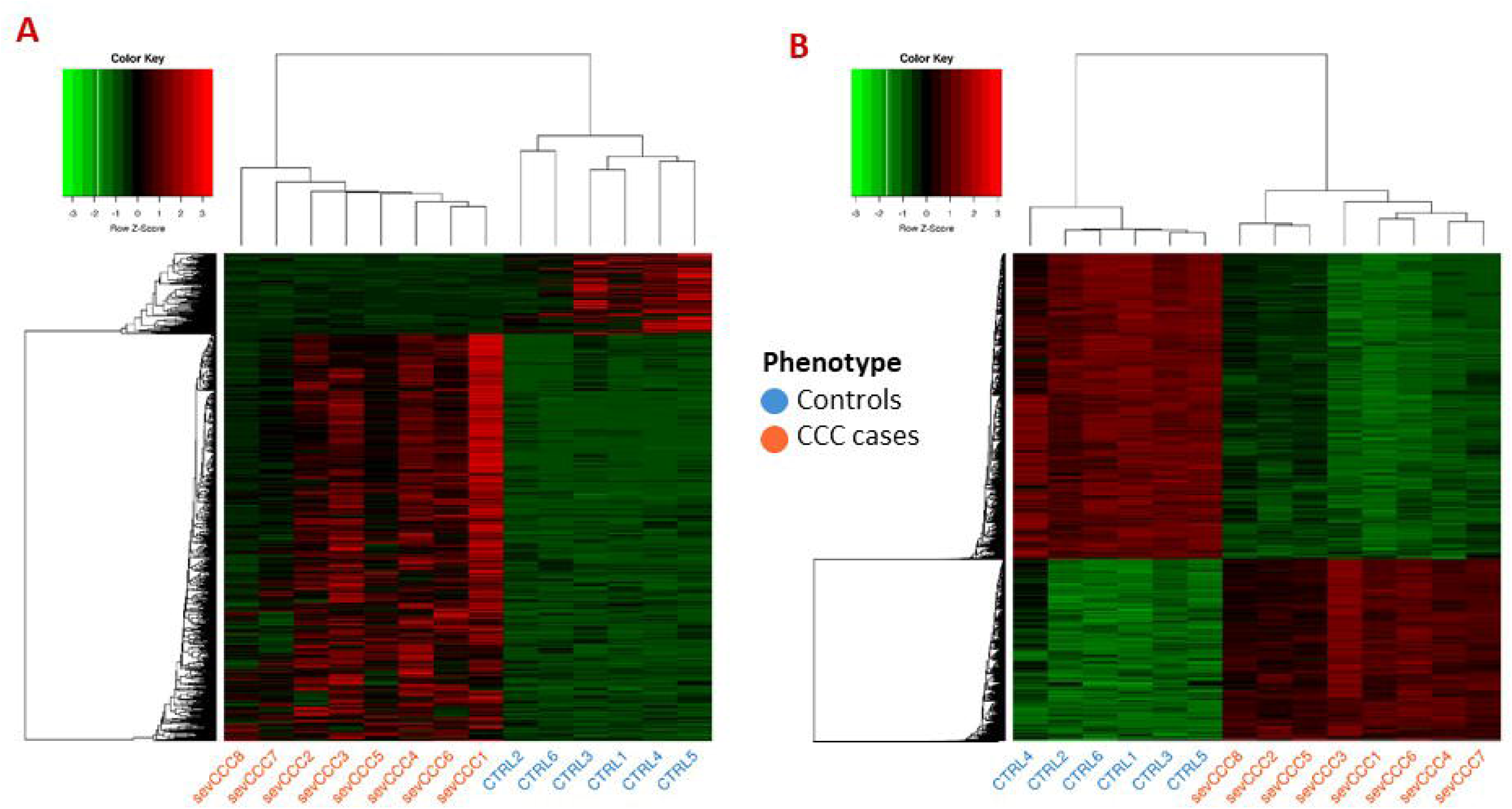
Analysis of samples clustering based on differentially expressed genes or differentially methylated CpG sites. Control samples identifiers are written in blue whereas case samples identifiers are written in red. **(A)** Hierarchical Clustering Analysis (HCA) performed on 6 control and 8 case samples, based on expression of 1409 differentially expressed genes. **(B)** Hierarchical Clustering Analysis (HCA) performed on the same samples as in A), based on methylation level of 16883 differentially methylated position.

### Severe CCC is characterized by a strong, specific immune response

In order to determine the biological processes specifically affected by CCCs, a comparative study was conducted on DCM patients. When we compared the gene expression pattern between DCM and controls, 3188 genes (707 up, 2483 down) are differentially expressed in DCMs. Among them, of which only 290 (9%) DEGs are in common with CCCs (120 up, 162 down) (**Supplementary Table 4**). It suggests that the mechanism involved in CCC and DCM are not similar.

A functional analysis of the DEGs identified either in CCCs or DCMs was conducted (**Supplementary Figure 3**). For each GO ontology, the percentage of DEGs and their relative contribution to each phenotype were measured. As example, the “T cell selection” GO ontology entry is including 53 genes. 24 (45.3%) genes were DEGs in CCC or DCM or both. Among them, 21/24 (87.5%) of these DEGs are CCC DEGs and 4/24 (16.6%) of these DEGs are DCM DEGs. So, this process seems to be more CCC specific. Thus, some biological functions (GO ontologies) could be predominantly attributed to either of CCC or DCM (**Supplementary Table 5**).

Interestingly, calcium ions are particularly affected in CCC, not in DCM (calcium ion transmembrane import into cytosol, regulation of sequestering of calcium ion). With regard to CCCs, DEGs are almost exclusively specific to the immune response (innate or adaptive), characterized by immune receptors (adaptive immune response based on somatic recombination of immune receptors built from immunoglobulin superfamily domains, T cell receptor signaling pathway, immunological synapse formation). Most of enriched terms are related to T lymphocytes (T cell proliferation, positive regulation of T cell differentiation). More specifically, the T CD8+ (CD8-positive, alpha-beta T cell activation) and T CD4+ (CD4-positive, alpha-beta T cell differentiation) are associated to CCC. A strong enrichment of the Th1 response is associated to CCC (T-helper 1 type immune response, positive regulation of interferon-gamma production, response to interferon-gamma). Besides T cells, other immune cells seem to act in the pathogenic process of CCC, such as B cells (B cell differentiation, B cell proliferation), macrophages (macrophage migration) or NK cells (natural killer cell mediated cytotoxicity). The regulation of interleukin production is also affected, included IL-1, IL-4, IL-6, IL-10 and IL-12.

The DEGs identified in DCMs are involved in processes related to muscle and blood systems (animal organ development, blood vessel development, muscle cell proliferation, circulatory system process). Ion transport is also strongly represented (cation transport, ion transport, gated channel activity). Besides that, nervous system is also affected in DCM (nervous system development, neurogenesis), as well as cell damaging process (wound healing, regulation of cell death).

These diseases have similar pathogenic processes, including those related to ion transport (ion homeostasis, cellular cation homeostasis, regulation of metal ion transport) and smooth muscle (positive regulation of smooth muscle cell proliferation). The ERK1/ERK2 cascade (positive regulation of ERK1 and ERK2 cascade) also appears to be affected in both cases. Together, these results provide an overview of the pathogenic process associated with CCCs, which is mostly related to the immune response. Although it seems normal not to detect an immune response in non-inflammatory DCMs, it is important to note that in severe CCCs the pathogen is absent. Furthermore, calcium ion transport seems to be particularly important in CCCs, compared to DCMs.

### Several Non-coding RNA are differentially expressed

Non-coding RNAs are among the elements that can lead to genetic dysregulation. As gene expression analysis was performed by sequencing of total RNAs, we got information on non-coding RNAs (lincRNA, miRNA, snRNA, snoRNA, 3’overlapping ncRNA). Up to 3777 non-coding elements were detected in our 14 samples by total RNA sequencing. Among them, 179 ncRNAs, including 19 miRNAs and 145 lncRNAs; were differentially expressed (**Supplementary Table 6**). Those ncRNAs (**Supplementary Figure 4A**) were enough to classify samples according to their phenotype, demonstrating the importance of these non-coding elements in chronic Chagas cardiomyopathy. In this short list, the Myocardial Infarction Associated Transcript (MIAT) that has been also previously associated with an increased susceptibility to Chagas disease is present. Among the 19 miRNAs, 12 were annotated in miRNA databases and 7 of them were just tagged as miRNAs. Among the differentially expressed lncRNAs, 6 were annotated as targeting a gene (**lncRNA - targeted gene:** MIAT - miR-133a, RP11 - 276H19.1-GAS1, XIST - WNT1, MIR155HG - miR-155, LINC00707 - ELAVL1 and KB-1732A1.1 - E2F1).

A similar analysis was performed on the ncRNAs differentially expressed between the 6 controls and 8 samples affected by non-inflammatory dilated cardiomyopathy (DCM) (**Supplementary Table 6**). 327 ncRNAs, including 225 lncRNAs and 54 miRNAs, are associated to DCM (**Supplementary Figure 4B**). 143/179 ncRNAs are specific to CCC (**Supplementary Figure 4C**). These ncRNAs are mostly non annotated lncRNAs but including KB-1732A1.1 and RP11-276H19.1.

### Tissue DNA methylation profile

Given the importance of methylation in the regulation of genetic response (45), we also performed several DNA methylation analyses in controls and CCC patients. Tissue DNA methylation analysis, performed on the same samples as RNA-seq analysis, was carried out on a total of 721.802 CpGs, after quality control. Those CpGs are located in gene body (37.46%), but also in intergenic regions (27.55%), TSS ([TSS - 1500bp; TSS]) (20.11%), 5’UTR (8.66%), 1^st^ Exon (3.03%), 3’UTR (2.5%) and exon boundary (0.69%) (**Supplementary Table 7**). Only 16883 of those CpGs (2.34%) were differentially methylated (FDR ≤ 0.05 and |Δβ| ≥ 0.2) and the majority being hypo-methylated (n=10097, 59.81%) (**Supplementary Table 8**). A PCA conducted on the normalized methylation level of these 16883 Differentially Methylated Position (DMP) shows that a single principal component was enough to explain 89.6% of the full dataset variance (**Supplementary Figure 1B**), and to separate the CCC and control samples. A heatmap of the same DMP methylation levels with HCA (**Figure 2B**) confirmed the clustering of the samples according to their phenotypes. The sex and the age of the patients seems to have no impact on this clustering (**Supplementary Figure 2C-2D**). A total of 5814 genes are associated (i.e. DMP located in gene body, in 3’UTR or in gene upstream regions, from TSS to 1500bp) to all 16883 DMPs. The localization of these DMP is significantly different as compared to the localization of the whole tested CpGs (Chi square corrected p-value (FDR) < 2.2E-16).

### Several DEGs identified on tissue samples are also affected by DNA methylation alterations

On tissue samples, 16883 differentially methylated CpG sites were detected. These DMPs are located in or around 5814 genes. Among them, 996 DMPs are located in or around 390 DEGs. The localization of these 996 DMPs is : 5’UTR = 195; TSS1500 = 155; TSS200 = 91; 1^st^ Exon = 68; Body = 460; Exonic Boundary =10; 3’UTR = 17. Although none of these regions are significantly enriched (in total number of DMPs), the upstream regions of the gene are enriched in down-methylated DMPs : the 1500 bp before TSS (86.58%, padj=1.94E-45), the 1^st^ exon (88.23%, padj=4.22E-21) and the 5’UTR (88.72%, padj=2.56E-08). Moreover, these down-methylated DMPs are, in 86% of the cases, associated with over-expressed DEGs, the other cases being much less frequent (down-methylated DMPs - down-expressed DEG: 2%; up-methylated DMPs - up-expressed DEG: 9%; up-methylated DMPs - down-expressed DEG: 3%). Knowing the impact of DNA methylation on gene regulation, and in particular the association between promoter undermethylation and gene overexpression (46), we retained these regions for further analysis. In this article, we will call “regulatory region” the region between the TSS and 1500bp among the TSS as well as 5’UTR and gene 1^st^ exon.

### Identification of transcription factor potentially involved in severe CCC

Based on the 409 DMPs associated with DEG regulatory regions, we were able to define 92 regulatory DMRs (**Supplementary table 9)**. These DMRs span on average 245bp, ranging from 3 to 10121bp (Q1 = 90.25, median = 245.10, Q3 = 340), and are in the promoter regions of 89 DEGs.

In order to identify the transcription factors whose binding is specifically affected by the methylation/demethylation of these DMRs, three analyses were performed with OLOGRAM. The principle is to take a “query” set of sequences of interest (here, the DMRs), and to count their overlaps (in base pairs) with another “reference” set of sequences (here, each TFBS). The tool determines whether the query overlaps the reference more than what would be expected by chance, assuming that both query and reference can only be found in the inclusion region. This means that if query and reference overlap is only due to the fact that both are present in the promoter regions, their enrichment would be zero. In order to assess which TFs preferentially bind to DMRs, and not to the entire promoter regions of DEGs, two analyses were performed: (A) with the promoters of the DEGs as query and all genomic promoters as inclusion regions and (B) with the DMRs as query and all genomic promoters as inclusion regions (**Figure 3A**). The log2(FC)s of each TF in analysis (A) and (B) are significantly correlated (spearman p-value ≤ 0.05), but the r2 is low (0.49) (**Figure 3B**) suggesting a different signal carried by the DMRs compared to all the promoters including the DMRs. Moreover, the log2(FC)s obtained with the DMRs (analysis B) are significantly higher than the ones obtained with the promoters containing DMRs (analysis A) (Wilcoxon p-value = 4.23e-07) (**Figure 3C**). These two analyses clearly show that there is a stronger enrichment of TFs in DMRs than in the promoter set. To confirm these results, a third analysis (C) was performed with the DMRs as query and the promoters of the DEGs as inclusion regions. The obtained distribution of the log2(FC) with this analysis showed two distinct peaks, one around 0, and another around 1.7 (**Figure 3C**), confirming that some TFs are specifically enriched in DMRs compared to the promoter as a whole. A total of 30 TFs were found significantly associated to the DMRs with this analysis, and those 30 TFs were also found associated to the DMRs in analysis B (**Supplementary table 10)**.

**Figure 3.**
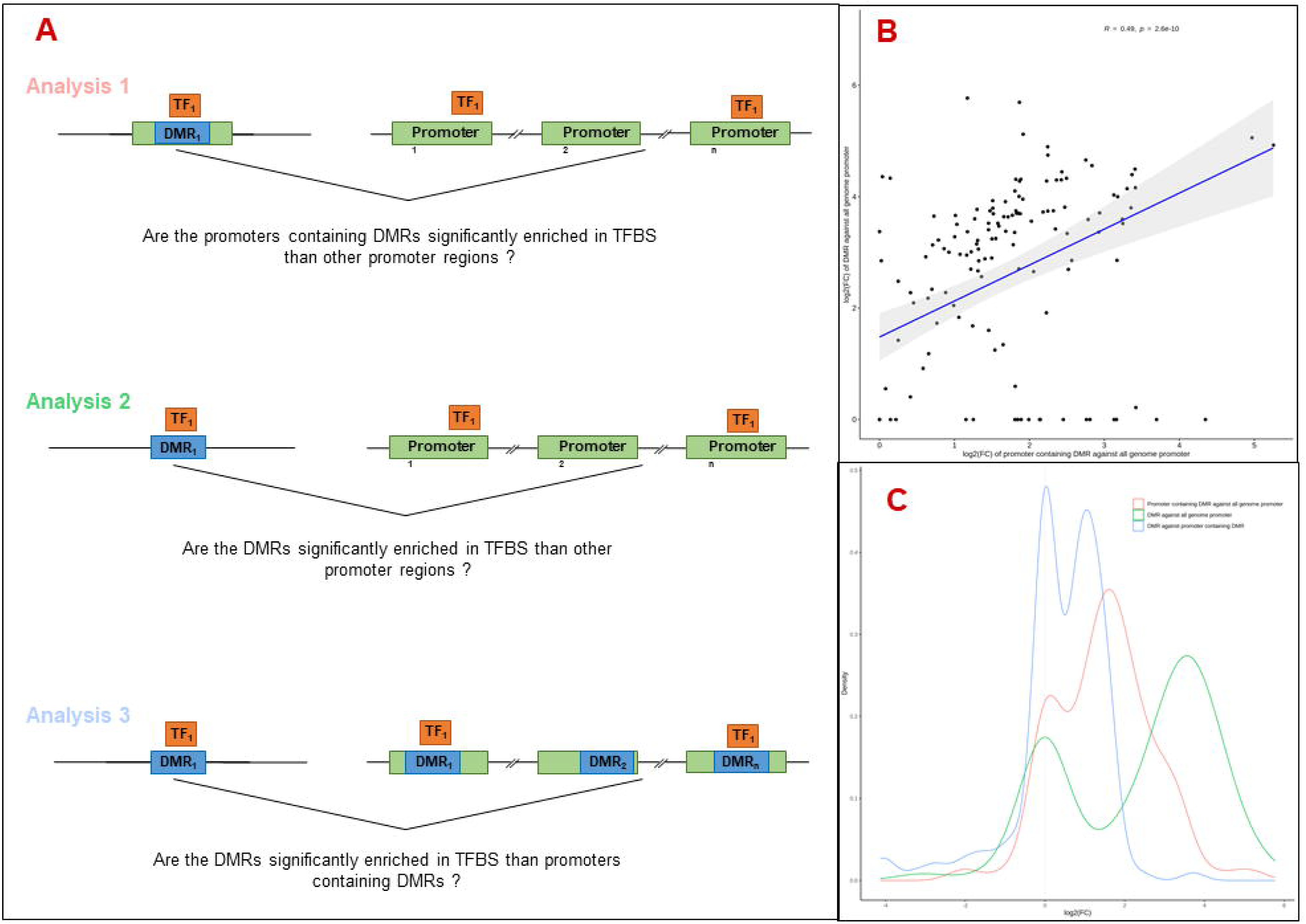
Analysis of the relation between TFBS (Transcription Factor Binding Site) **(A)** Schematic illustration of the three approaches used in this analysis. Differentially methylated region (DMR) is highlighted in blue, gene regulatory region in green, and transcription factor (TF) in orange. Analysis 1: TFBS enrichment in regulatory region containing at least one DMR, compared to all genome regulatory region. For each gene, a regulatory region is defined as the region from TSS-1500 to first exon. Analysis 2: TFBS enrichment in DMR compared to all genome regulatory region. Analysis 3: TFBS enrichment in DMR compared to regulatory region containing at least one DMR. **(B)** Scatter plot of the log2(FC) obtained with the analysis 1 and 2 and Spearman correlation of these values. The fold change is computed according to the observed S value compared to obtained S value, S corresponding to the number of overlapping bases between TFBS and query region. **(C)** Distribution of the log2(FC) obtained with the 3 approaches.

### Prediction of TF complex

Considering the fact that transcription factors often act together, forming complexes, an analysis of interaction between those transcription factors was performed using the OLOGRAM tool (44). Here, we considered as a Cis-Regulatory Module (CRMs) the regions where at least 2 TFs bind to the genome according to ReMap. After data filtering and considering combinations of up to 4 TFs, we have identified 16 regions significantly associated with our DMRs, involving a total of 12 transcription factors (**Supplementary table 10)**. Among the top-regulators, BRD4 which appear in 10 (83.33%) of the identified complex and overlap a total of 3090bp (number S). Comes after EED (S = 2777), EBF1 (S = 2389), BCLAF1 (S = 2312), TBX21 (S = 2271), RUNX3 (S = 2092) and RUNX1 (S = 2052). All these transcription factor complexes contain EED or BRD4. Interestingly, some complexes seemed to act on specific DEGs, such as BCL6 + BRD4 + GATA3.

### Biological process affected by TFBS methylation

The 30 TF previously identified are involved in several biological process, such as somatic recombination of immunoglobulin gene segments (TCF3, YY1, BCL6, TBX21), regulation of cardiac muscle tissue growth (NOTCH1, RBPJ, RUNX1, YY1) or peripheral nervous system neuron development (RUNX1, RUNX3) (**Supplementary table 11**). However, the main signal here is regulation of T cell differentiation (BCL6, CBFB, GATA3, IRF4, MYB, RUNX1, RUNX3, STAT5B, TBX21, ZEB1).

Subsequently, two biological aspects were discussed. A first analysis focused on the TFs involved in heart-relative or neurological process (**Supplementary Figure 5**) (n = 7: MYC, YY1, RBPJ, GATA3, RUNX1, RUNX3). Among these TFs, only RUNX3 was differentially expressed in our data. Interestingly, this up-regulated TF seemed to autoregulate itself (**Supplementary table 12**). The targeted genes (which are all up-regulated) are essentially involved in immune related processes, according to GO analysis, notably in lymphocyte pathways (regulation of T cell receptor signaling pathway, regulation of B cell proliferation, regulation of T cell activation…). However, according to our custom list of pathways of interest, some of those genes are also involved in fibrosis (SELPLG, CCR5, CD74, PRF1, KCNN4, IFN-γ), ion channel (KCNN4) or mitochondria (CASP8). These genes are targeted by at least one of these TF: RUNX1, MYC and RUNX3.

A second analysis was focused on the genes involved in the Th1/IFN-γ response according to our lists of interest. These 19 genes are potentially targeted by 28 of the 30 TF previously identified, again illustrating the importance of the immune response in the pathogenic process associated with CCC. In order to identify the TFs most involved in the Th1 response, only those targeting at least 11 of the 19 genes were selected: BCLAF1, BRD4, CBFB, EED, PAX5, RUNX3 and TBX21 (**Figure 4**). Although most of the selected DEGs are targeted by all TFs, a few appear to be targeted by specific TFs: IFN-γ by RUNX3+TBX21, ITK and SOAT2 by BRD4, CTLA4 by PAX5+BRD4, and S1PR4 by BRD4+CBFB+BCLAF1.

**Figure 4.**
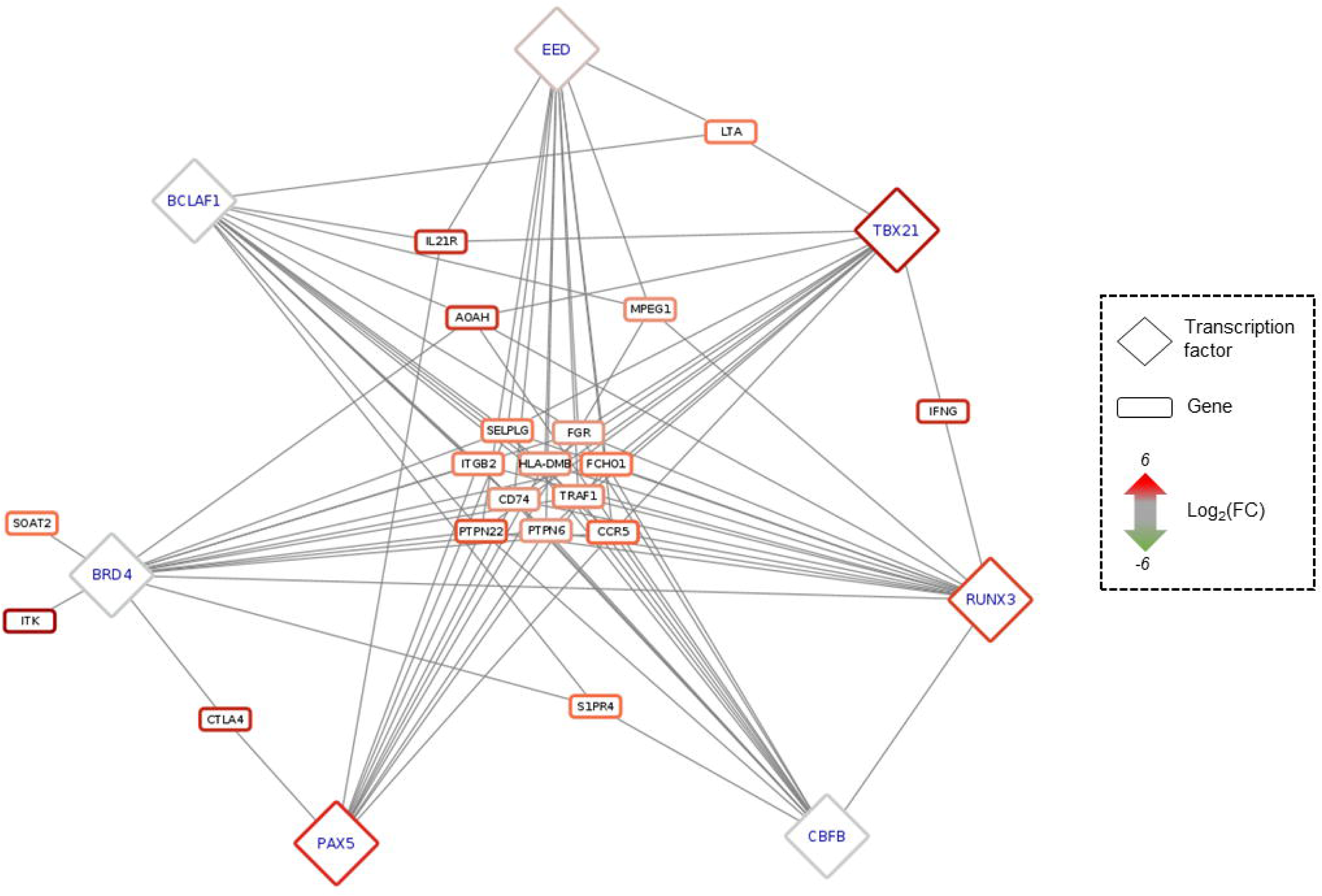
Predicted regulatory interaction in IFNy-Th1 pathway. Network composed by 19 genes involved in IFNy-Th1 pathway, and the top 7 TF predicted as targeting those 19 genes, according to *OLOGRAM* based on *ReMap* database. TF are written in blue in diamond, and genes in black in rectangle. Shapes borders are colored according to the fold change, from green to red.

### TFBS pattern affected by DNA methylation

To confirm that DNA methylation is indeed what affects the binding of the identified TFs, an analysis was conducted on their binding motif. Of the 30 TFs of interest, 20 have at least one known motif in the Jaspar database, providing a total of 45 distinct motif. 127,158 genomic sequences included in 92 of our DMR were significantly associated with at least one motif, 35,091 are present in the DMRs predicted with OLOGRAM, 22,325 have at least one CG and 939 have at least one DMP. After removing redundancy in TFBS, 423 TFBS-DMR specific sequence pairs have been identified. None of the TFs of interest were discarded by the set of filters used, all 20 having TFBSs in the DMRs. These sequences are located in the promoter regions of 48 DEGs. Among the TFs mainly involved in complexes (BRD4, EED, EBF1, BCLAF1, TBX21, RUNX3, RUNX1), TFBS pattern is known for EBF1, TBX21, RUNX3 and RUNX1. Interestingly, TBX21, RUNX3 and EBF1 are the TFs whose binding motif appears to be affected by DNA methylation in the largest number of genes (n = 25, 18 and 18 respectively). Because these TFs are involved in a large number of complexes, regulation of their binding may affect the binding of all TFs in the complexes.

Among the 25 genes with a low number of TFs binding in their promoter region (n <= 3), and thus being affected by specific TFs, 7 are targeted by TBX21 and 4 by RUNX3, showing again the importance of these TFs in CCC. Considering that RUNX3, a key regulator in CCCs, also has a DMR in its promoter region, further analysis was performed on this TF. On this 831 bp DMR, 5 TFBSs are present (**Figure 5**). This DMR is targeted by at least 6 of the following 7 TFs: RUNX3, PAX5, YY1, SP1, MAX, EBF1 and IRF4. SP1 and PAX5 seem to be the most affine with these sequences. Although no complex composed only by at least 6 of these 7 TFs have been identified, PAX5 is always found associated to IRF4, whatever the size of the complex, suggesting an interaction between those 2 TFs.

**Figure 5.**
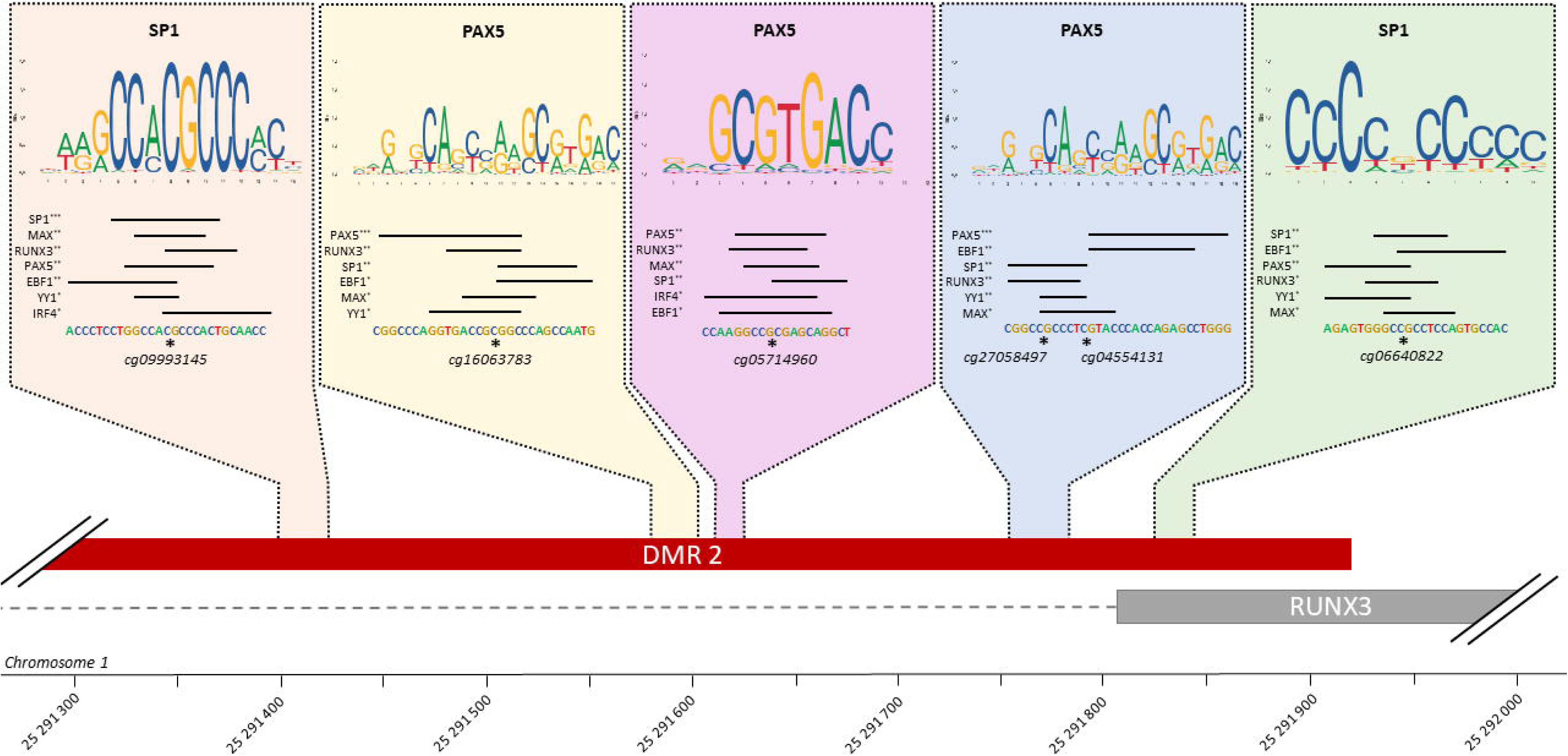
TFBS affected by methylation in RUNX3 regulatory region. Schematic representation of all TFBS found in RUNX3 regulatory region, using FIMO and Jaspar database. For each TFBS region, all the transcription factor predicted as affected by a differentiation of methylation in this region are rank by FIMO pvalue (***: pvalue ≤ 0.001, **: pvalue ≤ 0.01, *: pvalue ≤ 0.05). The top-rank TF binding profile is shown, as well as the differentially methylated position in the TFBS.

### Evaluation of cell types proportions in RNA-seq

Given the infiltration of immune cells in CCC myocardium, we looked for characterize and quantify the proportions of these cell types in our samples. To do so, we applied to our data, ADAPTS, a R tool to deconvolute the cell type proportions in RNA-seq by comparison to cell type signatures extracted from single-cell RNA-seq datasets (see Methods: Evaluation of cell types in heart tissue). In our study, we used two different single-cell datasets. First of all, immune cell signatures (36) (22 cell types) were used and showed in general a higher proportion of immune cells in CCCs compared to controls (**Supplementary Figure 6A**), such as activated NKs (Wilcoxon test, FDR=1.07E-02), and more interestingly T CD8, T Cell memory and T follicular helper (Wilcoxon test, respective FDR=1.07E-02; 1.07E-02; 1.83E-02). Secondly, a human heart tissue (left ventricle) cell signature (37) was used and showed that the CCCs had fewer cardiomyocytes (Wilcoxon test, FDR=1.16E-02) and smooth muscle (Wilcoxon test, FDR=3.33E-03) than the controls (**Supplementary Figure 6B**). Moreover, CCC myocardium had a higher proportion of macrophages (Wilcoxon test, FDR=1.90E-02), confirming the existence of immune infiltration in the cardiac tissues on CCC patients (**Supplementary table 13**).

### Blood methylation profile

Given the lack of myocardial tissues from patients with moderate CCC, our previous analyses were performed only on severe CCC. Therefore, it was impossible to study the progression of the pathogenic process during different stages of the disease. To overcome this problem, we studied the DNA methylation in the blood of 96 samples (33 asymptomatics and 63 CCCs (33 moderates and 30 severes)), by hypothesizing that the blood data reflect the phenotype. The analysis was made in two steps: first, we tested the difference of DNA methylation between asymptomatic and severe CCC, and then, we tested the difference of DNA methylation between moderate versus severe CCC samples to evaluate the disease progression.

Based on the low variability in DNA methylation levels observed in blood compared to tissue, (absolute values of tissue Δβ: mean = 4.96E-02, quartile 1 = 8.39E-03, quartile 3 = 7.38E-02; absolute values of blood Δβ: mean = 5.62E-03, q1 = 1.27E-03, q3 = 7.21E-03), only the FDR has been considered to define a DMP. We found 12624 DMPs between asymptomatic and severe CCC blood samples, with a FDR thresold of 0.05 (**Supplementary Table 14**). Of these, 7232 were found to be down-methylated (57.29%). 189 of those DMP (1.5%) are also found in the previous tissue DNA methylation analysis. Despite the small variation in the level of DNA methylation detected in the blood (Δβ), the methylation of these 12624 DMPs was enough to separate controls from cases, either via PCA (16.8% variability on PC1) (**Figure 6A**) or HCA (**Supplementary Figure 7A**). Association was found in 6436 genes with at least one DMP (ie. have one DMP in their body, promoter region or 3’UTR region), and 259 of those genes are also differentially expressed in RNA-seq analysis. Only 139 genes are in common between the three analyses (RNA-seq in tissue and DNA methylation in tissue and blood). As a result, there are few genes affected by expression and methylation in myocardial tissue and methylation in blood in common.

**Figure 6.**
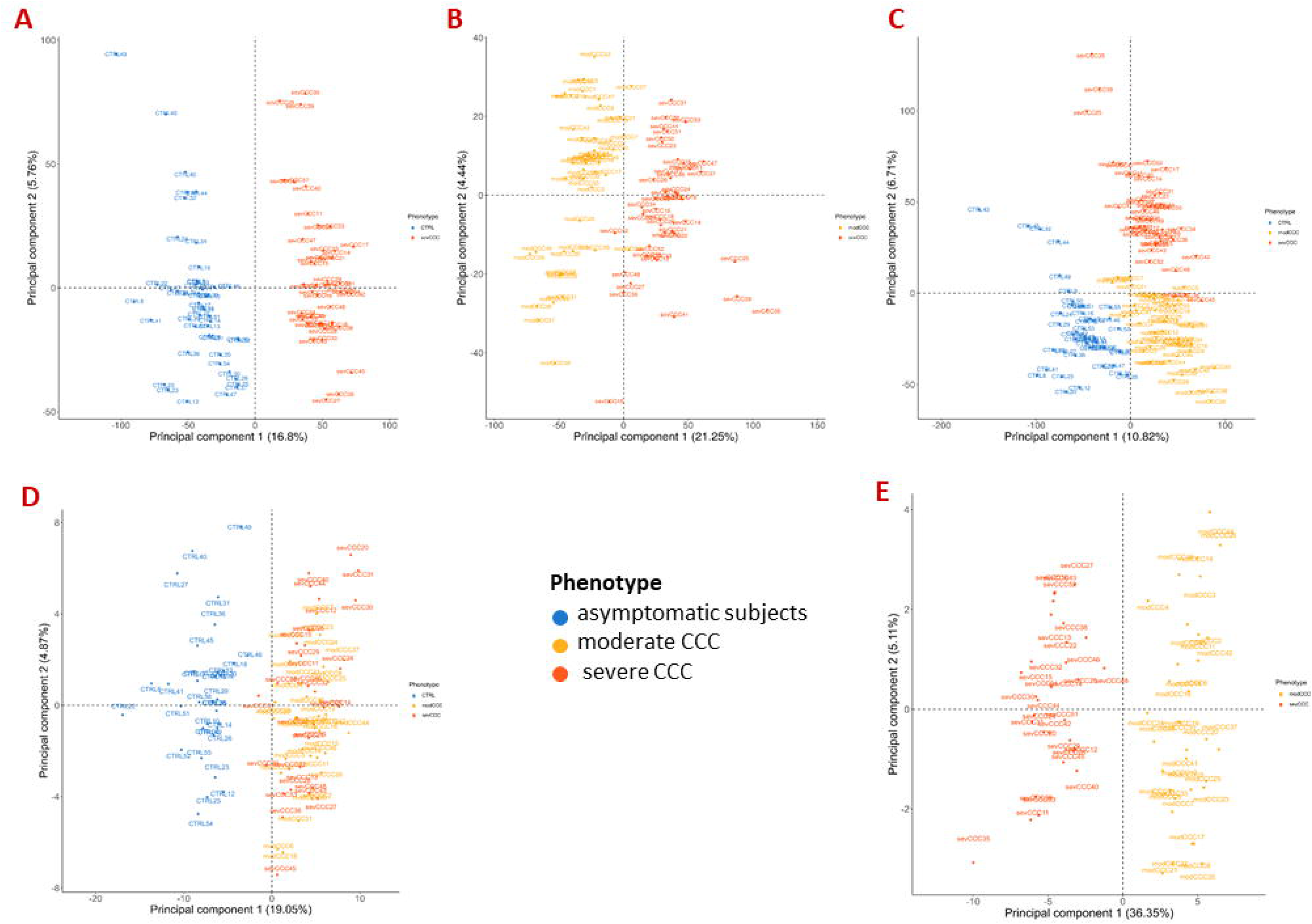
Analysis of samples distribution in the space of differentially methylated CpG sites for asymptomatic, moderate CCC and severe CCC samples. **(A)** Scatterplot of the two principal components of a PCA executed in the space of the 12624 CpG positions differentially methylated (DMP) between 48 asymptomatic blood samples and 90 CCC blood samples. **(B)** Scatterplot of the two principal components of a PCA executed in the space of the 6735 DMPs between 47 moderate CCC blood samples and 43 severe CCC blood samples. **(C)** Scatterplot of the two principal components of a PCA executed in the space of the 18889 CpG positions (union of the two previous sets) for the three groups of samples. **(D)** Scatterplot of the two principal components of a PCA executed in the space of the 198 DMPs between 33 asymptomatic blood samples and 63 CCC blood samples (training dataset). **(E)** Scatterplot of the two principal components of a PCA executed in the space of the 61 DMPs between 33 moderate CCC blood samples and 30 severe CCC blood samples (training dataset).

The disease progression was analyzed on 63 samples (33 moderate CCC and 30 severe CCC). 6735 CpGs were found as DMPs (**Supplementary Table 15**), with a FDR ≤0.05. Half of these DMPs (3178, 47.19%) were down-methylated. All the 6735 DMPs were enough to discriminate samples according to the stage of the disease on a PCA (21.25% of the variability in PC1) (**Figure 6B**). Similar conclusions were raised from HCA (**Supplementary Figure 7B**). Only 470 DMPs were also identified as DMP in controls vs severe CCC blood methylation analysis, and 1750 genes out of 3911 (44.75%) are shared by both blood methylation analysis (non-significant enrichment), showing epigenetic differences between the moderate and severe forms. Interestingly, looking at the DNA methylation level of all DMPs found in blood (merge of the two previous analysis, n=18889) allowed us to distinguish the three groups of individuals, according to their phenotype (**Figure 6C**), revealing a gradient of methylation in PC1xPC2 from controls to severe CCC through moderate CCC.

Finally, as on tissue methylation, the DMP distribution on genomic locations between asymptomatic and severe CCC (Chi square test T =30.63, p-value = 1.6E-04) was different for the original CpG distribution but not between moderate CCC and severe CCC (Chi square test T =12.57, p-value = 1.1E-01) (**Supplementary Table 16**).

### DNA methylation pattern on blood samples in severe CCC

The study of DNA methylation in the blood was done on 96 samples (33 asymptomatic, 33 moderate CCC and 30 severe CCC). We found 12624 CpGs between asymptomatic subjects and severe CCC. However, only a limited number of DMPs (n=189) is shared with the analysis done on heart tissue samples. The DNA methylation level variance detected in the blood was lower than the variation detected on tissues. Moreover, the general pattern detected on heart tissues and on blood are different and seemed to be tissue/fluid specific (**Figure 7A**). However, we conducted a Gene ontology analysis of the top 1000 genes of each analysis (RNA-seq (control vs severe CCC): 70.97%; tissue methylation (Control vs severe CCC): 17.20% and blood methylation (asymptomatic vs severe CCC): 15.54%) (**Supplementary Table 17**). Interestingly, all of the identified top GO are shared by all three analyses, showing that although few genes are found in common, they are involved in common biological functions. Therefore, three major biological processes are affected in our analyses. First, the immune system (1451 genes involved; 51.65%), including the lymphocyte activation (FDR=3.9E-21), the regulation of immune-system process (FDR=2.49E-16), the cytokine production (FDR=3.6E-14), the regulation of interleukin production (notably IL-2, FDR=2.93E-08; IL-4, FDR=8.42E-07; IL-6, FDR=1.44E-04) or the defense response (FDR=8E-10). Secondly, 557 genes (19.83%) are involved in biological processes related to system development (2.1E-16), including nervous system development (FDR=5.09E-09) and anatomical structure morphogenesis (FDR=7.64E-06), like cardiovascular system development (FDR=2.37E-03). Finally, 563 genes (20.04%) are linked to metal ion transport (6.39E-08), voltage-gated cation channel activity (1.51E-04) and cation homeostasis (6.09E-04) (**Figure 7B**). Even if the methylation patterns seem to be tissue/fluid specific, these patterns are concerned the same biological processes. Moreover, this biological process analogy is consistent with the gene expression analysis, finding similar biological pathways.

**Figure 7.**
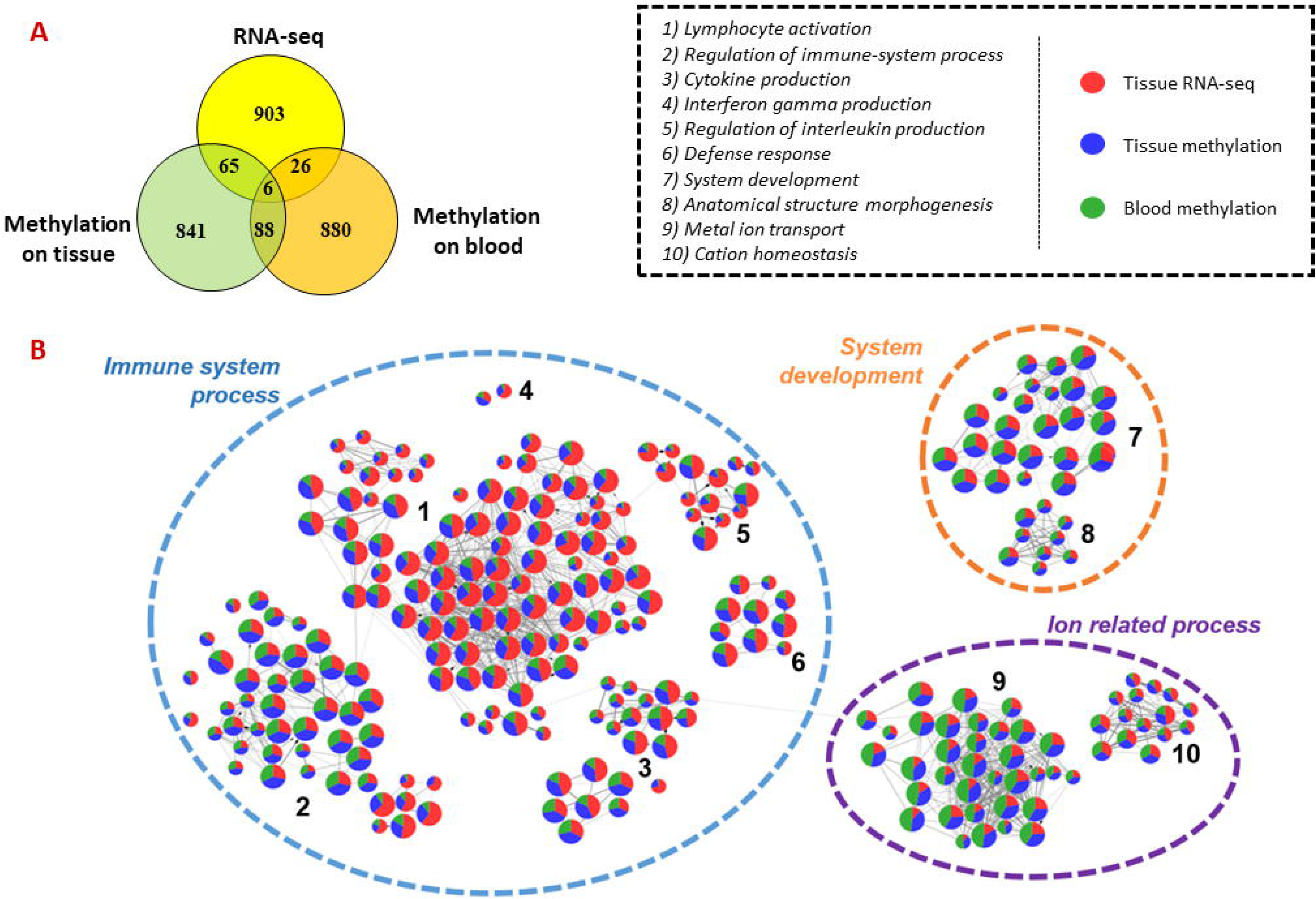
Comparison of differentially expressed genes, genes affected by methylation in tissue dataset and genes affected by methylation in blood dataset. **(A)** Venn diagram of top 1000 genes differentially expressed or methylated in previous tissue RNA-seq, tissue DNA methylation and blood DNA methylation analysis between control/asymptomatic and severe CCC samples. **(B)** Graph of the Gene ontology Biological Processes analysis of dysregulated element between control/asymptomatic and severe CCC. Nodes represents biological processes terms and are divided in 3 colors, according to the proportion of genes from RNA-seq (red), tissue methylation (blue) or blood methylation (green) analysis. Edges in the graph link GO terms having gene in common. 3 principal terms are highlighted in this synthesis. More precisely, several groups of gene ontology are enriched, involved in biological process related to : 1) lymphocyte activation; 2) Regulation of immune-system process; 3) Cytokine production; 4) Interferon gamma production; 5) Regulation of interleukin production; 6) Defense response; 7) System development; 8) Anatomical structure morphogenesis; 9) Metal ion transport; 10) Cation homeostasis.

Although all genes previously identified are involved in those biological pathways, whether they come from expression or methylation analysis, on tissue or on blood, some pathways seem to be mostly affected by one specific type of data. Indeed, the GOs related to the immune system are on average mostly carried by differentially expressed genes (49.12%), while the GOs related to system development (including nervous system development) and ion-related processes are mostly influenced by methylation.

### DNA methylation patterns on blood samples between moderate and severe CCC

Given the similarity between biological processes affected by modulation of the gene expression or by differences in DNA methylation in severe CCC, it seemed reasonable to analyze the methylation differences between moderate and severe CCC to understand the development of the disease (**Supplementary Table 18**). Unlikely to the results found between controls and severe CCC, here genes are mostly involved in neurogenesis (FDR=9.0E-18); anatomical structure morphogenesis (FDR=2.6E-13), including cardiovascular system development (FDR=1.1E-2); or actin filament organization (FDR=2.79E-09). To a lesser extent, these genes are associated with the immune response, notably in adaptive immune response (FDR=2.32E-07), including humoral immune response mediated by circulating immunoglobulin (FDR=9.05E-05), or B cell mediated immunity (FDR=3.16E-02). Finally, as found in previous results, genes are also associated with ion-related processes, like ion transport (FDR=1.56E-06) or sodium ion transmembrane transport (FDR=1.5E-05).

### Prediction of Chagas disease cardiomyopathy state based on blood DNA methylation

The point here was to identify the minimal number of markers allowing to predict the phenotype of patient. We aimed to identify two types of biomarkers: biomarkers indicating the presence of the disease (asymptomatic vs CCC), and those indicating its severity (moderate CCC vs severe CCC). For these two analyses, different machine learning models were fitted on 70% of the data (training data) and applied on the other 30% of the data (validation data). The models with the highest accuracy (TP+TN/all) were kept. In the first case, the number of DMPs previously found has been reduced, according to the delta beta value. In that case, the DMPs with a delta beta corresponding to the top 1 percentile have been retained. With these new filters, 198 DMPs were used to build a machine learning model (**Supplementary table 19**). The best parameters for the model were searched using a grid search, until we had an accuracy of 1 on our training data. The prediction was performed with a logistic regression, with a penalty L2 (Ridge) and a C of 5 (higher value of C specifies weaker regularization). When applied on our testing data, the final accuracy was 0.95, for a sensitivity of 1 and a specificity of 0.87. 40 individuals out of 42 in the testing data were thus correctly predicted. About these DMPs, 39% are in the gene body, 35% in intergenic regions, 24% in promoter regions and 2% in the 3’UTRs, which is not significantly different from the expected distribution (Chi square test p-value > 0.05). Among the genes with the highest number of DMPs is POU6F2 (n = 6), TM4SF1 (n = 5), CCDC144NL (n = 4), COLEC11 (n = 4) and ERICH1 (n = 4). All the genes associated to at least 1 of these 198 DMPs does not seems to be involved in any specific biological pathway.

In the second analysis (moderate CCC vs. severe CCC), all 61 DMPs were retained (**Supplementary table 19**). Since, the phenotype is more complex to predict, we used Random Forest algorithm algorithm. Using 100 trees with a maximum depth of 4, the validation data were predicted with an accuracy of 0.96 (sensitivity 0.92, specificity 1). Thus, 26 individuals out of 27 were correctly predicted. In this case, some regions are significantly enriched in DMPs (Chi-square test p-value = 2.2E-06), such as 41% of DMPs in the gene body, 35% in promoters, 22% in intergenic regions and 2% in the 3’UTRs. All 61 DMPs are located in 49 genes, but only 1 by gene. Of these genes, 6 of these genes are notably involved in the regulation of the Wnt signaling pathway (FDR = 6.5E-07): ARNTL, BTRC, GLI3, LZTS2, PTPRO, RNF220.

## Discussion

In a previous analysis, combining microarray and methylation data (Illumina 450k), we showed that patients with CCCs had significant differences in gene expression and DNA methylation (17). DEGs with DMPs were predominantly associated with the immune response, but also with several biological processes sur as arrhthmia, muscle contraction, fibrosis and mitochondria’s function. In vitro analyses had showed that differential methylation of some promoters had an impact on the gene expression and on protein production for immune- or heart-related genes, including KCNA4 and RUNX3. In the present study, we have set up more advanced analysis (RNA-seq and EPIC methylation), allowing us to have an in-depth analysis of genomic deregulation. In addition, we also conducted a DNA methylation analysis on blood samples from patients and asymptomatic controls.

All these approaches combined have put forward different pathways affected in patients with CCCs. Although the immune response seems to be strongly stimulated, especially the activation of T cells even in the absence of the parasite. This infiltrate, already known (47), is mainly composed of Th1 lymphocytes, macrophages and NK-cells. Our deconvolution analysis done on our heart tissue collection, had confirmed the content of this infiltrate. Our group, among others, have already shown that the T-cell infiltrating heart tissue strongly produced INF-γ and TNF-α (8). In parallel, lower quantities of IL-2, IL-4, IL-6 and IL-10 were detected in CCC heart tissue (48). Our results are consistent with current knowledge of the pathology, with high expression of genes involved in INF-γ and cytokine production, and regulation of interleukin production, including IL-2, IL-4 and IL-6. IFN-γ, which has been described simultaneously as a pathogen resistance and a disease tolerance gene is also acting as an upstream regulator (49). Overproduction of IFN-γ can induce tissue damage and death in the chronic phases. Indeed, IFN-γ stimulates the inflammatory response via the NF-κB pathway and activate the production of ROS and NOS, which, in an excessive quantity, have also deleterious effect on mitochondria and cardiomyocytes (12). Interestingly, we found a DMR in IFN-γ promoter region, targeted by RUNX3 and TBX21 (T-bet) transcription factors by interacting with GATA3 (50, 51). The micro-RNA MIR142, potentially target by RUNX3 and TBX21 according to our analysis, is also associated to Th1 differentiation (52) in neuronal autoimmune disease. In Chagas disease, TBX21 and IFN-γ expression are correlated with the left ventricular dilatation, and the ratio between TBX21 and GATA3 expression is significantly higher in CCC than in non-inflammatory cardiomyopathy (53), which is also the case in our data, whereas we compare CCC to control or DCM. Moreover, RUNX3 overexpression has been associated to the methylation of his promoter in CCC (17), as well as in our analysis. All our results are therefore consistent with our knowledge of this pathology, and have allowed us to identify a whole list of genes potentially deregulated by the action of these 3 TFs that are known to be involved in CCC.

In addition to this exacerbate immune response, genes involved in processes related to cardiac contraction, ion transport, nervous system, fibrosis or mitochondria have been identified as differentially expressed and/or differentially methylated. In particular, the nervous system appears to be strongly impacted between moderate and severe CCCs. These genes seem to be targeted by 6 transcription factors, involved in cardiac processes and nervous system, reinforcing this signal. Our methylation analysis, added to the previous studies carried out (17, 54), confirmed that epigenetic regulations are essential in Chagas disease. Among the genes associated to ion transport, and more precisely with voltage-gated ion channel, are KCNA2, KCNA5, KCNAB2, KCNB2, KCNC2, KCNG3 and KCNN4. Potassium channel gene deregulation has already been described in mouse (55) and human heart (17). Calcium and potassium ion channels functions are affected in mouse heart tissue (56), leading to a reduced Ca^2+^ release and a prolongation of action potential, showing a cardiomyocyte dysfunction. TNF-α may amplify this dysfunction, by inducing nitric oxide synthase (iNOS) and oxidant species, which promotes electrophysiological changes, as it has been shown in rat ventricular myocyte (57). KCNN4, also called KCa3.1 (calcium-activated potassium channel), act as a regulator of membrane potential in T cells. After antigen recognition by the T cell receptoire, the Ca^2+^ enter in the cytosol, which lead to the activation of KCa3.1, and then to Kv1.3 with the membrane depolarization (58). This calcium will activate the NFAT protein, which regulate genes involving in T cell activation (59). Moreover, high levels of Kv1.3 were found in multiple autoimmune disease (60–62). In our data, these two potassium channels are up-regulated (Kv1.3: log2(FC) = 4.01, FDR = 4.86E-15 KCa3.1: log2(FC) = 1.56, FDR = 2.11E-03), and KCa3.1 seems to be targeted by RUNX3. Galectin-3 (Gal-3) has been regarded as a potential biomarker for heart disease and a causative mediator of cardiac fibrosis. Gal-3 promotes cardiac fibrosis through upregulating the expression and activity of KCa3.1 channel in inflammatory cells and fibroblasts and that upregulated KCa3.1 facilitates inflammatory cell infiltration into the myocardium and fibroblast differentiation into activated form. Recently, it has been shown mice treated with TRAM-34, a KCa3.1 channel-specific inhibitor, either for 1- or 2-month period effectively reduced collagen deposition (63). KCa3.1 inhibition by TRAM-34 therapy attenuated the increased inflammatory cell infiltration. Moreover, Gang et al. had shown that KCa3.1 inhibition by TRAM-34 therapy attenuated the increased inflammatory cell infiltration. Our study underlined that several genes associated with moderate or severe CCC are involved in the nervous system. This enrichment is even stronger in moderate CCCs. Actually, the loss of neuron cells is a well-known phenomenon in digestive forms of Chagas disease (64, 65). In human hearts with dilated cardiomyopathy, the number of neuron cells is significantly reduced compared to controls. This neuronal depopulation is even more important in the CCC (66), and particularly during the acute phase in mice model (67). In the dog heart, during the acute phase, a rarefaction of the noradrenergic and acetylcholine nerve terminals has been observed only when myocarditis occurs, and the ventricles sympathetic denervation was present when the inflammatory process was moderate to intense (68). Moreover, in the rat model, moderate myocarditis lasting for two weeks may cause complete denervation (69). A more recent study demonstrates that the lack of acetylcholine in mice heart tissue seems to promote the Th1 response, inducing a reduction of the circulating parasites, but a worsening of the cardiac lesions and inflammatory infiltration (70). In the continuity of these observations, the increase in the amount of acetylcholine reduces the weight of heart mice, the inflammatory infiltration and the fibrosis area, suggesting a reversing of cardiac hypertrophy. Those observations are correlated with a reduced level of IFN-γ in serum (71). Moreover, number studies have shown the impact of the nervous system on immune-response regulation. In short, macrophages, Th1, CD4 and B cells express adrenergic receptors (72–74). Their stimulation induces an increase of cAMP, which inhibits NF-kB activation. This leads to the suppression of type 1 pro-inflammatory cytokines and promotes the production of type 2 anti-inflammatory cytokines, as IL10. This cytokine inhibits the antigen presenting capacity of macrophages and dendritic cells, and thus the differentiation of CD4 T cells to Th1 (75). In our data, the genes related to acetylcholine production, or acetylcholine transporter are not differentially methylated or expressed in tissue samples according to fold change cut-off, but present a FDR≤0.05. In blood methylation samples, ACHE (acetylcholine production), and CHRNA3, CHRNA4 and CHRNA7 (acetylcholine receptors) genes have differentially methylated positions, in promoter or body. All these observations on animal models, together with our data showing the involvement of genes related to the nervous system in CCCs, especially in the moderate phase, strongly suggest an involvement of the nervous system in the shift of the immune response towards a Th1 response, inducing heart tissue damage. In particular, the involvement of acetylcholine seems to be related to inflammatory infiltration and fibrosis. Finally, circulating antibodies binding acetylcholine and norepinephrine receptors have been found in Chagas patient serum, suggesting an autoimmune response (Borda 1996 10.1016/0167-5273(96)02592-2), and, correlated with the previous study, inducing the lack of neurotransmitters, and then the over production of Th1 cells.

Non coding RNA, notably lncRNA and miRNA, are known to be involved in many of cardiovascular diseases (55, 76), mostly by acting directly on gene expression or protein stability for miRNAs, and by repressing the action of miRNAs by acting as a sponge for lncRNAs. The presented results extend the list of miRNAs associated to CCC, and not differentially expressed in DCM (18, 55, 77–83). Among the 19 miRNAs identified in this study, 3 have already been associated with severe CCC: MIR-223, MIR-208 and MIR-151 (14). In addition, the long non coding MIR155HG codes for MIR-155, which is also found as up-regulated in severe CCC. Interestingly, this miRNA is responsible for many inflammatory stimuli, including TNF-α and interferons (84). Moreover, it is up-regulated in viral myocarditis; where it is expressed by infiltrating immune cells, and seems to be involved in TNF-α, IFN-γ and IL-6 production, as well as immune cell infiltration (85). Finally, the lack of this microRNA seems to decrease IFN-γ and TNF-α in the acute stage of Chagas cardiomyopathy in mice heart tissue (55). Several studies are ongoing to test specific agonist or antagonist effects of the miRNAs that may use in the future to avoid or slow disease progression. Several encouraging studies have led to interesting modeling, in animals at least (86, 87). In addition, several lncRNAs were also differentially expressed in CCC patient tissues. Among them, MIAT has been associated to multiple diseases leading to various phenotypes (88). In breast cancer, it targets the microRNA-133, dysregulated in CCC (18), and down-regulated in patients with heart failure (89). This lncRNA is always acting as a sponge for specific miRNAs. Moreover, XIST is one of the top up-regulated lncRNA in our disease. It is associated with the WNT1 gene, belonging to the Wnt family. Because of the sex dispersion in our dataset, it is hard to confirm its involvement in CCC. However, WNT1, as well as WNT10A and WNT10B are up-regulated in severe CCC, suggesting their involvement in this pathology. In addition, the DMPs used as biomarkers to determine whether patients are in moderate or severe phase are located in 6 genes related to the Wnt pathway: ARNTL, BTRC, GLI3, LZTS2, PTPRO and RNF220. The Wnt signaling pathway has a role in cardiac development, and is involved in cardiovascular disease (90) including fibrosis (91–93). After infection by T. cruzi, Wnt expression is induced in macrophages, and Wnt signaling has a role in the regulation of the immune response, as well as in the parasite replication (90). In fibrosis, some study confirmed the interaction between the Wnt pathway and TGF-β signalling (94–96). TGF-β receptor overexpression has already been observed in acute phase of CCC (97), and his role in CCC development has been confirmed, including parasitic invasion, inflammation, immune response, heart fibrosis and heart conduction (98). A therapeutic strategy, based on TGF-β inhibition, has already been proposed and try on mouse (80). Moreover, some study suggest an interaction between the Wnt pathway, the TGF-β signalling and the NOTCH signaling (99, 100). Our result showing the involvement of the Wnt pathway in the evolution of the disease and the potential implication of NOTCH1 in the genetic deregulation in severe CCC reinforce the hypothesis of the Wnt-NOTCH-TGF-β in CCC evolution. For the others, their modes of action are not obvious at all. Several approaches are ongoing such as Loss-of-Function Strategies for lncRNAs, lncRNA-Interactome Analyses, Identification of lncRNAs Associated with Translation Machinery (review in (101)) but a database, namely a well annotated one, is slow to come. Therefore, the use of these molecules as biomarkers and even more as complementary treatments remains a distant prospect.

Although present in many countries, Chagas disease affects mostly poor populations with little access to health services (102). As a result, providing diagnosis and treatment for millions of people is proving difficult, and this disease has been characterized as “the most neglected of the neglected diseases”, in terms of research and development. In the acute phase of the disease, the most effective method of diagnosis is the direct detection of antigens produced by T. cruzi, although this method suffers from limitations in terms of specificity (∼80-90%) (103). Therefore, diagnosis is often done by microscopy (104). With regard to the chronic phase, due to low or no parasitic activity, direct detection of T. cruzi cannot be considered. In addition, recent qPCR-based methods have been tested on acute and chronic samples, but have shown low sensitivity (∼80%), and require financial and material resources that are difficult to implement in endemic areas (105). Recently, blood markers typical of inflammatory responses and tissue damage due to chagas disease, including IFN-y, IL-6, TNF-a, NTproBNP (among others) have been proposed as biomarkers of CCC (106). However, the low specificity of these markers, or their lack of specificity to CCCs, such as NTproBNP, representative of cardiac pathologies (107); does not allow for clinical use at the present time. In our study, we have identified 198 CCC-specific methylation markers. Those CpGs could distinguish controls to CCC from blood samples with 100% of sensitivity and 97% of specificity in independent validation sets. In addition, 61 CpGs have been identified, allowing to predict the progression of this pathology (from moderate to severe CCC), with a sensitivity of 92% and a specificity of 100%. In order to propose large-scale reproducible biomarkers, a consensus Target Product Profile (TPP) has been developed for Chagas disease (108), stipulating that markers should be able to detect the effects of drug treatments, be detectable with limited resources and not vary according to the strain of the parasite. Given the high specificity of these two assays, these methylation sites appear to be good candidates for use as blood biomarkers, and further studies will be necessary to potentially validate their possible use in the clinic, in accordance with the TPP consensus.

All the results obtained in our study converge towards a combined involvement of processes related to the immune response, ion transport, cardiac contraction and the nervous system. Gene expression analysis alone has revealed some non-coding elements, but has not provided so much new information about the disease. However, the inclusion of methylation put forward less obvious biological process. Thus, the majority of the identified genes (differentially expressed and/or methylated) are generally involved in several of these processes, highlighting links between them. These results, combined with those obtained in previous analyses of chagasic cardiomyopathies (17, 54), confirm the importance of DNA methylation in the development of CCCs.

## Data Availability

All data produced in the present study are available upon reasonable request to the authors

## Acknowledgments

Center de Calcul Intensif d’Aix-Marseille is acknowledged for granting access to its high performance computing resources. Authors thanks Benoit Ballester, Pierre Milpied, Delphine Potier for helpful discussions and comments.

## Supplementary materials

**Supplementary table 1.** Cell lines description.

**Supplementary table 2.** Statistics on coding sequences detected by RNA-seq and on differentially expressed genes.

**Supplementary table 3.** List of unique differentially expressed genes identified by RNA-seq between 8 end stage CCC heart tissue samples and 6 heart tissue samples obtained from organ donors.

**Supplementary table 4.** List of unique differentially expressed genes identified by RNA-seq between 8 DCM heart tissue samples and 6 heart tissue samples obtained from organ donors.

**Supplementary table 5.** Gene ontology analysis based on differentially expressed genes between control and severe CCC and/or between control and DCM.

**Supplementary table 6.** List of differentially expressed ncRNAs between control and severe CCC, or between control and dilated cardiomyopathies.

**Supplementary table 7.** Features of the tested CpG sites and of the DMPs.

**Supplementary table 8.** List of the Differentially Methylated CpGs on heart tissues samples (controls versus CCC).

**Supplementary table 9.** List of the regulatory DMRs associated to the 89 DEGs.

**Supplementary table 10.** List of transcription factor and transcription factor complexes testing against our DMRs.

**Supplementary table 11.** Gene ontology analysis of the TFs binding our DMRs.

**Supplementary table 12.** List of differentially expressed ncRNAs between control and severe CCC, or between control and dilated cardiomyopathies.

**Supplementary table 13.** Cell type proportions in RNA-seq by comparison to cell type signatures.

**Supplementary table 14.** List of the Differentially Methylated CpGs between blood asymptomatic samples and blood CCC samples.

**Supplementary table 15.** List of the Differentially Methylated CpGs between blood moderate CCC samples and blood severe CCC samples.

**Supplementary table 16.** Features of the DMPs on blood samples.

**Supplementary table 17.** Gene ontology analysis based on differentially expressed genes between control and severe CCC and/or genes affected by at least one differentially methylated CpGs between tissue control and severe CCC and/or genes affected by at least one differentially methylated CpGs between blood asymptomatic and severe CCC samples.

**Supplementary table 18.** Gene ontology analysis based on genes affected by at least one differentially methylated CpGs between blood moderate CCC samples and blood severe CCC samples.

**Supplementary figure 1. Analysis of samples distribution in the space of differentially expressed genes or differentially methylated CpG sites between control and severe CCC**

(A) Scatterplot of the two first principal component of a PCA of 6 Control and 8 Case samples executed in the space of the 1409 differentially expressed genes between the two conditions. B) Scatterplot of the two first principal component of a PCA of 6 Control and 8 Case samples executed in the space of the 16883 differentially metylated CpG sites between the two conditions.

**Supplementary figure 2. Age and sex impact on samples distribution in the space of differentially expressed genes or differentially methylated CpG sites between control and severe CCC**

**(A)** Scatterplot of the two first principal component of a PCA of 6 Control and 8 Case samples executed in the space of the 1409 differentially expressed genes between the two conditions colored by sex (blue: male, pink : female). **(B)** Scatterplot of the two first principal component of a PCA of 6 Control and 8 Case samples executed in the space of the 1409 differentially expressed genes between the two conditions colored by age (from grey to blue). **(C)** Scatterplot of the two first principal component of a PCA of 6 Control and 8 Case samples executed in the space of the 16883 differentially metylated CpG sites between the two conditions colored by sex (blue: male, pink : female). **(D)** Scatterplot of the two first principal component of a PCA of 6 Control and 8 Case samples executed in the space of the 16883 differentially metylated CpG sites between the two conditions colored by age (from grey to blue).

**Supplementary figure 3. Gene Ontology Biological Process affected in severe CCC and/or DCM**

Scatterplot of Gene Ontology Biological Process according to percent of severe CCC differentially expressed genes (DEG) and percent of total DEG (severe CCC + DCM) involved in each GO term. The size of each dot is associated to the enrichment of each GO term (- log_10_(FDR)) and its color to disease specificity (from green for DCM to red for severe CCC).

**Supplementary figure 4. ncRNA analysis in severe CCC and DCM**

Control samples identifiers are written in blue, severe CCC samples identifiers in red and DCM samples in green. **(A)** Hierarchical Clustering Analysis (HCA) performed on 6 control and 8 severe CCC samples, based on expression of 179 differentially expressed ncRNAs. **(B)** Hierarchical Clustering Analysis (HCA) performed on 6 control and 8 DCM samples, based on expression of 327 differentially expressed ncRNAs. **(C)** Venn diagram of shared differentially expressed ncRNAs between severe CCC and DCM.

**Supplementary figure 5. Predicted regulatory interaction in cardiac muscular or nervous system process**

Network composed by 6 transcription factors (TF) involved in cardiac muscular and/or nervous system process, and their 68 targeted genes, predicted by *OLOGRAM* according to *ReMap* database. TF are written in blue in diamond, and genes in black in rectangle. Shapes borders are colored according to the fold change, from green to red.

**Supplementary figure 6. Estimation of cell proportion in control and severe CCC heart tissue**

Deconvolution of the RNA-seq bulk gene expressions of 6 control and 8 severe CCC samples to infer the proportion of cells in the samples. The * represent cell types whose proportion is significantly different (Wilcoxon test, FDR <0.05) between controls and cases. **(A)** Deconvolution using cell type signature with 22 immunological cell types (LM22 signature matrix). **(B)** Deconvolution using cell type signature with 5 left ventricle related cell types.

**Supplementary figure 7. Analysis of samples clustering based on differentially methylated CpG sites in blood samples**

Asymptomatic samples identifiers are written in blue, moderate CCC in orange and severe CCC in red. **(A)** Hierarchical Clustering Analysis (HCA) performed on 48 asymptomatic and 90 severe CCC samples, based on expression of 12624 differentially methylated position. **(B)** Hierarchical Clustering Analysis (HCA) performed on 47 moderate CCC and 43 severe CCC samples, based on methylation level of 6735 differentially methylated position.

## References

1. Perez-Molina JA, and Molina I. Chagas disease. Lancet. 2018;391(10115):82–94.

2. Hotez PJ, Dumonteil E, Betancourt Cravioto M, Bottazzi ME, Tapia-Conyer R, Meymandi S, et al. An unfolding tragedy of Chagas disease in North America. PLoS Negl Trop Dis. 2013;7(10):e2300.

3. Antinori S, Galimberti L, Bianco R, Grande R, Galli M, and Corbellino M. Chagas disease in Europe: A review for the internist in the globalized world. Eur J Intern Med. 2017;43:6–15.

4. Imai K, Maeda T, Sayama Y, Osa M, Mikita K, Kurane I, et al. Chronic Chagas disease with advanced cardiac complications in Japan: Case report and literature review. Parasitol Int. 2015;64(5):240–2.

5. Jackson Y, Pinto A, and Pett S. Chagas disease in Australia and New Zealand: risks and needs for public health interventions. Trop Med Int Health. 2014;19(2):212–8.

6. Freitas HF, Chizzola PR, Paes AT, Lima AC, and Mansur AJ. Risk stratification in a Brazilian hospital-based cohort of 1220 outpatients with heart failure: role of Chagas’ heart disease. Int J Cardiol. 2005;102(2):239–47.

7. Morillo CA, Marin-Neto JA, Avezum A, Sosa-Estani S, Rassi A, Jr., Rosas F, et al. Randomized Trial of Benznidazole for Chronic Chagas’ Cardiomyopathy. N Engl J Med. 2015;373(14):1295–306.

8. Abel LC, Rizzo LV, Ianni B, Albuquerque F, Bacal F, Carrara D, et al. Chronic Chagas’ disease cardiomyopathy patients display an increased IFN-gamma response to Trypanosoma cruzi infection. J Autoimmun. 2001;17(1):99–107.

9. Higuchi Mde L, Gutierrez PS, Aiello VD, Palomino S, Bocchi E, Kalil J, et al. Immunohistochemical characterization of infiltrating cells in human chronic chagasic myocarditis: comparison with myocardial rejection process. Virchows Arch A Pathol Anat Histopathol. 1993;423(3):157–60.

10. Reis DD, Jones EM, Tostes S, Jr., Lopes ER, Gazzinelli G, Colley DG, et al. Characterization of inflammatory infiltrates in chronic chagasic myocardial lesions: presence of tumor necrosis factor-alpha+ cells and dominance of granzyme A+, CD8+ lymphocytes. Am J Trop Med Hyg. 1993;48(5):637–44.

11. Rocha Rodrigues DB, dos Reis MA, Romano A, Pereira SA, Teixeira Vde P, Tostes S, Jr., et al. In situ expression of regulatory cytokines by heart inflammatory cells in Chagas’ disease patients with heart failure. Clin Dev Immunol. 2012;2012:361730.

12. Cunha-Neto E, Dzau VJ, Allen PD, Stamatiou D, Benvenutti L, Higuchi ML, et al. Cardiac gene expression profiling provides evidence for cytokinopathy as a molecular mechanism in Chagas’ disease cardiomyopathy. Am J Pathol. 2005;167(2):305–13.

13. Nogueira LG, Santos RH, Ianni BM, Fiorelli AI, Mairena EC, Benvenuti LA, et al. Myocardial chemokine expression and intensity of myocarditis in Chagas cardiomyopathy are controlled by polymorphisms in CXCL9 and CXCL10. PLoS Negl Trop Dis. 2012;6(10):e1867.

14. Laugier L, Ferreira LRP, Ferreira FM, Cabantous S, Frade AF, Nunes JP, et al. miRNAs may play a major role in the control of gene expression in key pathobiological processes in Chagas disease cardiomyopathy. PLoS Negl Trop Dis. 2020;14(12):e0008889.

15. Jo BS, Koh IU, Bae JB, Yu HY, Jeon ES, Lee HY, et al. Methylome analysis reveals alterations in DNA methylation in the regulatory regions of left ventricle development genes in human dilated cardiomyopathy. Genomics. 2016;108(2):84–92.

16. Haas J, Frese KS, Park YJ, Keller A, Vogel B, Lindroth AM, et al. Alterations in cardiac DNA methylation in human dilated cardiomyopathy. EMBO Mol Med. 2013;5(3):413–29.

17. Laugier L, Frade AF, Ferreira FM, Baron MA, Teixeira PC, Cabantous S, et al. Whole-Genome Cardiac DNA Methylation Fingerprint and Gene Expression Analysis Provide New Insights in the Pathogenesis of Chronic Chagas Disease Cardiomyopathy. Clin Infect Dis. 2017;65(7):1103–11.

18. Ferreira LR, Frade AF, Santos RH, Teixeira PC, Baron MA, Navarro IC, et al. MicroRNAs miR-1, miR-133a, miR-133b, miR-208a and miR-208b are dysregulated in Chronic Chagas disease Cardiomyopathy. Int J Cardiol. 2014;175(3):409–17.

19. Frade AF, Laugier L, Ferreira LR, Baron MA, Benvenuti LA, Teixeira PC, et al. Myocardial Infarction-Associated Transcript, a Long Noncoding RNA, Is Overexpressed During Dilated Cardiomyopathy Due to Chronic Chagas Disease. J Infect Dis. 2016;214(1):161–5.

20. Pidsley R, Zotenko E, Peters TJ, Lawrence MG, Risbridger GP, Molloy P, et al. Critical evaluation of the Illumina MethylationEPIC BeadChip microarray for whole-genome DNA methylation profiling. Genome Biol. 2016;17(1):208.

21. Frade AF, Teixeira PC, Ianni BM, Pissetti CW, Saba B, Wang LH, et al. Polymorphism in the alpha cardiac muscle actin 1 gene is associated to susceptibility to chronic inflammatory cardiomyopathy. PLoS One. 2013;8(12):e83446.

22. Bolger AM, Lohse M, and Usadel B. Trimmomatic: a flexible trimmer for Illumina sequence data. Bioinformatics. 2014;30(15):2114–20.

23. Dobin A, Davis CA, Schlesinger F, Drenkow J, Zaleski C, Jha S, et al. STAR: ultrafast universal RNA-seq aligner. Bioinformatics. 2013;29(1):15–21.

24. Okonechnikov K, Conesa A, and Garcia-Alcalde F. Qualimap 2: advanced multi-sample quality control for high-throughput sequencing data. Bioinformatics. 2016;32(2):292–4.

25. Liao Y, Smyth GK, and Shi W. featureCounts: an efficient general purpose program for assigning sequence reads to genomic features. Bioinformatics. 2014;30(7):923–30.

26. Molder F, Jablonski KP, Letcher B, Hall MB, Tomkins-Tinch CH, Sochat V, et al. Sustainable data analysis with Snakemake. F1000Res. 2021;10:33.

27. Love MI, Huber W, and Anders S. Moderated estimation of fold change and dispersion for RNA-seq data with DESeq2. Genome Biol. 2014;15(12):550.

28. Ito K, and Murphy D. Application of ggplot2 to Pharmacometric Graphics. CPT Pharmacometrics Syst Pharmacol. 2013;2:e79.

29. Bindea G, Mlecnik B, Hackl H, Charoentong P, Tosolini M, Kirilovsky A, et al. ClueGO: a Cytoscape plug-in to decipher functionally grouped gene ontology and pathway annotation networks. Bioinformatics. 2009;25(8):1091–3.

30. Luo W, and Brouwer C. Pathview: an R/Bioconductor package for pathway-based data integration and visualization. Bioinformatics. 2013;29(14):1830–1.

31. Luo W, Friedman MS, Shedden K, Hankenson KD, and Woolf PJ. GAGE: generally applicable gene set enrichment for pathway analysis. BMC Bioinformatics. 2009;10:161.

32. Cheng L, Wang P, Tian R, Wang S, Guo Q, Luo M, et al. LncRNA2Target v2.0: a comprehensive database for target genes of lncRNAs in human and mouse. Nucleic Acids Res. 2019;47(D1):D140–D4.

33. Zhao H, Shi J, Zhang Y, Xie A, Yu L, Zhang C, et al. LncTarD: a manually-curated database of experimentally-supported functional lncRNA-target regulations in human diseases. Nucleic Acids Res. 2020;48(D1):D118–D26.

34. Bao Z, Yang Z, Huang Z, Zhou Y, Cui Q, and Dong D. LncRNADisease 2.0: an updated database of long non-coding RNA-associated diseases. Nucleic Acids Res. 2019;47(D1):D1034–D7.

35. Danziger SA, Gibbs DL, Shmulevich I, McConnell M, Trotter MWB, Schmitz F, et al. ADAPTS: Automated deconvolution augmentation of profiles for tissue specific cells. PLoS One. 2019;14(11):e0224693.

36. Newman AM, Liu CL, Green MR, Gentles AJ, Feng W, Xu Y, et al. Robust enumeration of cell subsets from tissue expression profiles. Nat Methods. 2015;12(5):453–7.

37. Wang L, Yu P, Zhou B, Song J, Li Z, Zhang M, et al. Single-cell reconstruction of the adult human heart during heart failure and recovery reveals the cellular landscape underlying cardiac function. Nat Cell Biol. 2020;22(1):108–19.

38. Tian Y, Morris TJ, Webster AP, Yang Z, Beck S, Feber A, et al. ChAMP: updated methylation analysis pipeline for Illumina BeadChips. Bioinformatics. 2017;33(24):3982–4.

39. Teschendorff AE, Marabita F, Lechner M, Bartlett T, Tegner J, Gomez-Cabrero D, et al. A beta-mixture quantile normalization method for correcting probe design bias in Illumina Infinium 450 k DNA methylation data. Bioinformatics. 2013;29(2):189–96.

40. Chen C, Grennan K, Badner J, Zhang D, Gershon E, Jin L, et al. Removing batch effects in analysis of expression microarray data: an evaluation of six batch adjustment methods. PLoS One. 2011;6(2):e17238.

41. Bibikova M, Le J, Barnes B, Saedinia-Melnyk S, Zhou L, Shen R, et al. Genome-wide DNA methylation profiling using Infinium(R) assay. Epigenomics. 2009;1(1):177–200.

42. Peters TJ, Buckley MJ, Statham AL, Pidsley R, Samaras K, R VL, et al. De novo identification of differentially methylated regions in the human genome. Epigenetics Chromatin. 2015;8:6.

43. Cheneby J, Gheorghe M, Artufel M, Mathelier A, and Ballester B. ReMap 2018: an updated atlas of regulatory regions from an integrative analysis of DNA-binding ChIP-seq experiments. Nucleic Acids Res. 2018;46(D1):D267–D75.

44. Ferre Q, Charbonnier G, Sadouni N, Lopez F, Kermezli Y, Spicuglia S, et al. OLOGRAM: Determining significance of total overlap length between genomic regions sets. Bioinformatics. 2019.

45. Razin A, and Cedar H. DNA methylation and gene expression. Microbiol Rev. 1991;55(3):451–8.

46. Moore LD, Le T, and Fan G. DNA methylation and its basic function. Neuropsychopharmacology. 2013;38(1):23–38.

47. Bonney KM, and Engman DM. Autoimmune pathogenesis of Chagas heart disease: looking back, looking ahead. Am J Pathol. 2015;185(6):1537–47.

48. Machado FS, Dutra WO, Esper L, Gollob KJ, Teixeira MM, Factor SM, et al. Current understanding of immunity to Trypanosoma cruzi infection and pathogenesis of Chagas disease. Semin Immunopathol. 2012;34(6):753–70.

49. Chevillard C, Nunes JPS, Frade AF, Almeida RR, Pandey RP, Nascimento MS, et al. Disease Tolerance and Pathogen Resistance Genes May Underlie Trypanosoma cruzi Persistence and Differential Progression to Chagas Disease Cardiomyopathy. Front Immunol. 2018;9:2791.

50. Kanhere A, Hertweck A, Bhatia U, Gokmen MR, Perucha E, Jackson I, et al. T-bet and GATA3 orchestrate Th1 and Th2 differentiation through lineage-specific targeting of distal regulatory elements. Nat Commun. 2012;3:1268.

51. Kohu K, Ohmori H, Wong WF, Onda D, Wakoh T, Kon S, et al. The Runx3 transcription factor augments Th1 and down-modulates Th2 phenotypes by interacting with and attenuating GATA3. J Immunol. 2009;183(12):7817–24.

52. Talebi F, Ghorbani S, Chan WF, Boghozian R, Masoumi F, Ghasemi S, et al. MicroRNA-142 regulates inflammation and T cell differentiation in an animal model of multiple sclerosis. J Neuroinflammation. 2017;14(1):55.

53. Nogueira LG, Santos RH, Fiorelli AI, Mairena EC, Benvenuti LA, Bocchi EA, et al. Myocardial gene expression of T-bet, GATA-3, Ror-gammat, FoxP3, and hallmark cytokines in chronic Chagas disease cardiomyopathy: an essentially unopposed TH1-type response. Mediators Inflamm. 2014;2014:914326.

54. Casares-Marfil D, Kerick M, Andres-Leon E, Bosch-Nicolau P, Molina I, Chagas Genetics CN, et al. GWAS loci associated with Chagas cardiomyopathy influences DNA methylation levels. PLoS Negl Trop Dis. 2021;15(10):e0009874.

55. Navarro IC, Ferreira FM, Nakaya HI, Baron MA, Vilar-Pereira G, Pereira IR, et al. MicroRNA Transcriptome Profiling in Heart of Trypanosoma cruzi-Infected Mice: Parasitological and Cardiological Outcomes. PLoS Negl Trop Dis. 2015;9(6):e0003828.

56. Roman-Campos D, Sales-Junior P, Duarte HL, Gomes ER, Guatimosim S, Ropert C, et al. Cardiomyocyte dysfunction during the chronic phase of Chagas disease. Mem Inst Oswaldo Cruz. 2013;108(2):243–5.

57. Fernandez-Velasco M, Ruiz-Hurtado G, Hurtado O, Moro MA, and Delgado C. TNF-alpha downregulates transient outward potassium current in rat ventricular myocytes through iNOS overexpression and oxidant species generation. Am J Physiol Heart Circ Physiol. 2007;293(1):H238–45.

58. Lewis RS. Calcium signaling mechanisms in T lymphocytes. Annu Rev Immunol. 2001;19:497–521.

59. Hogan PG. Calcium-NFAT transcriptional signalling in T cell activation and T cell exhaustion. Cell Calcium. 2017;63:66–9.

60. Beeton C, Wulff H, Barbaria J, Clot-Faybesse O, Pennington M, Bernard D, et al. Selective blockade of T lymphocyte K(+) channels ameliorates experimental autoimmune encephalomyelitis, a model for multiple sclerosis. Proc Natl Acad Sci U S A. 2001;98(24):13942–7.

61. Beeton C, Wulff H, Standifer NE, Azam P, Mullen KM, Pennington MW, et al. Kv1.3 channels are a therapeutic target for T cell-mediated autoimmune diseases. Proc Natl Acad Sci U S A. 2006;103(46):17414–9.

62. Rus H, Pardo CA, Hu L, Darrah E, Cudrici C, Niculescu T, et al. The voltage-gated potassium channel Kv1.3 is highly expressed on inflammatory infiltrates in multiple sclerosis brain. Proc Natl Acad Sci U S A. 2005;102(31):11094–9.

63. She G, Hou MC, Zhang Y, Zhang Y, Wang Y, Wang HF, et al. Gal-3 (Galectin-3) and KCa3.1 Mediate Heterogeneous Cell Coupling and Myocardial Fibrogenesis Driven by betaAR (beta-Adrenoceptor) Activation. Hypertension. 2020;75(2):393–404.

64. Koberle F. The causation and importance of nervous lesions in American trypanosomiasis. Bull World Health Organ. 1970;42(5):739–43.

65. Villanueva MS. Trypanosomiasis of the central nervous system. Semin Neurol. 1993;13(2):209–18.

66. Amorim DS, and Olsen EG. Assessment of heart neurons in dilated (congestive) cardiomyopathy. Br Heart J. 1982;47(1):11–8.

67. de Souza MM, Andrade SG, Barbosa AA, Jr., Macedo Santos RT, Alves VA, and Andrade ZA. Trypanosoma cruzi strains and autonomic nervous system pathology in experimental Chagas disease. Mem Inst Oswaldo Cruz. 1996;91(2):217–24.

68. Machado CR, Caliari MV, de Lana M, and Tafuri WL. Heart autonomic innervation during the acute phase of experimental American trypanosomiasis in the dog. Am J Trop Med Hyg. 1998;59(3):492–6.

69. Machado CR, and Ribeiro AL. Experimental American trypanomiasis in rats: sympathetic denervation, parasitism and inflammatory process. Mem Inst Oswaldo Cruz. 1989;84(4):549–56.

70. Machado MP, Rocha AM, de Oliveira LF, de Cuba MB, de Oliveira Loss I, Castellano LR, et al. Autonomic nervous system modulation affects the inflammatory immune response in mice with acute Chagas disease. Exp Physiol. 2012;97(11):1186–202.

71. de Cuba MB, Machado MP, Farnesi TS, Alves AC, Martins LA, de Oliveira LF, et al. Effects of cholinergic stimulation with pyridostigmine bromide on chronic chagasic cardiomyopathic mice. Mediators Inflamm. 2014;2014:475946.

72. Elenkov IJ, Wilder RL, Chrousos GP, and Vizi ES. The sympathetic nerve--an integrative interface between two supersystems: the brain and the immune system. Pharmacol Rev. 2000;52(4):595–638.

73. Kin NW, and Sanders VM. It takes nerve to tell T and B cells what to do. J Leukoc Biol. 2006;79(6):1093–104.

74. Kohm AP, and Sanders VM. Norepinephrine and beta 2-adrenergic receptor stimulation regulate CD4+ T and B lymphocyte function in vitro and in vivo. Pharmacol Rev. 2001;53(4):487–525.

75. Sternberg EM. Neural regulation of innate immunity: a coordinated nonspecific host response to pathogens. Nat Rev Immunol. 2006;6(4):318–28.

76. Hobuss L, Bar C, and Thum T. Long Non-coding RNAs: At the Heart of Cardiac Dysfunction? Front Physiol. 2019;10:30.

77. Ballinas-Verdugo MA, Jimenez-Ortega RF, Martinez-Martinez E, Rivas N, Contreras-Lopez EA, Carbo R, et al. Circulating miR-146a as a possible candidate biomarker in the indeterminate phase of Chagas disease. Biol Res. 2021;54(1):21.

78. Ferreira LRP. MicroRNA Transcriptome Profiling in Heart of Trypanosoma cruzi-Infected Mice. Methods Mol Biol. 2019;1955:203–14.

79. Ferreira LRP, Ferreira FM, Laugier L, Cabantous S, Navarro IC, da Silva Candido D, et al. Integration of miRNA and gene expression profiles suggest a role for miRNAs in the pathobiological processes of acute Trypanosoma cruzi infection. Sci Rep. 2017;7(1):17990.

80. Ferreira RR, Abreu RDS, Vilar-Pereira G, Degrave W, Meuser-Batista M, Ferreira NVC, et al. TGF-beta inhibitor therapy decreases fibrosis and stimulates cardiac improvement in a pre-clinical study of chronic Chagas’ heart disease. PLoS Negl Trop Dis. 2019;13(7):e0007602.

81. Jha BK, Varikuti S, Seidler GR, Volpedo G, Satoskar AR, and McGwire BS. MicroRNA-155 Deficiency Exacerbates Trypanosoma cruzi Infection. Infect Immun. 2020;88(7).

82. Nonaka CKV, Macedo CT, Cavalcante BRR, Alcantara AC, Silva DN, Bezerra MDR, et al. Circulating miRNAs as Potential Biomarkers Associated with Cardiac Remodeling and Fibrosis in Chagas Disease Cardiomyopathy. Int J Mol Sci. 2019;20(16).

83. Nonaka CKV, Sampaio GL, Silva KN, Khouri R, Macedo CT, Chagas Translational Research C, et al. Therapeutic miR-21 Silencing Reduces Cardiac Fibrosis and Modulates Inflammatory Response in Chronic Chagas Disease. Int J Mol Sci. 2021;22(7).

84. O’Connell RM, Taganov KD, Boldin MP, Cheng G, and Baltimore D. MicroRNA-155 is induced during the macrophage inflammatory response. Proc Natl Acad Sci U S A. 2007;104(5):1604–9.

85. Corsten MF, Papageorgiou A, Verhesen W, Carai P, Lindow M, Obad S, et al. MicroRNA profiling identifies microRNA-155 as an adverse mediator of cardiac injury and dysfunction during acute viral myocarditis. Circ Res. 2012;111(4):415–25.

86. Hullinger TG, Montgomery RL, Seto AG, Dickinson BA, Semus HM, Lynch JM, et al. Inhibition of miR-15 protects against cardiac ischemic injury. Circ Res. 2012;110(1):71–81.

87. Ye Y, Hu Z, Lin Y, Zhang C, and Perez-Polo JR. Downregulation of microRNA-29 by antisense inhibitors and a PPAR-gamma agonist protects against myocardial ischaemia-reperfusion injury. Cardiovasc Res. 2010;87(3):535–44.

88. Sun C, Huang L, Li Z, Leng K, Xu Y, Jiang X, et al. Long non-coding RNA MIAT in development and disease: a new player in an old game. J Biomed Sci. 2018;25(1):23.

89. Vogel B, Keller A, Frese KS, Leidinger P, Sedaghat-Hamedani F, Kayvanpour E, et al. Multivariate miRNA signatures as biomarkers for non-ischaemic systolic heart failure. Eur Heart J. 2013;34(36):2812–22.

90. Foulquier S, Daskalopoulos EP, Lluri G, Hermans KCM, Deb A, and Blankesteijn WM. WNT Signaling in Cardiac and Vascular Disease. Pharmacol Rev. 2018;70(1):68–141.

91. Chilosi M, Poletti V, Zamo A, Lestani M, Montagna L, Piccoli P, et al. Aberrant Wnt/beta-catenin pathway activation in idiopathic pulmonary fibrosis. Am J Pathol. 2003;162(5):1495–502.

92. Henderson WR, Jr., Chi EY, Ye X, Nguyen C, Tien YT, Zhou B, et al. Inhibition of Wnt/beta-catenin/CREB binding protein (CBP) signaling reverses pulmonary fibrosis. Proc Natl Acad Sci U S A. 2010;107(32):14309–14.

93. Konigshoff M, Balsara N, Pfaff EM, Kramer M, Chrobak I, Seeger W, et al. Functional Wnt signaling is increased in idiopathic pulmonary fibrosis. PLoS One. 2008;3(5):e2142.

94. Akhmetshina A, Palumbo K, Dees C, Bergmann C, Venalis P, Zerr P, et al. Activation of canonical Wnt signalling is required for TGF-beta-mediated fibrosis. Nat Commun. 2012;3:735.

95. Piersma B, Bank RA, and Boersema M. Signaling in Fibrosis: TGF-beta, WNT, and YAP/TAZ Converge. Front Med (Lausanne). 2015;2:59.

96. Vallee A, Lecarpentier Y, Guillevin R, and Vallee JN. Interactions between TGF-beta1, canonical WNT/beta-catenin pathway and PPAR gamma in radiation-induced fibrosis. Oncotarget. 2017;8(52):90579–604.

97. Ferreira RR, de Souza EM, de Oliveira FL, Ferrao PM, Gomes LH, Mendonca-Lima L, et al. Proteins involved on TGF-beta pathway are up-regulated during the acute phase of experimental Chagas disease. Immunobiology. 2016;221(5):587–94.

98. Araujo-Jorge TC, Waghabi MC, Bailly S, and Feige JJ. The TGF-beta pathway as an emerging target for Chagas disease therapy. Clin Pharmacol Ther. 2012;92(5):613–21.

99. Wang Y, Shen RW, Han B, Li Z, Xiong L, Zhang FY, et al. Notch signaling mediated by TGF-beta/Smad pathway in concanavalin A-induced liver fibrosis in rats. World J Gastroenterol. 2017;23(13):2330–6.

100. Pelullo M, Zema S, Nardozza F, Checquolo S, Screpanti I, and Bellavia D. Wnt, Notch, and TGF-beta Pathways Impinge on Hedgehog Signaling Complexity: An Open Window on Cancer. Front Genet. 2019;10:711.

101. Jathar S, Kumar V, Srivastava J, and Tripathi V. Technological Developments in lncRNA Biology. Adv Exp Med Biol. 2017;1008:283–323.

102. Prata A. Clinical and epidemiological aspects of Chagas disease. Lancet Infect Dis. 2001;1(2):92–100.

103. Gomes YM, Lorena VM, and Luquetti AO. Diagnosis of Chagas disease: what has been achieved? What remains to be done with regard to diagnosis and follow up studies? Mem Inst Oswaldo Cruz. 2009;104 Suppl 1:115–21.

104. Balouz V, Aguero F, and Buscaglia CA. Chagas Disease Diagnostic Applications: Present Knowledge and Future Steps. Adv Parasitol. 2017;97:1–45.

105. Ramirez JC, Cura CI, da Cruz Moreira O, Lages-Silva E, Juiz N, Velazquez E, et al. Analytical Validation of Quantitative Real-Time PCR Methods for Quantification of Trypanosoma cruzi DNA in Blood Samples from Chagas Disease Patients. J Mol Diagn. 2015;17(5):605–15.

106. Keating SM, Deng X, Fernandes F, Cunha-Neto E, Ribeiro AL, Adesina B, et al. Inflammatory and cardiac biomarkers are differentially expressed in clinical stages of Chagas disease. Int J Cardiol. 2015;199:451–9.

107. Panagopoulou V, Deftereos S, Kossyvakis C, Raisakis K, Giannopoulos G, Bouras G, et al. NTproBNP: an important biomarker in cardiac diseases. Curr Top Med Chem. 2013;13(2):82–94.

108. Porras AI, Yadon ZE, Altcheh J, Britto C, Chaves GC, Flevaud L, et al. Target Product Profile (TPP) for Chagas Disease Point-of-Care Diagnosis and Assessment of Response to Treatment. PLoS Negl Trop Dis. 2015;9(6):e0003697.

