## Supplementary material for "Specific methylation marks in promoter regions are associated to the pathogenic process of Chronic Chagas disease Cardiomyopathy by modifying transcription factor binding patterns": Supplementary table 1.docx

**Supplementary table 1.** Cell lines description.

| **Cell line** | **Description** |
| --- | --- |
| CHRF28811 | Acute megakaryoblastic leukemia |
| AMLBLAST | Acute Myeloid Leukemia |
| AMLPZ12 | Acute Myeloid Leukemia |
| TSU1621MT | AML cell lines |
| AC16 | Cardiomyocyte |
| BCELL | B lymphocyte |
| CA46 | B lymphocyte |
| NAMALWA | B lymphocyte |
| P4936 | B lymphocyte |
| RAJI | B lymphocyte |
| RAMOS | B lymphocyte |
| RCK8 | B lymphocyte |
| FARAGE | B lymphocyte |
| GM06990 | B lymphocyte |
| GM10248 | B lymphocyte |
| GM10266 | B lymphocyte |
| GM10847 | B lymphocyte |
| GM12801 | B lymphocyte |
| GM12864 | B lymphocyte |
| GM12865 | B lymphocyte |
| GM12866 | B lymphocyte |
| GM12867 | B lymphocyte |
| GM12868 | B lymphocyte |
| GM12869 | B lymphocyte |
| GM12870 | B lymphocyte |
| GM12872 | B lymphocyte |
| GM12873 | B lymphocyte |
| GM12874 | B lymphocyte |
| GM12875 | B lymphocyte |
| GM12878 | B lymphocyte |
| GM12891 | B lymphocyte |
| GM12892 | B lymphocyte |
| GM13976 | B lymphocyte |
| GM13977 | B lymphocyte |
| GM15510 | B lymphocyte |
| GM18505 | B lymphocyte |
| GM18526 | B lymphocyte |
| GM18951 | B lymphocyte |
| GM19099 | B lymphocyte |
| GM19193 | B lymphocyte |
| GM19238 | B lymphocyte |
| GM19239 | B lymphocyte |
| GM19240 | B lymphocyte |
| GM20000 | B lymphocyte |
| LYMPHOBLASTOID | B lymphocyte |
| MUTUL | B lymphocyte |
| U2932 | B lymphocyte |
| LP1 | B lymphocyte |
| ME1 | B lymphocyte |
| SUDHL10 | B lymphocyte |
| SUDHL2 | B lymphocyte |
| SUDHL4 | B lymphocyte |
| SUDHL5 | B lymphocyte |
| SUDHL6 | B lymphocyte |
| DOHH2 | B lymphocyte |
| GRANTA519 | B lymphocyte |
| HBL1 | B lymphocyte |
| KARPAS422 | B lymphocyte |
| MM1S | B lymphocyte |
| OCILY10 | B lymphocyte |
| OCILY19 | B lymphocyte |
| OCILY1 | B lymphocyte |
| OCILY3 | B lymphocyte |
| OCILY7 | B lymphocyte |
| OE33 | B lymphocyte |
| PFEIFFER | B lymphocyte |
| WSUDLCL2 | B lymphocyte |
| BC3 | B lymphocyte |
| BCBL1 | B lymphocyte |
| CD4 | T lymphocyte |
| CD8 | T lymphocyte |
| DND41 | T lymphocyte |
| CUTLL1 | T lymphocyte |
| RPMI8402 | T lymphocyte |
| CCRFCEM | T lymphocyte |
| PRIMA2 | T lymphocyte |
| PRIMA5 | T lymphocyte |
| JURKAT | T lymphocyte |
| HPBALL | T lymphocyte |
| NALM6 | Lymphocyte-like (myeloid leukemia) |
| MACROPHAGE | Macrophage |
| MV411 | Macrophage |
| KOPTK1 | T lymphocyte |
| LOUCY | T lymphocyte |
