## Supplementary material for "Specific methylation marks in promoter regions are associated to the pathogenic process of Chronic Chagas disease Cardiomyopathy by modifying transcription factor binding patterns": Supplementary table 2.docx

**Supplementary table 2.** Statistics on coding sequences detected by RNA-seq and on differentially expressed genes.

| Gene type | Nb. All genes | % (All genes) | Nb. DEGs | % (DEG) | pvalue | Corrected pvalue |
| --- | --- | --- | --- | --- | --- | --- |
| protein_coding | 18899 | 41.94 | 893 | 63.38 | 2.10E-21 | 3.36E-20 |
| pseudogene | 9565 | 21.22 | 155 | 11 | 6.20E-15 | 4.96E-14 |
| lincRNA | 5312 | 11.79 | 146 | 10.36 | 1.60E-01 | 2.56E-01 |
| antisense | 4499 | 9.98 | 132 | 9.37 | 5.20E-01 | 7.56E-01 |
| miRNA | 1705 | 3.78 | 19 | 1.35 | 4.90E-06 | 1.96E-05 |
| misc_RNA | 1455 | 3.23 | 8 | 0.57 | 5.10E-08 | 2.72E-07 |
| snRNA | 1150 | 2.55 | 9 | 0.64 | 1.20E-05 | 3.84E-05 |
| snoRNA | 922 | 2.05 | 7 | 0.5 | 8.10E-05 | 2.16E-04 |
| sense_intronic | 678 | 1.5 | 14 | 0.99 | 1.50E-01 | 2.56E-01 |
| processed_transcript | 404 | 0.9 | 14 | 0.99 | 8.20E-01 | 9.83E-01 |
| rRNA | 235 | 0.52 | 1 | 0.07 | 3.20E-02 | 6.40E-02 |
| sense_overlapping | 175 | 0.39 | 5 | 0.35 | 1.00E+00 | 1.00E+00 |
| polymorphic_pseudogene | 31 | 0.07 | 5 | 0.35 | 9.40E-04 | 2.15E-03 |
| Mt_tRNA | 22 | 0.05 | 0 | 0 | 8.40E-01 | 9.83E-01 |
| 3prime_overlapping_ncrna | 12 | 0.03 | 1 | 0.07 | 8.60E-01 | 9.83E-01 |
| Mt_rRNA | 2 | 0 | 0 | 0 | 1.00E+00 | 1.00E+00 |
