## Supplementary material for "Specific methylation marks in promoter regions are associated to the pathogenic process of Chronic Chagas disease Cardiomyopathy by modifying transcription factor binding patterns": Supplementary table 3.docx

**Supplementary table 3.** List of unique differentially expressed genes identified by RNA-seq between 8 end stage CCC heart tissue samples and 6 heart tissue samples obtained from organ donors.

| **Gene id** | **Gene Name** | **Fold Change** | **Log2 Fold Change** | **pvalue** | **Corrected pvalue** |
| --- | --- | --- | --- | --- | --- |
| ENSG00000270641 | TSIX | 3114.32 | 11.60 | 1.37E-08 | 8.49E-07 |
| ENSG00000226777 | KIAA0125 | 1553.24 | 10.60 | 1.25E-20 | 7.15E-18 |
| ENSG00000116748 | AMPD1 | 1203.96 | 10.23 | 1.76E-18 | 6.78E-16 |
| ENSG00000254709 | IGLL5 | 1078.2 | 10.07 | 3.85E-19 | 1.72E-16 |
| ENSG00000121895 | TMEM156 | 999.84 | 9.97 | 5.03E-20 | 2.58E-17 |
| ENSG00000231486 | AC096579.7 | 740.88 | 9.53 | 4.43E-17 | 1.30E-14 |
| ENSG00000136573 | BLK | 702.49 | 9.46 | 1.04E-14 | 2.05E-12 |
| ENSG00000109684 | CLNK | 502.16 | 8.97 | 2.37E-20 | 1.30E-17 |
| ENSG00000169435 | RASSF6 | 434.13 | 8.76 | 3.70E-13 | 5.97E-11 |
| ENSG00000172752 | COL6A5 | 400.34 | 8.65 | 4.63E-07 | 1.89E-05 |
| ENSG00000229807 | XIST | 290.75 | 8.18 | 4.31E-12 | 5.55E-10 |
| ENSG00000176083 | ZNF683 | 275.72 | 8.11 | 6.27E-18 | 2.22E-15 |
| ENSG00000246084 | CTD-2506J14.1 | 274.64 | 8.10 | 5.67E-11 | 5.88E-09 |
| ENSG00000158485 | CD1B | 263.86 | 8.04 | 3.82E-09 | 2.79E-07 |
| ENSG00000143297 | FCRL5 | 262.06 | 8.03 | 6.43E-33 | 1.98E-29 |
| ENSG00000205846 | CLEC6A | 202.7 | 7.66 | 8.62E-08 | 4.44E-06 |
| ENSG00000056291 | NPFFR2 | 198.83 | 7.64 | 2.41E-10 | 2.23E-08 |
| ENSG00000188822 | CNR2 | 180.78 | 7.50 | 7.10E-08 | 3.72E-06 |
| ENSG00000128438 | TBC1D27 | 165.49 | 7.37 | 2.79E-09 | 2.08E-07 |
| ENSG00000117215 | PLA2G2D | 164.89 | 7.37 | 1.29E-10 | 1.27E-08 |
| ENSG00000249318 | AC010468.2 | 162.43 | 7.34 | 2.30E-08 | 1.38E-06 |
| ENSG00000132465 | IGJ | 147.99 | 7.21 | 3.32E-34 | 1.46E-30 |
| ENSG00000251408 | RP11-586D19.2 | 142.08 | 7.15 | 4.26E-08 | 2.35E-06 |
| ENSG00000235621 | LINC00494 | 140.84 | 7.14 | 7.54E-09 | 4.99E-07 |
| ENSG00000255183 | RP11-720D4.3 | 138.72 | 7.12 | 1.62E-07 | 7.63E-06 |
| ENSG00000170819 | BFSP2 | 128.96 | 7.01 | 1.64E-08 | 1.00E-06 |
| ENSG00000256039 | RP11-291B21.2 | 128.08 | 7.00 | 4.07E-32 | 1.14E-28 |
| ENSG00000183625 | CCR3 | 126.08 | 6.98 | 1.15E-06 | 4.19E-05 |
| ENSG00000259731 | RP11-326N17.1 | 123.96 | 6.95 | 9.03E-08 | 4.60E-06 |
| ENSG00000091181 | IL5RA | 113.3 | 6.82 | 1.35E-13 | 2.29E-11 |
| ENSG00000196092 | PAX5 | 110.2 | 6.78 | 1.13E-09 | 9.23E-08 |
| ENSG00000204110 | RP1-153P14.8 | 109.34 | 6.77 | 7.58E-07 | 2.94E-05 |
| ENSG00000116147 | TNR | 108.12 | 6.76 | 4.48E-07 | 1.85E-05 |
| ENSG00000188282 | RUFY4 | 95.76 | 6.58 | 1.34E-10 | 1.30E-08 |
| ENSG00000258379 | RP11-204N11.2 | 95.33 | 6.57 | 6.11E-07 | 2.44E-05 |
| ENSG00000039068 | CDH1 | 93.15 | 6.54 | 5.85E-07 | 2.35E-05 |
| ENSG00000105369 | CD79A | 91.13 | 6.51 | 2.42E-26 | 3.11E-23 |
| ENSG00000240754 | RP11-38J22.6 | 88.59 | 6.47 | 6.85E-07 | 2.70E-05 |
| ENSG00000189233 | NUGGC | 86.06 | 6.43 | 3.42E-36 | 2.11E-32 |
| ENSG00000134028 | ADAMDEC1 | 84.41 | 6.40 | 2.19E-07 | 9.82E-06 |
| ENSG00000160882 | CYP11B1 | 83.33 | 6.38 | 6.23E-06 | 0.000179712 |
| ENSG00000249201 | CTD-3080P12.3 | 79.24 | 6.31 | 2.10E-07 | 9.45E-06 |
| ENSG00000138755 | CXCL9 | 78.46 | 6.29 | 3.56E-20 | 1.89E-17 |
| ENSG00000135443 | KRT85 | 76.5 | 6.26 | 8.14E-07 | 3.13E-05 |
| ENSG00000266088 | RP5-1028K7.2 | 70.38 | 6.14 | 3.52E-05 | 0.000743609 |
| ENSG00000256155 | RP11-277P12.9 | 68.27 | 6.09 | 6.78E-07 | 2.68E-05 |
| ENSG00000226025 | LGALS17A | 67.79 | 6.08 | 3.24E-08 | 1.85E-06 |
| ENSG00000146285 | SCML4 | 66.79 | 6.06 | 5.47E-34 | 2.11E-30 |
| ENSG00000230943 | RP11-367G18.1 | 63.31 | 5.98 | 1.09E-05 | 0.000286527 |
| ENSG00000227920 | RP1-153P14.5 | 62.89 | 5.97 | 3.94E-05 | 0.000813993 |
| ENSG00000180537 | RNF182 | 59.77 | 5.90 | 2.27E-05 | 0.000516203 |
| ENSG00000255833 | TIFAB | 59.74 | 5.90 | 5.10E-12 | 6.52E-10 |
| ENSG00000259343 | RP11-761I4.3 | 59.6 | 5.90 | 8.68E-06 | 0.000236605 |
| ENSG00000186265 | BTLA | 59.07 | 5.88 | 1.59E-17 | 5.01E-15 |
| ENSG00000169213 | RAB3B | 58.98 | 5.88 | 1.54E-09 | 1.21E-07 |
| ENSG00000265714 | AL122127.3 | 58.13 | 5.86 | 1.58E-05 | 0.000384478 |
| ENSG00000230146 | SEPHS1P4 | 57.28 | 5.84 | 8.00E-06 | 0.000221481 |
| ENSG00000249993 | BFSP2-AS1 | 56.19 | 5.81 | 3.12E-05 | 0.000672129 |
| ENSG00000122787 | AKR1D1 | 54.89 | 5.78 | 0.00012906 | 0.002167004 |
| ENSG00000258512 | RP11-796G6.2 | 54.69 | 5.77 | 8.06E-06 | 0.000222699 |
| ENSG00000254650 | RP11-665E10.5 | 54.58 | 5.77 | 3.90E-05 | 0.000807254 |
| ENSG00000230006 | ANKRD36BP2 | 53.3 | 5.74 | 3.98E-35 | 2.04E-31 |
| ENSG00000225720 | RP4-742C19.12 | 53.13 | 5.73 | 5.74E-05 | 0.001120525 |
| ENSG00000267178 | PHF5CP | 52.82 | 5.72 | 0.000167082 | 0.002649496 |
| ENSG00000174453 | VWC2L | 52.73 | 5.72 | 3.39E-06 | 0.000107205 |
| ENSG00000112812 | PRSS16 | 51.71 | 5.69 | 2.46E-05 | 0.000551218 |
| ENSG00000115884 | SDC1 | 50.73 | 5.66 | 9.40E-39 | 2.90E-34 |
| ENSG00000110777 | POU2AF1 | 50.6 | 5.66 | 1.23E-29 | 2.36E-26 |
| ENSG00000264781 | hsa-mir-4537 | 49.43 | 5.63 | 1.56E-05 | 0.000381946 |
| ENSG00000153789 | FAM92B | 48.96 | 5.61 | 0.000267006 | 0.003851456 |
| ENSG00000205592 | MUC19 | 48.7 | 5.61 | 4.88E-05 | 0.000973388 |
| ENSG00000105717 | PBX4 | 47.69 | 5.58 | 9.13E-15 | 1.83E-12 |
| ENSG00000238121 | LINC00426 | 47.36 | 5.57 | 3.65E-27 | 5.11E-24 |
| ENSG00000244618 | RN7SL334P | 47.34 | 5.56 | 0.000131665 | 0.002197579 |
| ENSG00000215764 | KIR2DL2 | 47.34 | 5.56 | 0.000278164 | 0.00397583 |
| ENSG00000196684 | HSH2D | 46.34 | 5.53 | 2.06E-27 | 3.35E-24 |
| ENSG00000228168 | HNRNPA1P21 | 46.18 | 5.53 | 0.000129476 | 0.00216996 |
| ENSG00000122043 | LINC00544 | 42.94 | 5.42 | 6.61E-05 | 0.00125217 |
| ENSG00000234663 | AC104820.2 | 42.82 | 5.42 | 2.10E-18 | 7.87E-16 |
| ENSG00000244649 | CTD-2377D24.6 | 42.76 | 5.42 | 3.28E-05 | 0.000702602 |
| ENSG00000235366 | LINC01055 | 42.17 | 5.40 | 0.00011857 | 0.002018384 |
| ENSG00000233093 | LINC00892 | 41.97 | 5.39 | 2.29E-15 | 5.15E-13 |
| ENSG00000241577 | RP11-523O18.7 | 41.16 | 5.36 | 6.15E-05 | 0.001181739 |
| ENSG00000255354 | RP11-148O21.2 | 41.07 | 5.36 | 8.99E-05 | 0.001610778 |
| ENSG00000166105 | GLB1L3 | 39.5 | 5.30 | 1.76E-07 | 8.14E-06 |
| ENSG00000253980 | CTB-4E7.1 | 39.21 | 5.29 | 3.66E-07 | 1.55E-05 |
| ENSG00000166211 | SPIC | 38.67 | 5.27 | 0.000315609 | 0.004396129 |
| ENSG00000260719 | AC009133.17 | 38.56 | 5.27 | 4.73E-08 | 2.58E-06 |
| ENSG00000244432 | RPL39P28 | 38.53 | 5.27 | 0.000166031 | 0.002634398 |
| ENSG00000127318 | IL22 | 38.14 | 5.25 | 0.000131266 | 0.002193293 |
| ENSG00000220517 | ASS1P1 | 36.8 | 5.20 | 9.85E-11 | 9.86E-09 |
| ENSG00000204475 | NCR3 | 36.79 | 5.20 | 1.31E-10 | 1.27E-08 |
| ENSG00000150556 | LYPD6B | 36.65 | 5.20 | 0.000206933 | 0.003128688 |
| ENSG00000104970 | KIR3DX1 | 36.55 | 5.19 | 2.24E-07 | 1.00E-05 |
| ENSG00000226659 | RP11-137H2.4 | 36.25 | 5.18 | 9.19E-05 | 0.001639493 |
| ENSG00000204969 | PCDHA2 | 36.23 | 5.18 | 0.00016923 | 0.002679413 |
| ENSG00000203416 | FAM32B | 36.17 | 5.18 | 0.000140092 | 0.002314412 |
| ENSG00000225079 | FTH1P22 | 35.88 | 5.16 | 3.93E-10 | 3.52E-08 |
| ENSG00000170476 | MZB1 | 35.8 | 5.16 | 1.10E-37 | 8.50E-34 |
| ENSG00000143185 | XCL2 | 35.48 | 5.15 | 5.09E-10 | 4.47E-08 |
| ENSG00000261786 | RP4-555D20.2 | 34.65 | 5.11 | 0.000956987 | 0.010144149 |
| ENSG00000152766 | ANKRD22 | 34.58 | 5.11 | 4.19E-09 | 3.05E-07 |
| ENSG00000156738 | MS4A1 | 34.28 | 5.10 | 6.86E-15 | 1.40E-12 |
| ENSG00000132704 | FCRL2 | 34.25 | 5.10 | 5.62E-09 | 3.88E-07 |
| ENSG00000162739 | SLAMF6 | 33.7 | 5.07 | 3.53E-12 | 4.67E-10 |
| ENSG00000168671 | UGT3A2 | 33.24 | 5.05 | 0.00071087 | 0.008110876 |
| ENSG00000237301 | RP4-680D5.2 | 32.89 | 5.04 | 0.000131046 | 0.0021908 |
| ENSG00000141293 | SKAP1 | 32.61 | 5.03 | 1.32E-17 | 4.28E-15 |
| ENSG00000152969 | JAKMIP1 | 32.4 | 5.02 | 3.07E-11 | 3.35E-09 |
| ENSG00000243772 | KIR2DL3 | 32.37 | 5.02 | 0.000771725 | 0.008608838 |
| ENSG00000168421 | RHOH | 32.36 | 5.02 | 6.60E-26 | 7.82E-23 |
| ENSG00000235576 | AC092580.4 | 32.15 | 5.01 | 7.04E-13 | 1.05E-10 |
| ENSG00000223511 | RP13-297E16.4 | 32.03 | 5.00 | 6.30E-07 | 2.50E-05 |
| ENSG00000205809 | KLRC2 | 31.88 | 4.99 | 8.57E-13 | 1.25E-10 |
| ENSG00000013725 | CD6 | 31.83 | 4.99 | 1.99E-15 | 4.50E-13 |
| ENSG00000258869 | RP11-204N11.1 | 31.72 | 4.99 | 4.19E-12 | 5.42E-10 |
| ENSG00000181036 | FCRL6 | 31.38 | 4.97 | 2.29E-38 | 3.52E-34 |
| ENSG00000167618 | LAIR2 | 31.07 | 4.96 | 0.001053032 | 0.010905871 |
| ENSG00000181143 | MUC16 | 31.03 | 4.96 | 5.39E-12 | 6.84E-10 |
| ENSG00000237286 | AC004906.3 | 30.87 | 4.95 | 0.000557397 | 0.006758735 |
| ENSG00000100450 | GZMH | 30.04 | 4.91 | 1.19E-09 | 9.63E-08 |
| ENSG00000026751 | SLAMF7 | 29.66 | 4.89 | 1.70E-14 | 3.20E-12 |
| ENSG00000078081 | LAMP3 | 29.47 | 4.88 | 1.45E-07 | 6.95E-06 |
| ENSG00000251309 | RP11-498M5.2 | 29.04 | 4.86 | 0.000389148 | 0.005173796 |
| ENSG00000173578 | XCR1 | 28.94 | 4.86 | 2.72E-11 | 3.00E-09 |
| ENSG00000226667 | RP11-292F22.5 | 28.49 | 4.83 | 0.000412356 | 0.005394953 |
| ENSG00000122188 | LAX1 | 28.33 | 4.82 | 1.06E-24 | 1.02E-21 |
| ENSG00000255391 | CTD-2028E8.1 | 28.23 | 4.82 | 4.62E-07 | 1.89E-05 |
| ENSG00000153283 | CD96 | 27.93 | 4.80 | 6.76E-14 | 1.20E-11 |
| ENSG00000172673 | THEMIS | 27.21 | 4.77 | 1.06E-20 | 6.24E-18 |
| ENSG00000222179 | RN7SKP26 | 27.1 | 4.76 | 0.000689379 | 0.007946298 |
| ENSG00000177494 | ZBED2 | 26.91 | 4.75 | 5.35E-08 | 2.86E-06 |
| ENSG00000113263 | ITK | 26.32 | 4.72 | 3.66E-30 | 8.68E-27 |
| ENSG00000060140 | STYK1 | 26.3 | 4.72 | 7.88E-17 | 2.21E-14 |
| ENSG00000176320 | RP11-404O13.5 | 26.24 | 4.71 | 3.45E-07 | 1.47E-05 |
| ENSG00000248969 | CTD-2113L7.1 | 26.22 | 4.71 | 0.000478709 | 0.006054805 |
| ENSG00000127152 | BCL11B | 25.99 | 4.70 | 2.26E-25 | 2.32E-22 |
| ENSG00000259278 | RP11-62C7.2 | 25.86 | 4.69 | 1.20E-07 | 5.89E-06 |
| ENSG00000254965 | RP11-113K21.2 | 25.8 | 4.69 | 0.00043755 | 0.005633661 |
| ENSG00000137078 | SIT1 | 25.66 | 4.68 | 7.32E-16 | 1.75E-13 |
| ENSG00000122224 | LY9 | 25.58 | 4.68 | 9.44E-26 | 1.04E-22 |
| ENSG00000100298 | APOBEC3H | 25.16 | 4.65 | 2.11E-11 | 2.42E-09 |
| ENSG00000224523 | RP13-52K8.1 | 24.8 | 4.63 | 0.000600992 | 0.007138511 |
| ENSG00000176769 | TCERG1L | 24.77 | 4.63 | 0.000652035 | 0.007604034 |
| ENSG00000272763 | RP11-357H14.17 | 24.74 | 4.63 | 0.000873693 | 0.009458666 |
| ENSG00000264198 | RP11-94L15.2 | 24.69 | 4.63 | 6.09E-23 | 4.81E-20 |
| ENSG00000172724 | CCL19 | 24.53 | 4.62 | 1.90E-10 | 1.79E-08 |
| ENSG00000231764 | DLX6-AS1 | 24.27 | 4.60 | 0.001349515 | 0.013172503 |
| ENSG00000124203 | ZNF831 | 24.17 | 4.60 | 1.06E-22 | 8.19E-20 |
| ENSG00000168685 | IL7R | 23.78 | 4.57 | 7.81E-16 | 1.82E-13 |
| ENSG00000244720 | RP11-402J7.2 | 23.72 | 4.57 | 3.97E-11 | 4.20E-09 |
| ENSG00000162676 | GFI1 | 23.68 | 4.57 | 8.03E-22 | 5.50E-19 |
| ENSG00000167077 | MEI1 | 23.42 | 4.55 | 6.81E-30 | 1.50E-26 |
| ENSG00000226003 | RP11-312J18.3 | 22.98 | 4.52 | 0.00083065 | 0.009088484 |
| ENSG00000168081 | PNOC | 22.89 | 4.52 | 5.97E-07 | 2.39E-05 |
| ENSG00000124334 | IL9R | 22.84 | 4.51 | 1.93E-08 | 1.17E-06 |
| ENSG00000188820 | FAM26F | 22.64 | 4.50 | 8.38E-07 | 3.21E-05 |
| ENSG00000188011 | CXXC11 | 22.56 | 4.50 | 9.25E-07 | 3.49E-05 |
| ENSG00000158488 | CD1E | 22.34 | 4.48 | 3.82E-06 | 0.000118407 |
| ENSG00000263787 | RP11-456D7.1 | 22.24 | 4.47 | 2.55E-06 | 8.42E-05 |
| ENSG00000259803 | SLC22A31 | 22.2 | 4.47 | 0.001753107 | 0.016042462 |
| ENSG00000233261 | LINC00264 | 22.16 | 4.47 | 0.001931322 | 0.017347396 |
| ENSG00000161405 | IKZF3 | 22.1 | 4.47 | 2.42E-12 | 3.33E-10 |
| ENSG00000253701 | AL928768.3 | 22.04 | 4.46 | 3.72E-06 | 0.000115698 |
| ENSG00000230539 | AOAH-IT1 | 22.03 | 4.46 | 6.27E-11 | 6.44E-09 |
| ENSG00000216054 | AC019201.1 | 21.77 | 4.44 | 9.08E-06 | 0.000246063 |
| ENSG00000198851 | CD3E | 21.76 | 4.44 | 4.57E-19 | 1.98E-16 |
| ENSG00000167476 | JSRP1 | 21.72 | 4.44 | 2.17E-09 | 1.65E-07 |
| ENSG00000269919 | RP1-134E15.3 | 21.66 | 4.44 | 1.28E-12 | 1.83E-10 |
| ENSG00000161570 | CCL5 | 21.65 | 4.44 | 6.65E-12 | 8.36E-10 |
| ENSG00000180535 | BHLHA15 | 21.22 | 4.41 | 3.97E-10 | 3.55E-08 |
| ENSG00000253364 | RP11-731F5.2 | 21.14 | 4.40 | 0.000907337 | 0.009744946 |
| ENSG00000182183 | FAM159A | 21.09 | 4.40 | 3.77E-12 | 4.97E-10 |
| ENSG00000236481 | AC002331.1 | 21.01 | 4.39 | 0.001375222 | 0.013341296 |
| ENSG00000181215 | C4orf50 | 20.85 | 4.38 | 1.06E-08 | 6.76E-07 |
| ENSG00000224805 | LINC00853 | 20.82 | 4.38 | 0.001483903 | 0.014145808 |
| ENSG00000173239 | LIPM | 20.81 | 4.38 | 8.24E-06 | 0.000226808 |
| ENSG00000144406 | UNC80 | 20.63 | 4.37 | 1.44E-11 | 1.68E-09 |
| ENSG00000189238 | LINC00943 | 20.19 | 4.34 | 1.17E-13 | 2.01E-11 |
| ENSG00000073861 | TBX21 | 20.18 | 4.34 | 2.41E-27 | 3.72E-24 |
| ENSG00000163519 | TRAT1 | 20.15 | 4.33 | 7.40E-17 | 2.11E-14 |
| ENSG00000229754 | CXCR2P1 | 20.08 | 4.33 | 4.37E-08 | 2.40E-06 |
| ENSG00000124256 | ZBP1 | 20.05 | 4.33 | 2.70E-16 | 7.12E-14 |
| ENSG00000227039 | ITGB2-AS1 | 20.04 | 4.33 | 7.97E-19 | 3.27E-16 |
| ENSG00000140284 | SLC27A2 | 19.97 | 4.32 | 8.35E-08 | 4.33E-06 |
| ENSG00000153563 | CD8A | 19.96 | 4.32 | 1.24E-23 | 1.09E-20 |
| ENSG00000213434 | VTI1BP2 | 19.75 | 4.30 | 0.000769585 | 0.008600534 |
| ENSG00000196934 | RIMBP3B | 19.73 | 4.30 | 0.001319156 | 0.012935876 |
| ENSG00000153898 | MCOLN2 | 19.61 | 4.29 | 4.28E-18 | 1.55E-15 |
| ENSG00000227678 | RP11-73O6.3 | 19.46 | 4.28 | 1.20E-05 | 0.000307815 |
| ENSG00000135925 | WNT10A | 19.39 | 4.28 | 8.67E-08 | 4.45E-06 |
| ENSG00000122223 | CD244 | 19.27 | 4.27 | 2.55E-19 | 1.16E-16 |
| ENSG00000183542 | KLRC4 | 19.26 | 4.27 | 2.67E-11 | 2.97E-09 |
| ENSG00000116824 | CD2 | 19.24 | 4.27 | 5.00E-19 | 2.11E-16 |
| ENSG00000267311 | RP11-99A1.2 | 19.13 | 4.26 | 0.001313566 | 0.012893367 |
| ENSG00000187323 | DCC | 18.92 | 4.24 | 1.31E-05 | 0.000330091 |
| ENSG00000005844 | ITGAL | 18.86 | 4.24 | 9.01E-26 | 1.03E-22 |
| ENSG00000172116 | CD8B | 18.78 | 4.23 | 4.67E-11 | 4.87E-09 |
| ENSG00000259307 | PLCB2-AS1 | 18.67 | 4.22 | 0.001167101 | 0.011793882 |
| ENSG00000240787 | RP11-615J4.4 | 18.64 | 4.22 | 1.09E-06 | 4.03E-05 |
| ENSG00000159618 | GPR114 | 18.32 | 4.20 | 4.65E-14 | 8.42E-12 |
| ENSG00000065675 | PRKCQ | 18.22 | 4.19 | 7.27E-25 | 7.23E-22 |
| ENSG00000251215 | GOLGA5P1 | 17.89 | 4.16 | 4.98E-09 | 3.54E-07 |
| ENSG00000224557 | HLA-DPB2 | 17.65 | 4.14 | 1.59E-05 | 0.000387545 |
| ENSG00000238685 | ACA64 | 17.52 | 4.13 | 0.001710574 | 0.015732689 |
| ENSG00000227145 | IL21-AS1 | 17.49 | 4.13 | 2.41E-05 | 0.000542415 |
| ENSG00000102245 | CD40LG | 17.48 | 4.13 | 3.37E-17 | 9.98E-15 |
| ENSG00000164287 | CDC20B | 17.39 | 4.12 | 0.002687135 | 0.022224307 |
| ENSG00000205021 | CCL3L1 | 17.22 | 4.11 | 1.55E-05 | 0.00037835 |
| ENSG00000167286 | CD3D | 17.13 | 4.10 | 3.91E-16 | 9.96E-14 |
| ENSG00000249948 | GBA3 | 17.13 | 4.10 | 3.09E-05 | 0.000665713 |
| ENSG00000263264 | CTB-133G6.1 | 17.09 | 4.09 | 5.53E-15 | 1.14E-12 |
| ENSG00000245164 | LINC00861 | 16.89 | 4.08 | 2.82E-09 | 2.10E-07 |
| ENSG00000232812 | RP11-459K23.2 | 16.83 | 4.07 | 0.002593646 | 0.021633145 |
| ENSG00000213809 | KLRK1 | 16.66 | 4.06 | 1.92E-10 | 1.80E-08 |
| ENSG00000110848 | CD69 | 16.61 | 4.05 | 3.72E-23 | 3.19E-20 |
| ENSG00000134545 | KLRC1 | 16.57 | 4.05 | 2.40E-14 | 4.43E-12 |
| ENSG00000179934 | CCR8 | 16.57 | 4.05 | 0.000265712 | 0.003838181 |
| ENSG00000163599 | CTLA4 | 16.53 | 4.05 | 1.25E-16 | 3.37E-14 |
| ENSG00000169436 | COL22A1 | 16.49 | 4.04 | 6.12E-08 | 3.23E-06 |
| ENSG00000154451 | GBP5 | 16.42 | 4.04 | 5.20E-10 | 4.55E-08 |
| ENSG00000205890 | RP11-473M20.5 | 16.33 | 4.03 | 0.002002204 | 0.017849237 |
| ENSG00000213886 | UBD | 16.29 | 4.03 | 4.82E-05 | 0.000964548 |
| ENSG00000177272 | KCNA3 | 16.19 | 4.02 | 1.57E-17 | 4.99E-15 |
| ENSG00000256128 | LINC00944 | 16.18 | 4.02 | 3.49E-11 | 3.75E-09 |
| ENSG00000125084 | WNT1 | 16.13 | 4.01 | 9.54E-06 | 0.000257587 |
| ENSG00000139193 | CD27 | 16.07 | 4.01 | 1.45E-18 | 5.64E-16 |
| ENSG00000173988 | LRRC63 | 16.03 | 4.00 | 2.04E-06 | 6.99E-05 |
| ENSG00000174123 | TLR10 | 15.94 | 3.99 | 7.99E-08 | 4.15E-06 |
| ENSG00000229228 | LINC00582 | 15.91 | 3.99 | 5.11E-06 | 0.000151128 |
| ENSG00000160654 | CD3G | 15.9 | 3.99 | 1.10E-19 | 5.45E-17 |
| ENSG00000111537 | IFNG | 15.89 | 3.99 | 4.22E-09 | 3.05E-07 |
| ENSG00000186810 | CXCR3 | 15.88 | 3.99 | 1.97E-17 | 6.08E-15 |
| ENSG00000155961 | RAB39B | 15.88 | 3.99 | 2.97E-15 | 6.50E-13 |
| ENSG00000101842 | VSIG1 | 15.82 | 3.98 | 3.39E-05 | 0.000721422 |
| ENSG00000229405 | AC092580.1 | 15.81 | 3.98 | 6.85E-10 | 5.84E-08 |
| ENSG00000177455 | CD19 | 15.79 | 3.98 | 1.07E-07 | 5.30E-06 |
| ENSG00000117560 | FASLG | 15.76 | 3.98 | 6.69E-12 | 8.38E-10 |
| ENSG00000103522 | IL21R | 15.72 | 3.97 | 3.23E-13 | 5.25E-11 |
| ENSG00000262400 | RP11-191A15.1 | 15.7 | 3.97 | 0.002843788 | 0.023250111 |
| ENSG00000160185 | UBASH3A | 15.59 | 3.96 | 6.86E-14 | 1.22E-11 |
| ENSG00000164512 | ANKRD55 | 15.48 | 3.95 | 1.80E-07 | 8.32E-06 |
| ENSG00000034053 | APBA2 | 15.38 | 3.94 | 3.99E-18 | 1.46E-15 |
| ENSG00000167653 | PSCA | 15.38 | 3.94 | 0.002512548 | 0.02112831 |
| ENSG00000129277 | CCL4 | 15.37 | 3.94 | 1.07E-20 | 6.24E-18 |
| ENSG00000227159 | DDX11L16 | 15.32 | 3.94 | 0.002217625 | 0.019236274 |
| ENSG00000262823 | RP13-580F15.2 | 15.28 | 3.93 | 9.96E-06 | 0.000266909 |
| ENSG00000251149 | MTND5P5 | 15.25 | 3.93 | 0.00315174 | 0.025123209 |
| ENSG00000089012 | SIRPG | 15.03 | 3.91 | 4.61E-12 | 5.92E-10 |
| ENSG00000184613 | NELL2 | 15.03 | 3.91 | 7.68E-07 | 2.98E-05 |
| ENSG00000115085 | ZAP70 | 14.94 | 3.90 | 8.77E-29 | 1.59E-25 |
| ENSG00000166523 | CLEC4E | 14.94 | 3.90 | 6.36E-18 | 2.23E-15 |
| ENSG00000243536 | ANTXRLP1 | 14.94 | 3.90 | 5.61E-08 | 2.98E-06 |
| ENSG00000015413 | DPEP1 | 14.94 | 3.90 | 3.74E-05 | 0.00078036 |
| ENSG00000160593 | AMICA1 | 14.9 | 3.90 | 3.61E-27 | 5.11E-24 |
| ENSG00000215044 | AHCYP1 | 14.89 | 3.90 | 3.71E-06 | 0.000115561 |
| ENSG00000245648 | RP11-277P12.20 | 14.84 | 3.89 | 7.64E-13 | 1.13E-10 |
| ENSG00000137265 | IRF4 | 14.82 | 3.89 | 7.35E-22 | 5.15E-19 |
| ENSG00000085265 | FCN1 | 14.81 | 3.89 | 6.80E-10 | 5.82E-08 |
| ENSG00000153064 | BANK1 | 14.71 | 3.88 | 1.59E-10 | 1.52E-08 |
| ENSG00000136250 | AOAH | 14.67 | 3.87 | 1.51E-22 | 1.13E-19 |
| ENSG00000226557 | TRAF6P1 | 14.58 | 3.87 | 0.00265606 | 0.022028493 |
| ENSG00000109943 | CRTAM | 14.55 | 3.86 | 3.08E-13 | 5.02E-11 |
| ENSG00000117322 | CR2 | 14.54 | 3.86 | 0.007929526 | 0.049052473 |
| ENSG00000255776 | RP11-436I9.3 | 14.37 | 3.85 | 1.76E-06 | 6.09E-05 |
| ENSG00000228314 | CYP4F29P | 14.36 | 3.84 | 4.82E-05 | 0.000964569 |
| ENSG00000147168 | IL2RG | 14.23 | 3.83 | 1.67E-22 | 1.22E-19 |
| ENSG00000240403 | KIR3DL2 | 14.18 | 3.83 | 0.003326204 | 0.026134264 |
| ENSG00000186481 | ANKRD20A5P | 14.14 | 3.82 | 3.76E-11 | 4.02E-09 |
| ENSG00000205810 | KLRC3 | 14.13 | 3.82 | 1.15E-06 | 4.21E-05 |
| ENSG00000174885 | NLRP6 | 13.91 | 3.80 | 3.04E-11 | 3.34E-09 |
| ENSG00000100351 | GRAP2 | 13.7 | 3.78 | 4.53E-20 | 2.37E-17 |
| ENSG00000272279 | RP11-157J24.2 | 13.7 | 3.78 | 0.000452718 | 0.005795049 |
| ENSG00000078589 | P2RY10 | 13.68 | 3.77 | 2.22E-13 | 3.70E-11 |
| ENSG00000226681 | AC020595.1 | 13.58 | 3.76 | 0.0041233 | 0.030458645 |
| ENSG00000204659 | CBY3 | 13.53 | 3.76 | 0.003602303 | 0.027637095 |
| ENSG00000235300 | AC090627.1 | 13.44 | 3.75 | 6.99E-10 | 5.95E-08 |
| ENSG00000174946 | GPR171 | 13.38 | 3.74 | 1.64E-19 | 7.77E-17 |
| ENSG00000169245 | CXCL10 | 13.35 | 3.74 | 1.09E-06 | 4.01E-05 |
| ENSG00000205784 | ARRDC5 | 13.32 | 3.74 | 1.84E-10 | 1.75E-08 |
| ENSG00000228058 | RP11-552D4.1 | 13.3 | 3.73 | 0.004095012 | 0.030310359 |
| ENSG00000240505 | TNFRSF13B | 13.25 | 3.73 | 6.75E-06 | 0.000192032 |
| ENSG00000243836 | WDR86-AS1 | 13.05 | 3.71 | 2.64E-05 | 0.000584087 |
| ENSG00000145649 | GZMA | 13.02 | 3.70 | 2.10E-13 | 3.54E-11 |
| ENSG00000198286 | CARD11 | 13 | 3.70 | 3.95E-16 | 9.98E-14 |
| ENSG00000240535 | CTD-2313F11.1 | 12.97 | 3.70 | 3.91E-06 | 0.000120463 |
| ENSG00000134539 | KLRD1 | 12.82 | 3.68 | 1.31E-20 | 7.31E-18 |
| ENSG00000007264 | MATK | 12.79 | 3.68 | 1.66E-21 | 1.09E-18 |
| ENSG00000187912 | CLEC17A | 12.79 | 3.68 | 0.000215489 | 0.003235588 |
| ENSG00000163508 | EOMES | 12.73 | 3.67 | 8.27E-13 | 1.21E-10 |
| ENSG00000020633 | RUNX3 | 12.58 | 3.65 | 1.71E-25 | 1.82E-22 |
| ENSG00000109956 | B3GAT1 | 12.5 | 3.64 | 2.66E-11 | 2.97E-09 |
| ENSG00000175779 | C15orf53 | 12.49 | 3.64 | 7.76E-07 | 3.00E-05 |
| ENSG00000160856 | FCRL3 | 12.29 | 3.62 | 3.87E-08 | 2.15E-06 |
| ENSG00000242258 | LINC00996 | 12.29 | 3.62 | 1.96E-07 | 8.93E-06 |
| ENSG00000100385 | IL2RB | 12.28 | 3.62 | 1.25E-18 | 5.00E-16 |
| ENSG00000105366 | SIGLEC8 | 12.26 | 3.62 | 1.33E-05 | 0.000334705 |
| ENSG00000121966 | CXCR4 | 12.25 | 3.61 | 1.50E-26 | 2.01E-23 |
| ENSG00000265787 | CYP4F35P | 12.09 | 3.60 | 6.17E-05 | 0.001185365 |
| ENSG00000090382 | LYZ | 12.08 | 3.60 | 7.66E-17 | 2.17E-14 |
| ENSG00000171954 | CYP4F22 | 11.99 | 3.58 | 8.21E-06 | 0.000226244 |
| ENSG00000117091 | CD48 | 11.91 | 3.57 | 1.20E-17 | 4.03E-15 |
| ENSG00000176092 | AIM1L | 11.9 | 3.57 | 1.52E-05 | 0.000372252 |
| ENSG00000132437 | DDC | 11.77 | 3.56 | 0.004563201 | 0.032819042 |
| ENSG00000255819 | KLRC4-KLRK1 | 11.75 | 3.56 | 8.62E-08 | 4.44E-06 |
| ENSG00000182866 | LCK | 11.67 | 3.55 | 1.39E-17 | 4.45E-15 |
| ENSG00000173762 | CD7 | 11.67 | 3.54 | 6.44E-15 | 1.32E-12 |
| ENSG00000057657 | PRDM1 | 11.6 | 3.54 | 2.48E-31 | 6.37E-28 |
| ENSG00000241490 | RP11-553L6.2 | 11.6 | 3.54 | 6.80E-05 | 0.00127867 |
| ENSG00000172543 | CTSW | 11.59 | 3.54 | 3.83E-38 | 3.93E-34 |
| ENSG00000237988 | OR2I1P | 11.59 | 3.53 | 6.93E-05 | 0.001297218 |
| ENSG00000259242 | AC002306.1 | 11.44 | 3.52 | 5.15E-05 | 0.001020139 |
| ENSG00000105374 | NKG7 | 11.37 | 3.51 | 1.74E-21 | 1.12E-18 |
| ENSG00000183813 | CCR4 | 11.37 | 3.51 | 1.02E-13 | 1.77E-11 |
| ENSG00000164483 | SAMD3 | 11.33 | 3.50 | 5.45E-21 | 3.29E-18 |
| ENSG00000197540 | GZMM | 11.31 | 3.50 | 4.36E-13 | 6.81E-11 |
| ENSG00000178773 | CPNE7 | 11.26 | 3.49 | 1.12E-05 | 0.000292338 |
| ENSG00000259834 | RP11-284N8.3 | 11.25 | 3.49 | 4.14E-21 | 2.55E-18 |
| ENSG00000158517 | NCF1 | 11.23 | 3.49 | 2.44E-21 | 1.53E-18 |
| ENSG00000186188 | FFAR4 | 11.16 | 3.48 | 2.35E-05 | 0.000532169 |
| ENSG00000222086 | AC010609.1 | 11.1 | 3.47 | 0.000419021 | 0.005458961 |
| ENSG00000134242 | PTPN22 | 11.05 | 3.47 | 1.11E-21 | 7.42E-19 |
| ENSG00000122025 | FLT3 | 11.04 | 3.47 | 1.26E-06 | 4.54E-05 |
| ENSG00000228427 | RP5-1091N2.9 | 10.99 | 3.46 | 3.64E-05 | 0.000764218 |
| ENSG00000179593 | ALOX15B | 10.94 | 3.45 | 4.72E-05 | 0.00094973 |
| ENSG00000241525 | AC108004.3 | 10.93 | 3.45 | 0.000157685 | 0.002541739 |
| ENSG00000175463 | TBC1D10C | 10.89 | 3.44 | 9.04E-30 | 1.86E-26 |
| ENSG00000197153 | HIST1H3J | 10.89 | 3.44 | 4.11E-06 | 0.000125779 |
| ENSG00000035720 | STAP1 | 10.87 | 3.44 | 1.18E-06 | 4.27E-05 |
| ENSG00000131401 | NAPSB | 10.85 | 3.44 | 1.14E-14 | 2.21E-12 |
| ENSG00000175857 | GAPT | 10.7 | 3.42 | 4.96E-10 | 4.37E-08 |
| ENSG00000204165 | CXorf65 | 10.68 | 3.42 | 6.34E-05 | 0.001209545 |
| ENSG00000207939 | MIR223 | 10.65 | 3.41 | 1.23E-05 | 0.000313407 |
| ENSG00000139626 | ITGB7 | 10.6 | 3.41 | 1.30E-33 | 4.46E-30 |
| ENSG00000157303 | SUSD3 | 10.58 | 3.40 | 4.20E-13 | 6.63E-11 |
| ENSG00000185811 | IKZF1 | 10.56 | 3.40 | 9.31E-24 | 8.44E-21 |
| ENSG00000050730 | TNIP3 | 10.46 | 3.39 | 7.81E-08 | 4.06E-06 |
| ENSG00000152495 | CAMK4 | 10.42 | 3.38 | 3.41E-20 | 1.84E-17 |
| ENSG00000163564 | PYHIN1 | 10.41 | 3.38 | 3.00E-12 | 4.06E-10 |
| ENSG00000272053 | RP11-367G6.3 | 10.36 | 3.37 | 1.52E-05 | 0.000372252 |
| ENSG00000272264 | RP11-92K15.3 | 10.35 | 3.37 | 0.000120971 | 0.002046806 |
| ENSG00000073734 | ABCB11 | 10.31 | 3.37 | 0.002506976 | 0.021087203 |
| ENSG00000205020 | CCL4L1 | 10.3 | 3.37 | 0.000702007 | 0.008046701 |
| ENSG00000185101 | ANO9 | 10.2 | 3.35 | 4.51E-19 | 1.98E-16 |
| ENSG00000110448 | CD5 | 10.18 | 3.35 | 1.55E-16 | 4.15E-14 |
| ENSG00000104814 | MAP4K1 | 10.15 | 3.34 | 4.14E-26 | 5.10E-23 |
| ENSG00000251301 | RP11-81H14.2 | 10.08 | 3.33 | 1.52E-07 | 7.20E-06 |
| ENSG00000236790 | LINC00299 | 10.06 | 3.33 | 1.48E-07 | 7.05E-06 |
| ENSG00000253522 | MIR146A | 10.04 | 3.33 | 2.24E-07 | 1.00E-05 |
| ENSG00000253535 | RP11-624C23.1 | 9.97 | 3.32 | 7.60E-05 | 0.001405199 |
| ENSG00000199377 | RNU5F-1 | 9.96 | 3.32 | 0.000270505 | 0.003892822 |
| ENSG00000250421 | RP11-83M16.6 | 9.87 | 3.30 | 1.82E-05 | 0.000433496 |
| ENSG00000258535 | RP11-280K24.4 | 9.84 | 3.30 | 0.005894203 | 0.039479626 |
| ENSG00000135697 | BCMO1 | 9.83 | 3.30 | 0.000229421 | 0.003409889 |
| ENSG00000250829 | RP11-11N5.1 | 9.78 | 3.29 | 7.18E-05 | 0.001338896 |
| ENSG00000240219 | RP11-430C7.5 | 9.76 | 3.29 | 9.83E-07 | 3.67E-05 |
| ENSG00000088340 | FER1L4 | 9.69 | 3.28 | 9.79E-13 | 1.42E-10 |
| ENSG00000187808 | SOWAHD | 9.66 | 3.27 | 0.000264555 | 0.003823271 |
| ENSG00000256582 | RP11-75L1.1 | 9.65 | 3.27 | 0.000214779 | 0.003229655 |
| ENSG00000232871 | SEC1P | 9.64 | 3.27 | 0.000321148 | 0.004451612 |
| ENSG00000113088 | GZMK | 9.62 | 3.27 | 7.76E-09 | 5.13E-07 |
| ENSG00000127324 | TSPAN8 | 9.62 | 3.27 | 3.48E-05 | 0.00073741 |
| ENSG00000253686 | CTB-43E15.3 | 9.54 | 3.25 | 0.000409406 | 0.005369569 |
| ENSG00000126264 | HCST | 9.5 | 3.25 | 3.61E-22 | 2.59E-19 |
| ENSG00000231128 | RP5-1073O3.2 | 9.49 | 3.25 | 0.00023334 | 0.003460314 |
| ENSG00000115165 | CYTIP | 9.47 | 3.24 | 1.54E-19 | 7.39E-17 |
| ENSG00000197057 | DTHD1 | 9.47 | 3.24 | 2.24E-11 | 2.52E-09 |
| ENSG00000203876 | RP11-451M19.3 | 9.47 | 3.24 | 5.31E-05 | 0.001045951 |
| ENSG00000070190 | DAPP1 | 9.43 | 3.24 | 7.41E-16 | 1.76E-13 |
| ENSG00000199592 | RNA5SP321 | 9.41 | 3.23 | 0.000416191 | 0.005426688 |
| ENSG00000230747 | AC021188.4 | 9.39 | 3.23 | 1.26E-09 | 1.01E-07 |
| ENSG00000102970 | CCL17 | 9.37 | 3.23 | 4.03E-05 | 0.000830247 |
| ENSG00000070915 | SLC12A3 | 9.35 | 3.22 | 0.001163104 | 0.011768928 |
| ENSG00000182487 | NCF1B | 9.32 | 3.22 | 3.19E-16 | 8.27E-14 |
| ENSG00000235785 | AL109767.1 | 9.32 | 3.22 | 0.000119014 | 0.00202258 |
| ENSG00000086300 | SNX10 | 9.29 | 3.22 | 5.50E-14 | 9.84E-12 |
| ENSG00000160791 | CCR5 | 9.29 | 3.22 | 3.46E-12 | 4.60E-10 |
| ENSG00000255026 | RP11-326C3.2 | 9.28 | 3.21 | 9.86E-08 | 4.94E-06 |
| ENSG00000235833 | AC159540.14 | 9.27 | 3.21 | 6.51E-06 | 0.000186539 |
| ENSG00000225783 | MIAT | 9.26 | 3.21 | 4.72E-19 | 2.02E-16 |
| ENSG00000163568 | AIM2 | 9.21 | 3.20 | 5.74E-09 | 3.95E-07 |
| ENSG00000259772 | RP11-16E12.2 | 9.2 | 3.20 | 1.36E-09 | 1.08E-07 |
| ENSG00000100346 | CACNA1I | 9.14 | 3.19 | 5.66E-07 | 2.27E-05 |
| ENSG00000266999 | AC015849.16 | 9.09 | 3.18 | 0.000289111 | 0.004091286 |
| ENSG00000183347 | GBP6 | 9.01 | 3.17 | 1.19E-05 | 0.000307374 |
| ENSG00000167895 | TMC8 | 8.96 | 3.16 | 5.16E-23 | 4.19E-20 |
| ENSG00000082074 | FYB | 8.96 | 3.16 | 9.56E-18 | 3.27E-15 |
| ENSG00000102096 | PIM2 | 8.95 | 3.16 | 6.19E-24 | 5.78E-21 |
| ENSG00000225460 | RP13-93L13.1 | 8.94 | 3.16 | 1.39E-05 | 0.000346921 |
| ENSG00000270933 | CTD-2227E11.1 | 8.92 | 3.16 | 0.000254434 | 0.003708309 |
| ENSG00000100453 | GZMB | 8.91 | 3.16 | 2.87E-12 | 3.91E-10 |
| ENSG00000227507 | LTB | 8.91 | 3.15 | 5.38E-09 | 3.77E-07 |
| ENSG00000164691 | TAGAP | 8.89 | 3.15 | 1.00E-17 | 3.40E-15 |
| ENSG00000158714 | SLAMF8 | 8.89 | 3.15 | 5.42E-14 | 9.77E-12 |
| ENSG00000223750 | SIRPB3P | 8.89 | 3.15 | 0.001579688 | 0.014802852 |
| ENSG00000022556 | NLRP2 | 8.86 | 3.15 | 2.23E-05 | 0.000509592 |
| ENSG00000214787 | MS4A4E | 8.84 | 3.14 | 1.86E-19 | 8.69E-17 |
| ENSG00000213402 | PTPRCAP | 8.84 | 3.14 | 1.26E-17 | 4.18E-15 |
| ENSG00000074706 | IPCEF1 | 8.84 | 3.14 | 2.30E-17 | 7.01E-15 |
| ENSG00000048462 | TNFRSF17 | 8.83 | 3.14 | 6.11E-06 | 0.000176506 |
| ENSG00000265154 | MIR151B | 8.82 | 3.14 | 0.000117946 | 0.002012202 |
| ENSG00000149527 | PLCH2 | 8.81 | 3.14 | 3.77E-13 | 6.05E-11 |
| ENSG00000147138 | GPR174 | 8.79 | 3.14 | 1.93E-05 | 0.000454343 |
| ENSG00000236928 | RP11-792A8.1 | 8.78 | 3.13 | 0.000450518 | 0.005774084 |
| ENSG00000118322 | ATP10B | 8.76 | 3.13 | 3.67E-10 | 3.32E-08 |
| ENSG00000165682 | CLEC1B | 8.75 | 3.13 | 3.85E-05 | 0.000800719 |
| ENSG00000169442 | CD52 | 8.72 | 3.12 | 1.31E-17 | 4.28E-15 |
| ENSG00000179840 | C1orf200 | 8.72 | 3.12 | 0.000299424 | 0.004206814 |
| ENSG00000188305 | C19orf35 | 8.69 | 3.12 | 3.81E-13 | 6.09E-11 |
| ENSG00000252105 | RNU1-143P | 8.66 | 3.11 | 0.000180229 | 0.002818804 |
| ENSG00000171759 | PAH | 8.61 | 3.11 | 0.00048452 | 0.006102384 |
| ENSG00000180644 | PRF1 | 8.6 | 3.10 | 5.88E-28 | 1.01E-24 |
| ENSG00000121807 | CCR2 | 8.51 | 3.09 | 8.93E-17 | 2.48E-14 |
| ENSG00000132185 | FCRLA | 8.51 | 3.09 | 2.18E-06 | 7.39E-05 |
| ENSG00000147889 | CDKN2A | 8.48 | 3.08 | 1.54E-06 | 5.39E-05 |
| ENSG00000183918 | SH2D1A | 8.44 | 3.08 | 1.51E-08 | 9.26E-07 |
| ENSG00000254872 | RP13-870H17.3 | 8.41 | 3.07 | 0.000818661 | 0.008983349 |
| ENSG00000162374 | ELAVL4 | 8.37 | 3.07 | 0.000519026 | 0.006439468 |
| ENSG00000178199 | ZC3H12D | 8.36 | 3.06 | 7.76E-16 | 1.82E-13 |
| ENSG00000185905 | C16orf54 | 8.33 | 3.06 | 2.90E-18 | 1.08E-15 |
| ENSG00000041353 | RAB27B | 8.33 | 3.06 | 1.36E-11 | 1.60E-09 |
| ENSG00000006555 | TTC22 | 8.33 | 3.06 | 2.86E-09 | 2.13E-07 |
| ENSG00000160183 | TMPRSS3 | 8.33 | 3.06 | 0.00014474 | 0.002372112 |
| ENSG00000196533 | C1orf186 | 8.32 | 3.06 | 1.59E-09 | 1.24E-07 |
| ENSG00000102962 | CCL22 | 8.31 | 3.05 | 4.68E-06 | 0.000139849 |
| ENSG00000142512 | SIGLEC10 | 8.28 | 3.05 | 1.86E-05 | 0.00044004 |
| ENSG00000090104 | RGS1 | 8.22 | 3.04 | 1.97E-12 | 2.75E-10 |
| ENSG00000253702 | RP11-567J20.1 | 8.22 | 3.04 | 0.00018404 | 0.002863874 |
| ENSG00000150637 | CD226 | 8.2 | 3.03 | 3.14E-15 | 6.73E-13 |
| ENSG00000229162 | RP11-84D1.1 | 8.18 | 3.03 | 0.000568193 | 0.006841652 |
| ENSG00000220008 | LINGO3 | 8.17 | 3.03 | 1.05E-08 | 6.74E-07 |
| ENSG00000188869 | TMC3 | 8.14 | 3.03 | 1.36E-05 | 0.000338986 |
| ENSG00000234299 | CDK2AP2P1 | 8.13 | 3.02 | 0.000805777 | 0.008885582 |
| ENSG00000259277 | RP13-126C7.1 | 8.13 | 3.02 | 0.001262489 | 0.012540312 |
| ENSG00000158481 | CD1C | 8.09 | 3.02 | 2.51E-08 | 1.48E-06 |
| ENSG00000243323 | PTPRVP | 8.02 | 3.00 | 1.98E-07 | 8.99E-06 |
| ENSG00000123901 | GPR83 | 8.01 | 3.00 | 2.53E-06 | 8.38E-05 |
| ENSG00000135898 | GPR55 | 7.94 | 2.99 | 1.36E-05 | 0.000339858 |
| ENSG00000163534 | FCRL1 | 7.91 | 2.98 | 3.02E-06 | 9.70E-05 |
| ENSG00000198821 | CD247 | 7.86 | 2.97 | 1.95E-12 | 2.73E-10 |
| ENSG00000156475 | PPP2R2B | 7.86 | 2.97 | 1.24E-05 | 0.000315441 |
| ENSG00000088002 | SULT2B1 | 7.85 | 2.97 | 0.000405535 | 0.005335155 |
| ENSG00000079263 | SP140 | 7.84 | 2.97 | 6.10E-20 | 3.08E-17 |
| ENSG00000125910 | S1PR4 | 7.84 | 2.97 | 1.31E-11 | 1.57E-09 |
| ENSG00000272908 | RP11-121A8.1 | 7.83 | 2.97 | 2.31E-07 | 1.03E-05 |
| ENSG00000255987 | RP11-1094M14.4 | 7.77 | 2.96 | 1.32E-09 | 1.05E-07 |
| ENSG00000267554 | RP11-686D22.10 | 7.68 | 2.94 | 4.90E-09 | 3.51E-07 |
| ENSG00000228655 | AC096558.1 | 7.63 | 2.93 | 3.82E-10 | 3.44E-08 |
| ENSG00000243544 | RN7SL172P | 7.6 | 2.93 | 1.72E-05 | 0.000414472 |
| ENSG00000100427 | MLC1 | 7.6 | 2.93 | 0.00028531 | 0.004043557 |
| ENSG00000101057 | MYBL2 | 7.58 | 2.92 | 2.45E-05 | 0.000548999 |
| ENSG00000089692 | LAG3 | 7.56 | 2.92 | 7.12E-09 | 4.74E-07 |
| ENSG00000232953 | HSPA8P18 | 7.56 | 2.92 | 0.000928732 | 0.009898012 |
| ENSG00000182489 | XKRX | 7.55 | 2.92 | 0.000184672 | 0.002870796 |
| ENSG00000136492 | BRIP1 | 7.44 | 2.89 | 1.08E-08 | 6.89E-07 |
| ENSG00000260217 | RP11-809F4.3 | 7.44 | 2.90 | 0.001427755 | 0.01373417 |
| ENSG00000267046 | RP11-1094M14.9 | 7.41 | 2.89 | 1.90E-07 | 8.73E-06 |
| ENSG00000259686 | HNRNPA1P71 | 7.41 | 2.89 | 3.72E-06 | 0.000115698 |
| ENSG00000146070 | PLA2G7 | 7.34 | 2.88 | 1.32E-07 | 6.44E-06 |
| ENSG00000268027 | AC006129.2 | 7.33 | 2.87 | 5.30E-10 | 4.63E-08 |
| ENSG00000165178 | NCF1C | 7.33 | 2.87 | 2.06E-09 | 1.57E-07 |
| ENSG00000228601 | RPL39P | 7.26 | 2.86 | 3.58E-07 | 1.52E-05 |
| ENSG00000096996 | IL12RB1 | 7.25 | 2.86 | 1.03E-09 | 8.59E-08 |
| ENSG00000009790 | TRAF3IP3 | 7.24 | 2.86 | 4.60E-15 | 9.58E-13 |
| ENSG00000158077 | NLRP14 | 7.23 | 2.85 | 0.000160212 | 0.00256697 |
| ENSG00000272282 | RP11-222K16.2 | 7.23 | 2.85 | 0.00016596 | 0.002634398 |
| ENSG00000237593 | RP11-317B3.2 | 7.21 | 2.85 | 1.95E-05 | 0.000457402 |
| ENSG00000226675 | RP11-666A1.3 | 7.21 | 2.85 | 0.001182601 | 0.011907553 |
| ENSG00000101082 | SLA2 | 7.17 | 2.84 | 1.83E-14 | 3.41E-12 |
| ENSG00000101892 | ATP1B4 | 7.17 | 2.84 | 0.0007745 | 0.0086273 |
| ENSG00000198502 | HLA-DRB5 | 7.14 | 2.84 | 4.47E-06 | 0.000134458 |
| ENSG00000250290 | CTC-820M8.1 | 7.14 | 2.84 | 0.001054439 | 0.010916769 |
| ENSG00000124721 | DNAH8 | 7.12 | 2.83 | 1.12E-06 | 4.09E-05 |
| ENSG00000225792 | AC004540.4 | 7.09 | 2.83 | 0.00213259 | 0.018730681 |
| ENSG00000213262 | AL365331.2 | 7.05 | 2.82 | 0.000825588 | 0.009045867 |
| ENSG00000254815 | RP11-496I9.1 | 7.05 | 2.82 | 0.001162798 | 0.011768928 |
| ENSG00000174255 | ZNF80 | 7.03 | 2.81 | 0.000669341 | 0.007753035 |
| ENSG00000261008 | AC004158.2 | 7.02 | 2.81 | 0.000102611 | 0.001793285 |
| ENSG00000169248 | CXCL11 | 7.02 | 2.81 | 0.000437271 | 0.005632428 |
| ENSG00000130475 | FCHO1 | 7 | 2.81 | 2.58E-13 | 4.28E-11 |
| ENSG00000183742 | MACC1 | 7 | 2.81 | 4.23E-08 | 2.34E-06 |
| ENSG00000240487 | RP11-553L6.3 | 6.99 | 2.81 | 0.000111035 | 0.001910163 |
| ENSG00000167780 | SOAT2 | 6.97 | 2.80 | 0.001547638 | 0.014587932 |
| ENSG00000135127 | CCDC64 | 6.96 | 2.80 | 1.32E-10 | 1.28E-08 |
| ENSG00000239636 | RP4-728D4.2 | 6.92 | 2.79 | 1.29E-06 | 4.62E-05 |
| ENSG00000197471 | SPN | 6.88 | 2.78 | 1.06E-14 | 2.07E-12 |
| ENSG00000229757 | RP4-738P11.4 | 6.88 | 2.78 | 0.00080726 | 0.00889232 |
| ENSG00000167208 | SNX20 | 6.87 | 2.78 | 1.96E-14 | 3.64E-12 |
| ENSG00000166501 | PRKCB | 6.86 | 2.78 | 3.09E-15 | 6.70E-13 |
| ENSG00000231758 | AC092652.1 | 6.85 | 2.78 | 4.61E-09 | 3.33E-07 |
| ENSG00000163600 | ICOS | 6.85 | 2.78 | 0.000264037 | 0.003819367 |
| ENSG00000143851 | PTPN7 | 6.84 | 2.77 | 2.58E-12 | 3.53E-10 |
| ENSG00000215559 | ANKRD20A11P | 6.83 | 2.77 | 5.62E-08 | 2.98E-06 |
| ENSG00000224689 | ZNF812 | 6.81 | 2.77 | 4.91E-16 | 1.22E-13 |
| ENSG00000173200 | PARP15 | 6.79 | 2.76 | 4.77E-13 | 7.39E-11 |
| ENSG00000110934 | BIN2 | 6.77 | 2.76 | 5.12E-17 | 1.48E-14 |
| ENSG00000023892 | DEF6 | 6.77 | 2.76 | 4.76E-16 | 1.19E-13 |
| ENSG00000166278 | C2 | 6.77 | 2.76 | 5.62E-13 | 8.65E-11 |
| ENSG00000156127 | BATF | 6.77 | 2.76 | 6.98E-09 | 4.66E-07 |
| ENSG00000253651 | SOD1P3 | 6.77 | 2.76 | 0.000707125 | 0.008090315 |
| ENSG00000087589 | CASS4 | 6.74 | 2.75 | 4.74E-23 | 3.95E-20 |
| ENSG00000237943 | PRKCQ-AS1 | 6.72 | 2.75 | 2.86E-08 | 1.66E-06 |
| ENSG00000187862 | TTC24 | 6.72 | 2.75 | 5.59E-06 | 0.000163787 |
| ENSG00000159904 | ZNF890P | 6.68 | 2.74 | 0.002563835 | 0.02146585 |
| ENSG00000182611 | HIST1H2AJ | 6.66 | 2.73 | 1.08E-07 | 5.37E-06 |
| ENSG00000225528 | RP3-370M22.8 | 6.66 | 2.74 | 7.73E-05 | 0.001424873 |
| ENSG00000257284 | RP11-190J23.1 | 6.64 | 2.73 | 7.70E-08 | 4.02E-06 |
| ENSG00000241717 | VWFP1 | 6.64 | 2.73 | 0.000139615 | 0.002309113 |
| ENSG00000261218 | RP11-960L18.1 | 6.64 | 2.73 | 0.00121092 | 0.012139879 |
| ENSG00000235192 | AC009495.2 | 6.63 | 2.73 | 0.000377436 | 0.005053254 |
| ENSG00000248599 | RP11-302J23.1 | 6.61 | 2.72 | 3.59E-07 | 1.52E-05 |
| ENSG00000232979 | AC092580.2 | 6.6 | 2.72 | 1.81E-05 | 0.000431609 |
| ENSG00000150625 | GPM6A | 6.6 | 2.72 | 1.92E-05 | 0.000452638 |
| ENSG00000247193 | RP11-431M7.3 | 6.57 | 2.72 | 0.001403965 | 0.013567619 |
| ENSG00000116852 | KIF21B | 6.56 | 2.71 | 5.15E-17 | 1.48E-14 |
| ENSG00000111796 | KLRB1 | 6.55 | 2.71 | 8.00E-07 | 3.08E-05 |
| ENSG00000237567 | RP3-359N14.2 | 6.54 | 2.71 | 0.002771133 | 0.022762299 |
| ENSG00000143452 | HORMAD1 | 6.52 | 2.70 | 0.001020008 | 0.010628158 |
| ENSG00000167261 | DPEP2 | 6.51 | 2.70 | 2.08E-12 | 2.87E-10 |
| ENSG00000146192 | FGD2 | 6.5 | 2.70 | 5.98E-16 | 1.45E-13 |
| ENSG00000095585 | BLNK | 6.49 | 2.70 | 9.06E-17 | 2.49E-14 |
| ENSG00000151883 | PARP8 | 6.48 | 2.70 | 7.34E-19 | 3.06E-16 |
| ENSG00000159753 | RLTPR | 6.47 | 2.69 | 3.08E-13 | 5.02E-11 |
| ENSG00000271856 | RP11-861A13.4 | 6.43 | 2.69 | 8.79E-05 | 0.001584439 |
| ENSG00000229474 | PATL2 | 6.4 | 2.68 | 8.37E-12 | 1.04E-09 |
| ENSG00000165383 | LRRC18 | 6.37 | 2.67 | 4.12E-07 | 1.72E-05 |
| ENSG00000181291 | TMEM132E | 6.35 | 2.67 | 0.000125805 | 0.00211969 |
| ENSG00000081237 | PTPRC | 6.34 | 2.66 | 2.48E-15 | 5.54E-13 |
| ENSG00000156510 | HKDC1 | 6.32 | 2.66 | 1.21E-05 | 0.000311131 |
| ENSG00000143184 | XCL1 | 6.31 | 2.66 | 2.10E-06 | 7.16E-05 |
| ENSG00000225039 | LINC01058 | 6.31 | 2.66 | 0.001557134 | 0.014640478 |
| ENSG00000185792 | NLRP9 | 6.3 | 2.65 | 0.001330316 | 0.013024585 |
| ENSG00000177669 | MBOAT4 | 6.28 | 2.65 | 2.94E-05 | 0.000640065 |
| ENSG00000115232 | ITGA4 | 6.27 | 2.65 | 5.40E-16 | 1.33E-13 |
| ENSG00000237940 | AC093642.3 | 6.27 | 2.65 | 1.23E-06 | 4.45E-05 |
| ENSG00000259598 | RP11-275I4.1 | 6.27 | 2.65 | 0.000309361 | 0.004322172 |
| ENSG00000121053 | EPX | 6.27 | 2.65 | 0.000610078 | 0.007216849 |
| ENSG00000218991 | CCNG1P1 | 6.25 | 2.64 | 0.003034612 | 0.02441454 |
| ENSG00000230175 | RP11-466F5.3 | 6.24 | 2.64 | 0.001075625 | 0.011082799 |
| ENSG00000266705 | MIR4437 | 6.21 | 2.63 | 0.002121655 | 0.01866124 |
| ENSG00000105246 | EBI3 | 6.2 | 2.63 | 2.16E-08 | 1.30E-06 |
| ENSG00000196735 | HLA-DQA1 | 6.19 | 2.63 | 2.61E-10 | 2.40E-08 |
| ENSG00000100985 | MMP9 | 6.16 | 2.62 | 0.000284346 | 0.004033609 |
| ENSG00000237840 | FAM21FP | 6.14 | 2.62 | 1.94E-05 | 0.000455936 |
| ENSG00000111536 | IL26 | 6.14 | 2.62 | 0.000307845 | 0.004309406 |
| ENSG00000077984 | CST7 | 6.13 | 2.62 | 6.85E-16 | 1.65E-13 |
| ENSG00000130487 | KLHDC7B | 6.13 | 2.62 | 3.99E-05 | 0.000824847 |
| ENSG00000139970 | RTN1 | 6.11 | 2.61 | 8.84E-15 | 1.78E-12 |
| ENSG00000185760 | KCNQ5 | 6.09 | 2.61 | 7.72E-05 | 0.00142357 |
| ENSG00000254900 | RP11-152H18.4 | 6.08 | 2.60 | 0.000277805 | 0.003973744 |
| ENSG00000205436 | EXOC3L4 | 6.07 | 2.60 | 8.63E-08 | 4.44E-06 |
| ENSG00000271725 | RP11-761I4.4 | 6.07 | 2.60 | 0.000190069 | 0.002930549 |
| ENSG00000066294 | CD84 | 6.05 | 2.60 | 4.62E-14 | 8.42E-12 |
| ENSG00000105967 | TFEC | 6.05 | 2.60 | 3.23E-12 | 4.31E-10 |
| ENSG00000147234 | FRMPD3 | 6.02 | 2.59 | 4.54E-07 | 1.87E-05 |
| ENSG00000226979 | LTA | 6.02 | 2.59 | 0.001264166 | 0.012540312 |
| ENSG00000188452 | CERKL | 5.99 | 2.58 | 1.43E-11 | 1.67E-09 |
| ENSG00000110876 | SELPLG | 5.94 | 2.57 | 2.08E-18 | 7.87E-16 |
| ENSG00000267369 | RP11-1094M14.8 | 5.92 | 2.57 | 5.28E-09 | 3.72E-07 |
| ENSG00000073737 | DHRS9 | 5.92 | 2.57 | 1.66E-08 | 1.01E-06 |
| ENSG00000215863 | LINC01138 | 5.92 | 2.57 | 0.002644408 | 0.021943671 |
| ENSG00000257829 | RP11-845M18.6 | 5.91 | 2.56 | 0.001643892 | 0.015246826 |
| ENSG00000105122 | RASAL3 | 5.9 | 2.56 | 4.05E-12 | 5.29E-10 |
| ENSG00000236278 | PEBP1P3 | 5.9 | 2.56 | 2.44E-07 | 1.08E-05 |
| ENSG00000135905 | DOCK10 | 5.89 | 2.56 | 3.23E-17 | 9.65E-15 |
| ENSG00000128340 | RAC2 | 5.89 | 2.56 | 5.75E-13 | 8.81E-11 |
| ENSG00000254205 | RP11-92K15.1 | 5.89 | 2.56 | 0.000959733 | 0.010156459 |
| ENSG00000132514 | CLEC10A | 5.88 | 2.56 | 6.96E-13 | 1.04E-10 |
| ENSG00000112799 | LY86 | 5.87 | 2.55 | 9.17E-09 | 5.92E-07 |
| ENSG00000106948 | AKNA | 5.85 | 2.55 | 6.51E-18 | 2.25E-15 |
| ENSG00000261573 | RP11-553K8.5 | 5.85 | 2.55 | 5.47E-07 | 2.20E-05 |
| ENSG00000227403 | AC009299.3 | 5.84 | 2.55 | 0.000545749 | 0.006662074 |
| ENSG00000071909 | MYO3B | 5.83 | 2.54 | 2.07E-05 | 0.000478411 |
| ENSG00000188596 | C12orf55 | 5.82 | 2.54 | 1.23E-13 | 2.10E-11 |
| ENSG00000269404 | SPIB | 5.81 | 2.54 | 0.00177074 | 0.016165409 |
| ENSG00000205744 | DENND1C | 5.79 | 2.53 | 3.16E-11 | 3.42E-09 |
| ENSG00000181631 | P2RY13 | 5.78 | 2.53 | 1.87E-09 | 1.43E-07 |
| ENSG00000140368 | PSTPIP1 | 5.77 | 2.53 | 1.67E-14 | 3.18E-12 |
| ENSG00000240007 | RP6-206I17.4 | 5.77 | 2.53 | 0.00056235 | 0.006794994 |
| ENSG00000221476 | MIR1827 | 5.76 | 2.53 | 0.002156919 | 0.018890519 |
| ENSG00000077420 | APBB1IP | 5.75 | 2.52 | 1.53E-12 | 2.16E-10 |
| ENSG00000177807 | KCNJ10 | 5.75 | 2.52 | 0.000165139 | 0.002626796 |
| ENSG00000187037 | GPR141 | 5.72 | 2.52 | 4.86E-06 | 0.000144281 |
| ENSG00000172322 | CLEC12A | 5.72 | 2.52 | 6.00E-06 | 0.000173619 |
| ENSG00000137101 | CD72 | 5.71 | 2.51 | 6.91E-07 | 2.72E-05 |
| ENSG00000104972 | LILRB1 | 5.68 | 2.50 | 3.29E-08 | 1.88E-06 |
| ENSG00000264386 | MIR4513 | 5.66 | 2.50 | 0.003544907 | 0.027353398 |
| ENSG00000064547 | LPAR2 | 5.65 | 2.50 | 2.08E-11 | 2.40E-09 |
| ENSG00000267293 | RP11-8H2.1 | 5.65 | 2.50 | 6.52E-05 | 0.001238503 |
| ENSG00000101916 | TLR8 | 5.64 | 2.50 | 2.09E-08 | 1.26E-06 |
| ENSG00000180096 | sept-01 | 5.61 | 2.49 | 6.73E-09 | 4.52E-07 |
| ENSG00000226751 | AF127936.5 | 5.6 | 2.48 | 7.05E-06 | 0.000199153 |
| ENSG00000229990 | RP11-574K11.8 | 5.59 | 2.48 | 0.001340942 | 0.013107795 |
| ENSG00000252985 | SNORD116 | 5.58 | 2.48 | 8.50E-05 | 0.001543361 |
| ENSG00000236474 | GCNT1P1 | 5.57 | 2.48 | 0.001264144 | 0.012540312 |
| ENSG00000232815 | LINC00537 | 5.56 | 2.47 | 1.09E-09 | 8.97E-08 |
| ENSG00000184292 | TACSTD2 | 5.55 | 2.47 | 4.06E-05 | 0.000834938 |
| ENSG00000204252 | HLA-DOA | 5.53 | 2.47 | 1.94E-17 | 6.02E-15 |
| ENSG00000212456 | RNVU1-13 | 5.53 | 2.47 | 0.000105006 | 0.001828915 |
| ENSG00000226287 | TMEM191A | 5.52 | 2.46 | 4.61E-06 | 0.000138163 |
| ENSG00000147570 | DNAJC5B | 5.52 | 2.46 | 0.003347781 | 0.026253109 |
| ENSG00000140968 | IRF8 | 5.51 | 2.46 | 7.89E-13 | 1.16E-10 |
| ENSG00000110324 | IL10RA | 5.5 | 2.46 | 8.33E-16 | 1.93E-13 |
| ENSG00000117009 | KMO | 5.5 | 2.46 | 1.80E-07 | 8.32E-06 |
| ENSG00000256937 | RP11-436I9.5 | 5.5 | 2.46 | 5.88E-07 | 2.36E-05 |
| ENSG00000255080 | RP11-1082L8.3 | 5.5 | 2.46 | 0.004013248 | 0.029939994 |
| ENSG00000152229 | PSTPIP2 | 5.49 | 2.46 | 3.94E-11 | 4.20E-09 |
| ENSG00000143119 | CD53 | 5.48 | 2.45 | 9.06E-12 | 1.12E-09 |
| ENSG00000136167 | LCP1 | 5.46 | 2.45 | 3.74E-10 | 3.37E-08 |
| ENSG00000186891 | TNFRSF18 | 5.45 | 2.45 | 1.25E-06 | 4.50E-05 |
| ENSG00000250654 | RP11-834C11.7 | 5.45 | 2.45 | 0.001507931 | 0.014317689 |
| ENSG00000107099 | DOCK8 | 5.43 | 2.44 | 1.94E-15 | 4.42E-13 |
| ENSG00000160255 | ITGB2 | 5.4 | 2.43 | 9.88E-11 | 9.86E-09 |
| ENSG00000232687 | RPL12P9 | 5.4 | 2.43 | 0.00398062 | 0.029775888 |
| ENSG00000161929 | SCIMP | 5.38 | 2.43 | 9.25E-12 | 1.13E-09 |
| ENSG00000132518 | GUCY2D | 5.36 | 2.42 | 0.000968591 | 0.0102308 |
| ENSG00000155926 | SLA | 5.34 | 2.42 | 6.91E-13 | 1.04E-10 |
| ENSG00000134516 | DOCK2 | 5.32 | 2.41 | 4.10E-15 | 8.65E-13 |
| ENSG00000172794 | RAB37 | 5.32 | 2.41 | 2.74E-10 | 2.51E-08 |
| ENSG00000135749 | PCNXL2 | 5.31 | 2.41 | 2.84E-16 | 7.40E-14 |
| ENSG00000108370 | RGS9 | 5.3 | 2.41 | 4.91E-08 | 2.66E-06 |
| ENSG00000227777 | RP4-738P11.3 | 5.28 | 2.40 | 0.000760235 | 0.008517672 |
| ENSG00000128011 | LRFN1 | 5.26 | 2.39 | 8.17E-06 | 0.000225296 |
| ENSG00000180448 | HMHA1 | 5.25 | 2.39 | 1.67E-16 | 4.42E-14 |
| ENSG00000089847 | ANKRD24 | 5.25 | 2.39 | 2.87E-08 | 1.66E-06 |
| ENSG00000230362 | RP11-809F4.2 | 5.25 | 2.39 | 2.91E-05 | 0.000634903 |
| ENSG00000010610 | CD4 | 5.24 | 2.39 | 1.56E-13 | 2.65E-11 |
| ENSG00000231621 | AC013264.2 | 5.24 | 2.39 | 0.000119318 | 0.002026292 |
| ENSG00000145287 | PLAC8 | 5.22 | 2.38 | 1.12E-09 | 9.23E-08 |
| ENSG00000007129 | CEACAM21 | 5.22 | 2.38 | 4.14E-08 | 2.29E-06 |
| ENSG00000185338 | SOCS1 | 5.19 | 2.37 | 4.20E-09 | 3.05E-07 |
| ENSG00000187398 | LUZP2 | 5.19 | 2.37 | 0.000768172 | 0.0085941 |
| ENSG00000271631 | RP11-408O19.5 | 5.19 | 2.38 | 0.002497959 | 0.021034332 |
| ENSG00000122122 | SASH3 | 5.18 | 2.37 | 3.36E-11 | 3.62E-09 |
| ENSG00000267539 | RP11-138H8.7 | 5.16 | 2.37 | 0.003721087 | 0.028273835 |
| ENSG00000115523 | GNLY | 5.15 | 2.37 | 6.69E-11 | 6.85E-09 |
| ENSG00000214514 | KRT42P | 5.15 | 2.37 | 0.002555756 | 0.021429725 |
| ENSG00000185862 | EVI2B | 5.14 | 2.36 | 3.11E-12 | 4.18E-10 |
| ENSG00000138964 | PARVG | 5.13 | 2.36 | 1.05E-14 | 2.06E-12 |
| ENSG00000185522 | C11orf35 | 5.13 | 2.36 | 2.59E-05 | 0.000575381 |
| ENSG00000248568 | KRT8P48 | 5.13 | 2.36 | 0.000946209 | 0.010052981 |
| ENSG00000167634 | NLRP7 | 5.12 | 2.36 | 0.002659872 | 0.02204824 |
| ENSG00000237499 | RP11-356I2.4 | 5.11 | 2.35 | 6.18E-12 | 7.81E-10 |
| ENSG00000102879 | CORO1A | 5.11 | 2.35 | 3.12E-11 | 3.40E-09 |
| ENSG00000228278 | ORM2 | 5.11 | 2.35 | 7.69E-05 | 0.001419659 |
| ENSG00000248529 | RP11-2O17.2 | 5.09 | 2.35 | 0.00110907 | 0.011356453 |
| ENSG00000246526 | RP11-539L10.2 | 5.08 | 2.34 | 3.24E-06 | 0.000103428 |
| ENSG00000205045 | SLFN12L | 5.07 | 2.34 | 2.73E-08 | 1.60E-06 |
| ENSG00000175643 | RMI2 | 5.07 | 2.34 | 5.02E-06 | 0.000148728 |
| ENSG00000261208 | RP11-452D12.1 | 5.07 | 2.34 | 6.57E-06 | 0.000187844 |
| ENSG00000136541 | ERMN | 5.05 | 2.34 | 3.39E-08 | 1.92E-06 |
| ENSG00000103056 | SMPD3 | 5.04 | 2.33 | 8.94E-08 | 4.57E-06 |
| ENSG00000165171 | WBSCR27 | 5.04 | 2.33 | 1.86E-05 | 0.00044004 |
| ENSG00000261439 | CTD-2050B12.2 | 5.04 | 2.33 | 0.000120061 | 0.002034765 |
| ENSG00000269220 | LINC00528 | 5.01 | 2.33 | 4.05E-06 | 0.00012408 |
| ENSG00000225978 | HAR1A | 5.01 | 2.33 | 0.000528452 | 0.006523294 |
| ENSG00000214093 | RP11-247I13.3 | 5.01 | 2.32 | 0.003275674 | 0.025865399 |
| ENSG00000258733 | CTD-2341M24.1 | 5 | 2.32 | 3.51E-08 | 1.97E-06 |
| ENSG00000236467 | RP11-443A13.5 | 5 | 2.32 | 2.77E-05 | 0.000609505 |
| ENSG00000254838 | GVINP1 | 4.99 | 2.32 | 6.95E-13 | 1.04E-10 |
| ENSG00000147378 | FATE1 | 4.99 | 2.32 | 0.000125009 | 0.002108186 |
| ENSG00000064787 | BCAS1 | 4.99 | 2.32 | 0.002484553 | 0.020944342 |
| ENSG00000229677 | RP11-383F6.1 | 4.98 | 2.31 | 1.44E-05 | 0.00035671 |
| ENSG00000170571 | EMB | 4.97 | 2.31 | 1.17E-19 | 5.74E-17 |
| ENSG00000106952 | TNFSF8 | 4.97 | 2.31 | 5.62E-09 | 3.88E-07 |
| ENSG00000260487 | RP11-297C4.3 | 4.97 | 2.31 | 0.004159177 | 0.030642854 |
| ENSG00000168546 | GFRA2 | 4.96 | 2.31 | 2.23E-19 | 1.03E-16 |
| ENSG00000257289 | RP11-611O2.6 | 4.96 | 2.31 | 4.59E-05 | 0.000928856 |
| ENSG00000183484 | GPR132 | 4.94 | 2.31 | 3.14E-12 | 4.20E-10 |
| ENSG00000230399 | RBBP8P1 | 4.94 | 2.30 | 0.000328911 | 0.004526168 |
| ENSG00000118513 | MYB | 4.92 | 2.30 | 0.000267864 | 0.003862027 |
| ENSG00000128815 | WDFY4 | 4.91 | 2.30 | 1.54E-11 | 1.78E-09 |
| ENSG00000227681 | RP11-307P5.1 | 4.91 | 2.30 | 0.00276811 | 0.022749592 |
| ENSG00000222057 | RNU4-62P | 4.9 | 2.29 | 2.67E-05 | 0.000591413 |
| ENSG00000260727 | SLC7A5P1 | 4.89 | 2.29 | 1.61E-07 | 7.60E-06 |
| ENSG00000247774 | PCED1B-AS1 | 4.88 | 2.29 | 2.90E-09 | 2.15E-07 |
| ENSG00000204767 | FAM196B | 4.88 | 2.29 | 1.13E-08 | 7.13E-07 |
| ENSG00000261618 | RP11-79H23.3 | 4.88 | 2.29 | 0.001120116 | 0.011448561 |
| ENSG00000180353 | HCLS1 | 4.87 | 2.28 | 5.98E-16 | 1.45E-13 |
| ENSG00000135426 | TESPA1 | 4.85 | 2.28 | 5.93E-08 | 3.13E-06 |
| ENSG00000159307 | SCUBE1 | 4.85 | 2.28 | 0.000239008 | 0.003525169 |
| ENSG00000225342 | AC079630.4 | 4.85 | 2.28 | 0.001262834 | 0.012540312 |
| ENSG00000196550 | FAM72A | 4.84 | 2.27 | 7.56E-07 | 2.94E-05 |
| ENSG00000183831 | ANKRD45 | 4.82 | 2.27 | 0.000513907 | 0.006382104 |
| ENSG00000261292 | RP11-389G6.3 | 4.82 | 2.27 | 0.003834936 | 0.028929854 |
| ENSG00000216009 | MIR874 | 4.81 | 2.27 | 0.002116421 | 0.018631159 |
| ENSG00000213071 | LPAL2 | 4.8 | 2.26 | 7.59E-06 | 0.000211807 |
| ENSG00000124343 | XG | 4.8 | 2.26 | 3.74E-05 | 0.000780822 |
| ENSG00000263806 | AL592188.3 | 4.8 | 2.26 | 0.000762177 | 0.00853633 |
| ENSG00000244620 | AL122127.25 | 4.8 | 2.26 | 0.004187437 | 0.030792149 |
| ENSG00000172349 | IL16 | 4.79 | 2.26 | 3.97E-15 | 8.45E-13 |
| ENSG00000168071 | CCDC88B | 4.78 | 2.26 | 1.14E-16 | 3.11E-14 |
| ENSG00000125637 | PSD4 | 4.77 | 2.25 | 3.95E-11 | 4.20E-09 |
| ENSG00000054219 | LY75 | 4.77 | 2.25 | 4.31E-10 | 3.83E-08 |
| ENSG00000263642 | MIR4802 | 4.76 | 2.25 | 0.004326877 | 0.031576364 |
| ENSG00000112297 | AIM1 | 4.73 | 2.24 | 4.59E-15 | 9.58E-13 |
| ENSG00000236334 | PPIAL4G | 4.73 | 2.24 | 0.00210318 | 0.018551694 |
| ENSG00000172653 | C17orf66 | 4.71 | 2.24 | 0.000324042 | 0.004481178 |
| ENSG00000164266 | SPINK1 | 4.71 | 2.24 | 0.000627687 | 0.007387185 |
| ENSG00000106789 | CORO2A | 4.7 | 2.23 | 1.14E-11 | 1.39E-09 |
| ENSG00000106785 | TRIM14 | 4.7 | 2.23 | 3.33E-11 | 3.60E-09 |
| ENSG00000182010 | RTKN2 | 4.7 | 2.23 | 2.43E-06 | 8.09E-05 |
| ENSG00000223865 | HLA-DPB1 | 4.69 | 2.23 | 5.96E-13 | 9.09E-11 |
| ENSG00000053524 | MCF2L2 | 4.69 | 2.23 | 4.42E-07 | 1.82E-05 |
| ENSG00000035499 | DEPDC1B | 4.69 | 2.23 | 0.003322812 | 0.026123794 |
| ENSG00000161905 | ALOX15 | 4.68 | 2.23 | 0.001304096 | 0.012829025 |
| ENSG00000010671 | BTK | 4.67 | 2.22 | 8.40E-12 | 1.04E-09 |
| ENSG00000197146 | AL133458.1 | 4.67 | 2.22 | 7.04E-06 | 0.000199097 |
| ENSG00000198846 | TOX | 4.64 | 2.21 | 7.34E-11 | 7.49E-09 |
| ENSG00000167984 | NLRC3 | 4.63 | 2.21 | 1.32E-11 | 1.57E-09 |
| ENSG00000133321 | RARRES3 | 4.63 | 2.21 | 5.46E-08 | 2.91E-06 |
| ENSG00000196374 | HIST1H2BM | 4.63 | 2.21 | 0.000628098 | 0.007389199 |
| ENSG00000174332 | GLIS1 | 4.63 | 2.21 | 0.00113475 | 0.011561771 |
| ENSG00000198223 | CSF2RA | 4.62 | 2.21 | 5.13E-12 | 6.54E-10 |
| ENSG00000265148 | BZRAP1-AS1 | 4.62 | 2.21 | 9.32E-06 | 0.000252303 |
| ENSG00000056558 | TRAF1 | 4.58 | 2.20 | 1.70E-14 | 3.20E-12 |
| ENSG00000141506 | PIK3R5 | 4.58 | 2.20 | 1.35E-11 | 1.60E-09 |
| ENSG00000182557 | SPNS3 | 4.58 | 2.19 | 7.70E-06 | 0.000214827 |
| ENSG00000237914 | RP11-77C3.3 | 4.58 | 2.20 | 0.000156362 | 0.002524989 |
| ENSG00000238113 | RP11-262H14.1 | 4.57 | 2.19 | 1.69E-09 | 1.30E-07 |
| ENSG00000233098 | RP11-344E13.3 | 4.57 | 2.19 | 2.88E-08 | 1.66E-06 |
| ENSG00000189430 | NCR1 | 4.57 | 2.19 | 1.54E-05 | 0.000377416 |
| ENSG00000143320 | CRABP2 | 4.56 | 2.19 | 2.62E-05 | 0.000581551 |
| ENSG00000273142 | RP11-458F8.4 | 4.56 | 2.19 | 0.000106704 | 0.001847005 |
| ENSG00000036565 | SLC18A1 | 4.56 | 2.19 | 0.00591432 | 0.039562769 |
| ENSG00000138795 | LEF1 | 4.55 | 2.19 | 9.13E-08 | 4.63E-06 |
| ENSG00000258929 | RP11-58E21.3 | 4.54 | 2.18 | 5.68E-06 | 0.000165197 |
| ENSG00000145088 | EAF2 | 4.53 | 2.18 | 8.30E-14 | 1.45E-11 |
| ENSG00000084070 | SMAP2 | 4.52 | 2.18 | 2.93E-15 | 6.45E-13 |
| ENSG00000171659 | GPR34 | 4.52 | 2.18 | 1.10E-13 | 1.90E-11 |
| ENSG00000133246 | PRAM1 | 4.52 | 2.18 | 2.44E-05 | 0.000547618 |
| ENSG00000178445 | GLDC | 4.51 | 2.17 | 4.06E-05 | 0.000836008 |
| ENSG00000223804 | RP6-206I17.1 | 4.5 | 2.17 | 5.59E-09 | 3.88E-07 |
| ENSG00000249713 | CTD-2236F14.1 | 4.49 | 2.17 | 1.05E-05 | 0.000278939 |
| ENSG00000259415 | RP11-7M10.2 | 4.48 | 2.16 | 0.003695532 | 0.028135167 |
| ENSG00000136867 | SLC31A2 | 4.47 | 2.16 | 6.16E-09 | 4.18E-07 |
| ENSG00000180828 | BHLHE22 | 4.47 | 2.16 | 5.21E-07 | 2.11E-05 |
| ENSG00000237484 | AP000476.1 | 4.47 | 2.16 | 3.50E-06 | 0.000110421 |
| ENSG00000234425 | RP11-528G1.2 | 4.47 | 2.16 | 0.00352865 | 0.027275776 |
| ENSG00000154252 | GAL3ST2 | 4.47 | 2.16 | 0.004774537 | 0.033974196 |
| ENSG00000257277 | RP11-434H14.1 | 4.46 | 2.16 | 6.56E-06 | 0.000187548 |
| ENSG00000231858 | AC067945.4 | 4.45 | 2.15 | 4.28E-06 | 0.000130005 |
| ENSG00000206875 | RNU6-761P | 4.45 | 2.16 | 0.003541438 | 0.027340324 |
| ENSG00000183508 | FAM46C | 4.44 | 2.15 | 2.13E-10 | 1.99E-08 |
| ENSG00000187904 | AC097382.5 | 4.44 | 2.15 | 0.004406765 | 0.032015285 |
| ENSG00000203813 | HIST1H3H | 4.43 | 2.15 | 1.22E-09 | 9.88E-08 |
| ENSG00000120498 | TEX11 | 4.43 | 2.15 | 0.004850061 | 0.034408294 |
| ENSG00000251889 | RNU4-49P | 4.43 | 2.15 | 0.005771854 | 0.038841903 |
| ENSG00000015285 | WAS | 4.42 | 2.15 | 4.23E-10 | 3.76E-08 |
| ENSG00000124575 | HIST1H1D | 4.42 | 2.15 | 1.82E-09 | 1.40E-07 |
| ENSG00000249988 | RP11-669M16.1 | 4.42 | 2.14 | 0.004634642 | 0.033185671 |
| ENSG00000112232 | KHDRBS2 | 4.41 | 2.14 | 0.00028309 | 0.00402134 |
| ENSG00000253525 | CTD-2114J12.1 | 4.41 | 2.14 | 0.003067593 | 0.024632683 |
| ENSG00000121594 | CD80 | 4.4 | 2.14 | 7.19E-06 | 0.00020204 |
| ENSG00000267246 | RP11-798G7.7 | 4.4 | 2.14 | 0.000917709 | 0.009821306 |
| ENSG00000145569 | FAM105A | 4.39 | 2.13 | 1.43E-18 | 5.64E-16 |
| ENSG00000104894 | CD37 | 4.39 | 2.13 | 2.23E-11 | 2.52E-09 |
| ENSG00000147443 | DOK2 | 4.39 | 2.13 | 1.22E-08 | 7.63E-07 |
| ENSG00000186074 | CD300LF | 4.39 | 2.13 | 2.71E-05 | 0.000598971 |
| ENSG00000250614 | AC007078.4 | 4.39 | 2.13 | 0.004572137 | 0.032867967 |
| ENSG00000150681 | RGS18 | 4.38 | 2.13 | 6.77E-10 | 5.81E-08 |
| ENSG00000118242 | MREG | 4.38 | 2.13 | 9.75E-07 | 3.65E-05 |
| ENSG00000110077 | MS4A6A | 4.37 | 2.13 | 4.29E-13 | 6.74E-11 |
| ENSG00000116990 | MYCL | 4.37 | 2.13 | 8.46E-06 | 0.000232342 |
| ENSG00000258912 | RP11-1079H9.1 | 4.37 | 2.13 | 0.000611729 | 0.007229759 |
| ENSG00000236946 | HNRNPA1P70 | 4.35 | 2.12 | 6.95E-06 | 0.000196896 |
| ENSG00000257640 | RP11-570L15.2 | 4.35 | 2.12 | 1.72E-05 | 0.000414472 |
| ENSG00000118308 | LRMP | 4.34 | 2.12 | 1.26E-12 | 1.81E-10 |
| ENSG00000121933 | ADORA3 | 4.34 | 2.12 | 2.58E-10 | 2.37E-08 |
| ENSG00000230035 | RP11-174G17 | 4.34 | 2.12 | 0.002716627 | 0.022404172 |
| ENSG00000182572 | HIST1H3I | 4.33 | 2.11 | 3.21E-07 | 1.38E-05 |
| ENSG00000230649 | AC024084.1 | 4.33 | 2.12 | 0.004574732 | 0.03287895 |
| ENSG00000075884 | ARHGAP15 | 4.32 | 2.11 | 7.77E-15 | 1.58E-12 |
| ENSG00000043462 | LCP2 | 4.32 | 2.11 | 4.07E-11 | 4.26E-09 |
| ENSG00000255780 | RP11-1029F8.1 | 4.32 | 2.11 | 0.003173894 | 0.025242863 |
| ENSG00000124496 | TRERF1 | 4.31 | 2.11 | 6.47E-13 | 9.82E-11 |
| ENSG00000162888 | C1orf147 | 4.3 | 2.10 | 0.000357561 | 0.004834049 |
| ENSG00000256708 | RP11-444B24.2 | 4.3 | 2.10 | 0.00223056 | 0.019327625 |
| ENSG00000127084 | FGD3 | 4.29 | 2.10 | 9.97E-15 | 1.98E-12 |
| ENSG00000103490 | PYCARD | 4.28 | 2.10 | 2.52E-10 | 2.32E-08 |
| ENSG00000102575 | ACP5 | 4.28 | 2.10 | 3.46E-07 | 1.48E-05 |
| ENSG00000188610 | FAM72B | 4.28 | 2.10 | 0.000118503 | 0.002018345 |
| ENSG00000228991 | RP11-318K12.1 | 4.26 | 2.09 | 0.004607303 | 0.033020615 |
| ENSG00000118503 | TNFAIP3 | 4.23 | 2.08 | 3.01E-12 | 4.06E-10 |
| ENSG00000099958 | DERL3 | 4.23 | 2.08 | 9.02E-08 | 4.60E-06 |
| ENSG00000235522 | AC009505.2 | 4.23 | 2.08 | 2.13E-06 | 7.25E-05 |
| ENSG00000258181 | RP11-493L12.4 | 4.23 | 2.08 | 0.001689087 | 0.015562938 |
| ENSG00000120937 | NPPB | 4.23 | 2.08 | 0.002686561 | 0.022224307 |
| ENSG00000123338 | NCKAP1L | 4.22 | 2.08 | 3.68E-10 | 3.33E-08 |
| ENSG00000230606 | AC159540.1 | 4.22 | 2.08 | 2.85E-06 | 9.26E-05 |
| ENSG00000233264 | AC006042.8 | 4.22 | 2.08 | 6.24E-06 | 0.000179712 |
| ENSG00000259984 | RP11-335G20.7 | 4.22 | 2.08 | 0.002803056 | 0.022987746 |
| ENSG00000120280 | CXorf21 | 4.21 | 2.07 | 1.79E-06 | 6.17E-05 |
| ENSG00000172901 | AQPEP | 4.21 | 2.07 | 9.77E-06 | 0.000262986 |
| ENSG00000197262 | CCL4L2 | 4.21 | 2.08 | 0.005629814 | 0.03826609 |
| ENSG00000244681 | MTHFD2P1 | 4.19 | 2.07 | 0.000308478 | 0.004316309 |
| ENSG00000176049 | JAKMIP2 | 4.17 | 2.06 | 3.11E-08 | 1.79E-06 |
| ENSG00000105851 | PIK3CG | 4.15 | 2.05 | 2.21E-13 | 3.70E-11 |
| ENSG00000239713 | APOBEC3G | 4.15 | 2.05 | 7.15E-10 | 6.07E-08 |
| ENSG00000091490 | SEL1L3 | 4.15 | 2.05 | 7.67E-10 | 6.49E-08 |
| ENSG00000260249 | RP11-401P9.5 | 4.15 | 2.05 | 0.001169873 | 0.011818017 |
| ENSG00000157514 | TSC22D3 | 4.14 | 2.05 | 9.89E-11 | 9.86E-09 |
| ENSG00000187151 | ANGPTL5 | 4.14 | 2.05 | 1.45E-06 | 5.10E-05 |
| ENSG00000139144 | PIK3C2G | 4.14 | 2.05 | 0.000734533 | 0.008289995 |
| ENSG00000004948 | CALCR | 4.13 | 2.05 | 0.000429895 | 0.005555991 |
| ENSG00000132182 | NUP210 | 4.12 | 2.04 | 1.32E-08 | 8.25E-07 |
| ENSG00000240767 | RN7SL288P | 4.12 | 2.04 | 0.001639051 | 0.015214633 |
| ENSG00000255141 | HNRNPA1P76 | 4.11 | 2.04 | 3.73E-05 | 0.00078036 |
| ENSG00000231799 | RP13-93L13.2 | 4.1 | 2.04 | 4.57E-05 | 0.000925908 |
| ENSG00000229899 | AC084290.2 | 4.1 | 2.04 | 0.002953486 | 0.023897019 |
| ENSG00000265206 | MIR142 | 4.09 | 2.03 | 1.10E-06 | 4.05E-05 |
| ENSG00000231734 | RP6-206I17.2 | 4.09 | 2.03 | 0.003414964 | 0.026624101 |
| ENSG00000186517 | ARHGAP30 | 4.08 | 2.03 | 5.03E-11 | 5.24E-09 |
| ENSG00000171596 | NMUR1 | 4.07 | 2.02 | 2.10E-05 | 0.000485366 |
| ENSG00000127585 | FBXL16 | 4.05 | 2.02 | 6.07E-07 | 2.43E-05 |
| ENSG00000227155 | RP11-165F24.3 | 4.05 | 2.02 | 6.36E-07 | 2.52E-05 |
| ENSG00000258875 | CTD-2547L24.3 | 4.05 | 2.02 | 0.000108863 | 0.001876995 |
| ENSG00000240710 | RP11-430C7.4 | 4.05 | 2.02 | 0.000110468 | 0.001902534 |
| ENSG00000250771 | RP11-153M7.3 | 4.05 | 2.02 | 0.000257076 | 0.00373797 |
| ENSG00000183150 | GPR19 | 4.05 | 2.02 | 0.001431086 | 0.013757522 |
| ENSG00000154227 | CERS3 | 4.05 | 2.02 | 0.002315412 | 0.019866374 |
| ENSG00000118492 | ADGB | 4.04 | 2.01 | 0.003777855 | 0.028586086 |
| ENSG00000229647 | AC007879.7 | 4.04 | 2.01 | 0.005726279 | 0.038665875 |
| ENSG00000188404 | SELL | 4.03 | 2.01 | 2.82E-06 | 9.18E-05 |
| ENSG00000196126 | HLA-DRB1 | 4.02 | 2.01 | 1.33E-10 | 1.29E-08 |
| ENSG00000118432 | CNR1 | 4.02 | 2.01 | 9.91E-06 | 0.00026633 |
| ENSG00000183734 | ASCL2 | 4.02 | 2.01 | 0.000475736 | 0.006027094 |
| ENSG00000204472 | AIF1 | 4.01 | 2.00 | 1.18E-07 | 5.81E-06 |
| ENSG00000232487 | RASA3-IT1 | 4.01 | 2.00 | 0.004357645 | 0.031738063 |
| ENSG00000122872 | ARL4P | 4.01 | 2.00 | 0.00456735 | 0.03284122 |
| ENSG00000214212 | C19orf38 | 4 | 2.00 | 1.19E-05 | 0.000307374 |
| ENSG00000170396 | ZNF804A | 4 | 2.00 | 0.000609996 | 0.007216849 |
| ENSG00000243710 | WDR65 | 3.99 | 2.00 | 0.00023967 | 0.003533242 |
| ENSG00000180061 | TMEM150B | 3.98 | 1.99 | 2.49E-08 | 1.47E-06 |
| ENSG00000255587 | RAB44 | 3.98 | 1.99 | 0.000744852 | 0.008391091 |
| ENSG00000116299 | KIAA1324 | 3.97 | 1.99 | 9.96E-06 | 0.000266909 |
| ENSG00000236499 | LINC00896 | 3.97 | 1.99 | 0.000104457 | 0.001820381 |
| ENSG00000255163 | HSPE1P18 | 3.97 | 1.99 | 0.000531202 | 0.006544369 |
| ENSG00000233858 | AC026904.1 | 3.96 | 1.99 | 8.86E-05 | 0.00159105 |
| ENSG00000260997 | RP4-647J21.1 | 3.96 | 1.98 | 0.000130486 | 0.002182629 |
| ENSG00000000005 | TNMD | 3.95 | 1.98 | 0.001741739 | 0.015981158 |
| ENSG00000197629 | MPEG1 | 3.94 | 1.98 | 1.01E-11 | 1.24E-09 |
| ENSG00000204287 | HLA-DRA | 3.93 | 1.97 | 6.54E-09 | 4.42E-07 |
| ENSG00000169884 | WNT10B | 3.93 | 1.97 | 0.000282552 | 0.004019279 |
| ENSG00000255221 | CARD17 | 3.93 | 1.97 | 0.006143135 | 0.040737416 |
| ENSG00000166428 | PLD4 | 3.92 | 1.97 | 7.78E-06 | 0.000216331 |
| ENSG00000163131 | CTSS | 3.91 | 1.97 | 1.60E-08 | 9.84E-07 |
| ENSG00000165923 | AGBL2 | 3.91 | 1.97 | 0.00018843 | 0.002916766 |
| ENSG00000058335 | RASGRF1 | 3.9 | 1.96 | 0.001513567 | 0.014355282 |
| ENSG00000139055 | ERP27 | 3.89 | 1.96 | 5.11E-06 | 0.000151128 |
| ENSG00000272211 | RP11-347P5.1 | 3.88 | 1.96 | 3.94E-09 | 2.87E-07 |
| ENSG00000154096 | THY1 | 3.88 | 1.96 | 4.46E-06 | 0.000134182 |
| ENSG00000168447 | SCNN1B | 3.88 | 1.96 | 2.13E-05 | 0.000490508 |
| ENSG00000261996 | CTC-281F24.1 | 3.88 | 1.96 | 0.006429757 | 0.042085425 |
| ENSG00000109511 | ANXA10 | 3.87 | 1.95 | 0.000934083 | 0.009944729 |
| ENSG00000240474 | RN7SL116P | 3.87 | 1.95 | 0.002268853 | 0.019570447 |
| ENSG00000162777 | DENND2D | 3.86 | 1.95 | 5.62E-10 | 4.87E-08 |
| ENSG00000115956 | PLEK | 3.86 | 1.95 | 2.21E-06 | 7.46E-05 |
| ENSG00000182578 | CSF1R | 3.85 | 1.94 | 3.11E-15 | 6.71E-13 |
| ENSG00000242574 | HLA-DMB | 3.85 | 1.94 | 9.86E-11 | 9.86E-09 |
| ENSG00000134061 | CD180 | 3.85 | 1.95 | 9.39E-07 | 3.54E-05 |
| ENSG00000255733 | IFNG-AS1 | 3.85 | 1.95 | 4.00E-05 | 0.000825916 |
| ENSG00000100767 | PAPLN | 3.84 | 1.94 | 3.13E-09 | 2.31E-07 |
| ENSG00000254419 | RP11-261P9.4 | 3.84 | 1.94 | 1.97E-05 | 0.000460921 |
| ENSG00000112299 | VNN1 | 3.83 | 1.94 | 0.000106467 | 0.001844972 |
| ENSG00000232909 | RP3-510O8.4 | 3.83 | 1.94 | 0.005673141 | 0.038416516 |
| ENSG00000167094 | TTC16 | 3.82 | 1.93 | 1.10E-05 | 0.000290384 |
| ENSG00000256316 | HIST1H3F | 3.82 | 1.93 | 2.73E-05 | 0.000601605 |
| ENSG00000250651 | PABPC1P7 | 3.82 | 1.93 | 0.00015773 | 0.002541739 |
| ENSG00000171848 | RRM2 | 3.8 | 1.92 | 0.000756054 | 0.008495547 |
| ENSG00000272023 | CTC-350I8.1 | 3.8 | 1.93 | 0.005703991 | 0.038549171 |
| ENSG00000072818 | ACAP1 | 3.79 | 1.92 | 2.76E-14 | 5.07E-12 |
| ENSG00000228499 | TMSB10P1 | 3.79 | 1.92 | 0.005419678 | 0.037182298 |
| ENSG00000142347 | MYO1F | 3.78 | 1.92 | 4.93E-09 | 3.51E-07 |
| ENSG00000273447 | AC004067.5 | 3.78 | 1.92 | 2.81E-06 | 9.18E-05 |
| ENSG00000227531 | RP11-202G18.1 | 3.78 | 1.92 | 0.001211522 | 0.012139879 |
| ENSG00000028277 | POU2F2 | 3.76 | 1.91 | 2.22E-08 | 1.34E-06 |
| ENSG00000126759 | CFP | 3.76 | 1.91 | 9.19E-07 | 3.48E-05 |
| ENSG00000152932 | RAB3C | 3.76 | 1.91 | 0.001142571 | 0.011622239 |
| ENSG00000145703 | IQGAP2 | 3.75 | 1.91 | 3.20E-17 | 9.65E-15 |
| ENSG00000139194 | RBP5 | 3.75 | 1.91 | 1.46E-06 | 5.14E-05 |
| ENSG00000198010 | DLGAP2 | 3.75 | 1.91 | 0.003431569 | 0.026726513 |
| ENSG00000204482 | LST1 | 3.74 | 1.90 | 1.33E-05 | 0.000333554 |
| ENSG00000121281 | ADCY7 | 3.73 | 1.90 | 1.43E-14 | 2.75E-12 |
| ENSG00000143110 | C1orf162 | 3.72 | 1.90 | 2.68E-09 | 2.00E-07 |
| ENSG00000120899 | PTK2B | 3.71 | 1.89 | 8.53E-13 | 1.25E-10 |
| ENSG00000259792 | RP11-114H24.6 | 3.71 | 1.89 | 6.81E-05 | 0.001280697 |
| ENSG00000238266 | LINC00707 | 3.71 | 1.89 | 0.002068073 | 0.018310169 |
| ENSG00000229914 | RP11-404O13.4 | 3.71 | 1.89 | 0.003719122 | 0.028265875 |
| ENSG00000137841 | PLCB2 | 3.7 | 1.89 | 1.51E-15 | 3.47E-13 |
| ENSG00000181847 | TIGIT | 3.7 | 1.89 | 8.89E-06 | 0.000241407 |
| ENSG00000226091 | LINC00937 | 3.7 | 1.89 | 0.000229158 | 0.003407615 |
| ENSG00000268758 | EMR4P | 3.7 | 1.89 | 0.000992182 | 0.010398002 |
| ENSG00000159374 | M1AP | 3.7 | 1.89 | 0.001530566 | 0.014470165 |
| ENSG00000163606 | CD200R1 | 3.69 | 1.88 | 2.19E-06 | 7.43E-05 |
| ENSG00000162873 | KLHDC8A | 3.69 | 1.89 | 0.000625971 | 0.007369808 |
| ENSG00000162998 | FRZB | 3.68 | 1.88 | 9.61E-07 | 3.61E-05 |
| ENSG00000266017 | MIR4477A | 3.68 | 1.88 | 1.06E-05 | 0.000281557 |
| ENSG00000105352 | CEACAM4 | 3.68 | 1.88 | 2.49E-05 | 0.000556709 |
| ENSG00000256262 | USP30-AS1 | 3.68 | 1.88 | 0.000335647 | 0.004600368 |
| ENSG00000246375 | RP11-10L7.1 | 3.67 | 1.88 | 2.22E-06 | 7.47E-05 |
| ENSG00000137747 | TMPRSS13 | 3.67 | 1.88 | 5.15E-05 | 0.001020419 |
| ENSG00000012124 | CD22 | 3.67 | 1.87 | 7.86E-05 | 0.001445453 |
| ENSG00000133466 | C1QTNF6 | 3.66 | 1.87 | 2.36E-10 | 2.19E-08 |
| ENSG00000047365 | ARAP2 | 3.66 | 1.87 | 1.07E-09 | 8.86E-08 |
| ENSG00000161643 | SIGLEC16 | 3.66 | 1.87 | 1.22E-05 | 0.000313407 |
| ENSG00000258268 | RP11-570L15.1 | 3.66 | 1.87 | 2.32E-05 | 0.000526118 |
| ENSG00000135678 | CPM | 3.65 | 1.87 | 1.33E-08 | 8.25E-07 |
| ENSG00000168229 | PTGDR | 3.65 | 1.87 | 7.55E-06 | 0.000210834 |
| ENSG00000115415 | STAT1 | 3.64 | 1.86 | 2.81E-08 | 1.64E-06 |
| ENSG00000196189 | SEMA4A | 3.64 | 1.86 | 5.27E-07 | 2.13E-05 |
| ENSG00000240888 | RP13-635I23.3 | 3.63 | 1.86 | 5.20E-05 | 0.001026159 |
| ENSG00000258539 | RP11-12J10.3 | 3.63 | 1.86 | 9.61E-05 | 0.001699203 |
| ENSG00000213654 | GPSM3 | 3.62 | 1.85 | 9.17E-09 | 5.92E-07 |
| ENSG00000136490 | LIMD2 | 3.62 | 1.86 | 1.35E-08 | 8.32E-07 |
| ENSG00000149418 | ST14 | 3.62 | 1.86 | 1.46E-08 | 8.98E-07 |
| ENSG00000169413 | RNASE6 | 3.62 | 1.86 | 7.90E-07 | 3.05E-05 |
| ENSG00000102524 | TNFSF13B | 3.62 | 1.86 | 1.79E-06 | 6.17E-05 |
| ENSG00000030304 | MUSK | 3.61 | 1.85 | 1.20E-11 | 1.45E-09 |
| ENSG00000100100 | PIK3IP1 | 3.61 | 1.85 | 5.21E-09 | 3.67E-07 |
| ENSG00000231528 | FAM225A | 3.61 | 1.85 | 0.004562107 | 0.032818833 |
| ENSG00000107014 | RLN2 | 3.61 | 1.85 | 0.006018413 | 0.040067702 |
| ENSG00000237638 | AC007386.2 | 3.61 | 1.85 | 0.006797227 | 0.043817465 |
| ENSG00000171643 | S100Z | 3.61 | 1.85 | 0.007408722 | 0.046719225 |
| ENSG00000261270 | RP11-325K4.3 | 3.6 | 1.85 | 1.64E-07 | 7.64E-06 |
| ENSG00000264663 | KRT8P34 | 3.6 | 1.85 | 0.001618784 | 0.015086229 |
| ENSG00000225407 | CTD-2384B11.2 | 3.6 | 1.85 | 0.004938918 | 0.034802804 |
| ENSG00000170379 | FAM115C | 3.59 | 1.85 | 2.94E-08 | 1.69E-06 |
| ENSG00000162894 | FAIM3 | 3.59 | 1.84 | 3.28E-08 | 1.87E-06 |
| ENSG00000180539 | C9orf139 | 3.59 | 1.84 | 1.84E-05 | 0.00043718 |
| ENSG00000223491 | RP3-328E19.4 | 3.59 | 1.84 | 0.007454999 | 0.046934198 |
| ENSG00000072858 | SIDT1 | 3.58 | 1.84 | 1.28E-11 | 1.54E-09 |
| ENSG00000173372 | C1QA | 3.58 | 1.84 | 2.82E-08 | 1.64E-06 |
| ENSG00000136286 | MYO1G | 3.58 | 1.84 | 1.46E-06 | 5.14E-05 |
| ENSG00000248268 | CTC-499J9.1 | 3.58 | 1.84 | 0.000631884 | 0.007407075 |
| ENSG00000216802 | RP11-390P2.2 | 3.57 | 1.84 | 2.29E-05 | 0.000519625 |
| ENSG00000197459 | HIST1H2BH | 3.57 | 1.84 | 0.000360916 | 0.004868736 |
| ENSG00000163154 | TNFAIP8L2 | 3.55 | 1.83 | 2.44E-06 | 8.13E-05 |
| ENSG00000264773 | MIR4420 | 3.55 | 1.83 | 0.0069505 | 0.044540738 |
| ENSG00000256817 | TPT1P12 | 3.54 | 1.83 | 0.000564007 | 0.006806743 |
| ENSG00000155307 | SAMSN1 | 3.53 | 1.82 | 1.15E-06 | 4.20E-05 |
| ENSG00000267074 | RP11-1094M14.5 | 3.53 | 1.82 | 0.000204969 | 0.003106401 |
| ENSG00000172460 | PRSS30P | 3.52 | 1.82 | 0.00185486 | 0.016832728 |
| ENSG00000186854 | TRABD2A | 3.51 | 1.81 | 1.44E-07 | 6.94E-06 |
| ENSG00000151651 | ADAM8 | 3.51 | 1.81 | 3.50E-06 | 0.000110421 |
| ENSG00000149488 | TMC2 | 3.51 | 1.81 | 0.004437453 | 0.032169968 |
| ENSG00000081320 | STK17B | 3.5 | 1.81 | 2.03E-12 | 2.81E-10 |
| ENSG00000167483 | FAM129C | 3.5 | 1.81 | 0.002125352 | 0.018683091 |
| ENSG00000243811 | APOBEC3D | 3.49 | 1.80 | 1.29E-09 | 1.04E-07 |
| ENSG00000081985 | IL12RB2 | 3.49 | 1.80 | 1.30E-06 | 4.64E-05 |
| ENSG00000026297 | RNASET2 | 3.48 | 1.80 | 1.39E-09 | 1.10E-07 |
| ENSG00000128606 | LRRC17 | 3.48 | 1.80 | 2.80E-06 | 9.14E-05 |
| ENSG00000115607 | IL18RAP | 3.48 | 1.80 | 2.25E-05 | 0.00051281 |
| ENSG00000270127 | RP11-526I2.5 | 3.47 | 1.79 | 0.000291839 | 0.004117143 |
| ENSG00000240210 | RP11-204K16.1 | 3.47 | 1.79 | 0.004551406 | 0.032764809 |
| ENSG00000109674 | NEIL3 | 3.46 | 1.79 | 0.003492437 | 0.027050148 |
| ENSG00000239465 | RP11-330L19.2 | 3.46 | 1.79 | 0.004065188 | 0.030152264 |
| ENSG00000234996 | RP11-480I12.7 | 3.46 | 1.79 | 0.007207139 | 0.045775953 |
| ENSG00000248466 | RP11-640B6.1 | 3.45 | 1.79 | 0.000189549 | 0.002925954 |
| ENSG00000261471 | RP11-61F12.1 | 3.45 | 1.79 | 0.000543379 | 0.006651586 |
| ENSG00000249721 | RP11-83M16.4 | 3.45 | 1.79 | 0.002558795 | 0.021435304 |
| ENSG00000229727 | AC013460.1 | 3.45 | 1.79 | 0.003533006 | 0.027295747 |
| ENSG00000162711 | NLRP3 | 3.44 | 1.78 | 5.83E-10 | 5.03E-08 |
| ENSG00000172215 | CXCR6 | 3.44 | 1.78 | 4.73E-09 | 3.40E-07 |
| ENSG00000166825 | ANPEP | 3.44 | 1.78 | 6.96E-07 | 2.74E-05 |
| ENSG00000203497 | PDCD4-AS1 | 3.43 | 1.78 | 1.28E-06 | 4.57E-05 |
| ENSG00000047457 | CP | 3.43 | 1.78 | 1.54E-06 | 5.39E-05 |
| ENSG00000272825 | LL21NC02-1C16.2 | 3.43 | 1.78 | 0.0018351 | 0.016673918 |
| ENSG00000205663 | RP11-706O15.5 | 3.43 | 1.78 | 0.00218098 | 0.019014764 |
| ENSG00000136040 | PLXNC1 | 3.42 | 1.77 | 6.99E-14 | 1.23E-11 |
| ENSG00000120262 | CCDC170 | 3.42 | 1.77 | 1.91E-07 | 8.76E-06 |
| ENSG00000235568 | NFAM1 | 3.42 | 1.78 | 2.99E-07 | 1.29E-05 |
| ENSG00000228784 | LINC00954 | 3.42 | 1.77 | 5.06E-06 | 0.00014978 |
| ENSG00000197168 | NEK5 | 3.41 | 1.77 | 1.51E-06 | 5.30E-05 |
| ENSG00000269345 | VN1R85P | 3.41 | 1.77 | 4.13E-05 | 0.00084677 |
| ENSG00000270723 | RP11-401N16.1 | 3.41 | 1.77 | 0.00692344 | 0.044413513 |
| ENSG00000019582 | CD74 | 3.4 | 1.76 | 5.47E-09 | 3.82E-07 |
| ENSG00000139187 | KLRG1 | 3.4 | 1.77 | 4.41E-07 | 1.82E-05 |
| ENSG00000161944 | ASGR2 | 3.4 | 1.77 | 0.00026984 | 0.003886887 |
| ENSG00000221743 | Z95152.1 | 3.4 | 1.76 | 0.003825707 | 0.028876494 |
| ENSG00000074370 | ATP2A3 | 3.39 | 1.76 | 4.38E-10 | 3.88E-08 |
| ENSG00000168016 | TRANK1 | 3.39 | 1.76 | 3.76E-08 | 2.10E-06 |
| ENSG00000196668 | LINC00173 | 3.39 | 1.76 | 0.000142695 | 0.00234734 |
| ENSG00000231633 | LINC00283 | 3.39 | 1.76 | 0.00018853 | 0.002916766 |
| ENSG00000128383 | APOBEC3A | 3.39 | 1.76 | 0.000196658 | 0.00301155 |
| ENSG00000026950 | BTN3A1 | 3.38 | 1.76 | 1.19E-10 | 1.17E-08 |
| ENSG00000154188 | ANGPT1 | 3.38 | 1.76 | 2.63E-07 | 1.15E-05 |
| ENSG00000163013 | FBXO41 | 3.38 | 1.76 | 1.41E-06 | 4.98E-05 |
| ENSG00000169403 | PTAFR | 3.37 | 1.75 | 3.79E-07 | 1.60E-05 |
| ENSG00000228956 | AC144521.1 | 3.37 | 1.75 | 5.91E-05 | 0.001147384 |
| ENSG00000228403 | RP11-563N6.6 | 3.37 | 1.75 | 0.001431968 | 0.013757522 |
| ENSG00000101938 | CHRDL1 | 3.36 | 1.75 | 2.56E-05 | 0.000570257 |
| ENSG00000203815 | FAM231D | 3.36 | 1.75 | 8.43E-05 | 0.001535917 |
| ENSG00000228548 | ITPKB-AS1 | 3.36 | 1.75 | 0.000575338 | 0.006911008 |
| ENSG00000236320 | SLFN14 | 3.36 | 1.75 | 0.000758203 | 0.008507278 |
| ENSG00000103187 | COTL1 | 3.35 | 1.74 | 2.16E-06 | 7.35E-05 |
| ENSG00000085514 | PILRA | 3.35 | 1.74 | 4.27E-06 | 0.000129881 |
| ENSG00000234883 | MIR155HG | 3.35 | 1.75 | 0.000118783 | 0.00201989 |
| ENSG00000185527 | PDE6G | 3.34 | 1.74 | 0.003825297 | 0.028876494 |
| ENSG00000267474 | CTC-548K16.6 | 3.34 | 1.74 | 0.004401862 | 0.031987211 |
| ENSG00000197705 | KLHL14 | 3.33 | 1.74 | 0.001181143 | 0.011904554 |
| ENSG00000131355 | EMR3 | 3.33 | 1.73 | 0.005747056 | 0.03875521 |
| ENSG00000231389 | HLA-DPA1 | 3.32 | 1.73 | 3.51E-07 | 1.49E-05 |
| ENSG00000150782 | IL18 | 3.32 | 1.73 | 4.25E-07 | 1.77E-05 |
| ENSG00000169313 | P2RY12 | 3.32 | 1.73 | 4.37E-06 | 0.000132516 |
| ENSG00000231752 | EMBP1 | 3.32 | 1.73 | 0.000261958 | 0.003796418 |
| ENSG00000049768 | FOXP3 | 3.32 | 1.73 | 0.000529385 | 0.006532196 |
| ENSG00000255189 | GLYATL1P1 | 3.32 | 1.73 | 0.007490415 | 0.047099421 |
| ENSG00000196730 | DAPK1 | 3.31 | 1.72 | 2.19E-09 | 1.66E-07 |
| ENSG00000127951 | FGL2 | 3.31 | 1.73 | 2.34E-09 | 1.77E-07 |
| ENSG00000111679 | PTPN6 | 3.3 | 1.72 | 4.26E-08 | 2.35E-06 |
| ENSG00000235777 | DPYD-AS2 | 3.3 | 1.72 | 3.12E-06 | 0.000100067 |
| ENSG00000240498 | CDKN2B-AS1 | 3.3 | 1.72 | 0.000548393 | 0.006686397 |
| ENSG00000229124 | VIM-AS1 | 3.29 | 1.72 | 5.37E-06 | 0.000157696 |
| ENSG00000254254 | RP11-17A4.2 | 3.29 | 1.72 | 0.00070894 | 0.00809906 |
| ENSG00000229870 | RP11-507K13.6 | 3.29 | 1.72 | 0.004651453 | 0.033275113 |
| ENSG00000102854 | MSLN | 3.29 | 1.72 | 0.006525664 | 0.042534849 |
| ENSG00000156711 | MAPK13 | 3.28 | 1.71 | 9.73E-07 | 3.64E-05 |
| ENSG00000108932 | SLC16A6 | 3.28 | 1.72 | 0.000102791 | 0.001795401 |
| ENSG00000130812 | ANGPTL6 | 3.28 | 1.72 | 0.000862289 | 0.009360921 |
| ENSG00000065413 | ANKRD44 | 3.27 | 1.71 | 1.58E-12 | 2.22E-10 |
| ENSG00000230188 | RP11-405L18.4 | 3.27 | 1.71 | 0.001666001 | 0.015410133 |
| ENSG00000178015 | GPR150 | 3.27 | 1.71 | 0.006707597 | 0.043408482 |
| ENSG00000237424 | FOXD2-AS1 | 3.26 | 1.71 | 8.75E-05 | 0.001580097 |
| ENSG00000197249 | SERPINA1 | 3.26 | 1.70 | 0.000785669 | 0.008720194 |
| ENSG00000042980 | ADAM28 | 3.25 | 1.70 | 1.21E-10 | 1.19E-08 |
| ENSG00000140090 | SLC24A4 | 3.25 | 1.70 | 2.32E-05 | 0.000526694 |
| ENSG00000267096 | CTD-2537I9.13 | 3.25 | 1.70 | 0.005027116 | 0.035212265 |
| ENSG00000187764 | SEMA4D | 3.24 | 1.70 | 1.90E-10 | 1.79E-08 |
| ENSG00000108846 | ABCC3 | 3.24 | 1.69 | 7.32E-07 | 2.86E-05 |
| ENSG00000198829 | SUCNR1 | 3.24 | 1.69 | 5.17E-05 | 0.001022476 |
| ENSG00000223668 | EEF1A1P24 | 3.24 | 1.70 | 0.007096006 | 0.045256685 |
| ENSG00000165025 | SYK | 3.23 | 1.69 | 4.01E-11 | 4.22E-09 |
| ENSG00000230537 | RP11-305L7.1 | 3.22 | 1.69 | 0.00037987 | 0.005071127 |
| ENSG00000156466 | GDF6 | 3.22 | 1.69 | 0.000484649 | 0.006102384 |
| ENSG00000233968 | RP11-354E11.2 | 3.22 | 1.69 | 0.003451086 | 0.026858147 |
| ENSG00000174607 | UGT8 | 3.22 | 1.69 | 0.004183356 | 0.030776836 |
| ENSG00000129673 | AANAT | 3.21 | 1.68 | 2.86E-05 | 0.000625515 |
| ENSG00000089639 | GMIP | 3.2 | 1.68 | 9.20E-08 | 4.65E-06 |
| ENSG00000146530 | VWDE | 3.2 | 1.68 | 0.006804926 | 0.043817465 |
| ENSG00000196747 | HIST1H2AI | 3.19 | 1.67 | 0.000686512 | 0.007916218 |
| ENSG00000076944 | STXBP2 | 3.18 | 1.67 | 2.75E-05 | 0.000604424 |
| ENSG00000112414 | GPR126 | 3.17 | 1.66 | 1.01E-10 | 1.00E-08 |
| ENSG00000183748 | MRC1L1 | 3.17 | 1.66 | 2.83E-08 | 1.64E-06 |
| ENSG00000173369 | C1QB | 3.17 | 1.66 | 9.48E-06 | 0.000256348 |
| ENSG00000142185 | TRPM2 | 3.16 | 1.66 | 4.87E-09 | 3.49E-07 |
| ENSG00000186310 | NAP1L3 | 3.15 | 1.65 | 3.07E-05 | 0.000662549 |
| ENSG00000000938 | FGR | 3.15 | 1.66 | 6.70E-05 | 0.001263883 |
| ENSG00000184956 | MUC6 | 3.15 | 1.66 | 0.000129517 | 0.00216996 |
| ENSG00000006075 | CCL3 | 3.15 | 1.66 | 0.002814468 | 0.023069057 |
| ENSG00000153446 | C16orf89 | 3.14 | 1.65 | 9.63E-05 | 0.001701849 |
| ENSG00000142765 | SYTL1 | 3.13 | 1.65 | 7.88E-10 | 6.65E-08 |
| ENSG00000177602 | GSG2 | 3.13 | 1.65 | 5.61E-06 | 0.000164031 |
| ENSG00000254923 | RP11-1236K1.8 | 3.13 | 1.65 | 0.001563338 | 0.014680896 |
| ENSG00000235880 | RP11-59O6.3 | 3.13 | 1.65 | 0.006616871 | 0.04296671 |
| ENSG00000172345 | STARD5 | 3.12 | 1.64 | 1.79E-10 | 1.70E-08 |
| ENSG00000175567 | UCP2 | 3.12 | 1.64 | 4.18E-07 | 1.74E-05 |
| ENSG00000125384 | PTGER2 | 3.12 | 1.64 | 0.000319608 | 0.004443798 |
| ENSG00000229417 | NPM1P25 | 3.12 | 1.64 | 0.003682948 | 0.028053232 |
| ENSG00000074410 | CA12 | 3.12 | 1.64 | 0.00488144 | 0.034520622 |
| ENSG00000049089 | COL9A2 | 3.11 | 1.64 | 3.27E-07 | 1.40E-05 |
| ENSG00000055732 | MCOLN3 | 3.11 | 1.64 | 1.11E-06 | 4.05E-05 |
| ENSG00000168350 | DEGS2 | 3.11 | 1.64 | 0.000410007 | 0.005373336 |
| ENSG00000239264 | TXNDC5 | 3.1 | 1.63 | 1.40E-06 | 4.93E-05 |
| ENSG00000258926 | RP11-47I22.1 | 3.1 | 1.63 | 2.95E-06 | 9.53E-05 |
| ENSG00000152582 | SPEF2 | 3.1 | 1.63 | 1.34E-05 | 0.000335568 |
| ENSG00000248791 | CTD-2165H16.3 | 3.1 | 1.63 | 0.00152832 | 0.014453367 |
| ENSG00000254750 | CASP1P2 | 3.1 | 1.63 | 0.002012689 | 0.017908029 |
| ENSG00000144331 | ZNF385B | 3.1 | 1.63 | 0.002801659 | 0.022982405 |
| ENSG00000165457 | FOLR2 | 3.09 | 1.63 | 2.82E-08 | 1.64E-06 |
| ENSG00000266378 | RP11-214O1.3 | 3.09 | 1.63 | 0.001264979 | 0.012544346 |
| ENSG00000241163 | LINC00877 | 3.09 | 1.63 | 0.003084567 | 0.024702774 |
| ENSG00000148908 | RGS10 | 3.08 | 1.62 | 1.63E-07 | 7.64E-06 |
| ENSG00000143466 | IKBKE | 3.08 | 1.62 | 1.10E-05 | 0.000290384 |
| ENSG00000188761 | BCL2L15 | 3.08 | 1.62 | 0.000791899 | 0.008767822 |
| ENSG00000219249 | AMZ2P2 | 3.08 | 1.62 | 0.002525127 | 0.021214885 |
| ENSG00000187994 | RINL | 3.07 | 1.62 | 5.57E-10 | 4.84E-08 |
| ENSG00000114013 | CD86 | 3.07 | 1.62 | 3.47E-05 | 0.000736746 |
| ENSG00000165449 | SLC16A9 | 3.07 | 1.62 | 0.000185579 | 0.002880545 |
| ENSG00000198374 | HIST1H2AL | 3.07 | 1.62 | 0.000186931 | 0.002900063 |
| ENSG00000173715 | C11orf80 | 3.06 | 1.61 | 2.83E-08 | 1.64E-06 |
| ENSG00000186818 | LILRB4 | 3.06 | 1.61 | 1.29E-05 | 0.000326993 |
| ENSG00000140675 | SLC5A2 | 3.06 | 1.61 | 0.000144371 | 0.002367334 |
| ENSG00000125735 | TNFSF14 | 3.06 | 1.62 | 0.000223291 | 0.003334855 |
| ENSG00000166527 | CLEC4D | 3.06 | 1.61 | 0.000523241 | 0.006474535 |
| ENSG00000104081 | BMF | 3.05 | 1.61 | 4.58E-10 | 4.04E-08 |
| ENSG00000225217 | HSPA7 | 3.05 | 1.61 | 8.64E-09 | 5.63E-07 |
| ENSG00000151575 | TEX9 | 3.05 | 1.61 | 1.34E-07 | 6.55E-06 |
| ENSG00000201957 | SNORA25 | 3.05 | 1.61 | 0.000372904 | 0.004999258 |
| ENSG00000255836 | RP11-157G21.2 | 3.05 | 1.61 | 0.000570325 | 0.006861491 |
| ENSG00000272668 | RP11-190A12.8 | 3.05 | 1.61 | 0.002830339 | 0.023186807 |
| ENSG00000259513 | CYCSP38 | 3.05 | 1.61 | 0.003763406 | 0.028504008 |
| ENSG00000206965 | RNU6-5P | 3.05 | 1.61 | 0.005504571 | 0.037642709 |
| ENSG00000174130 | TLR6 | 3.04 | 1.60 | 1.67E-09 | 1.30E-07 |
| ENSG00000121380 | BCL2L14 | 3.04 | 1.61 | 0.001645565 | 0.015257747 |
| ENSG00000015133 | CCDC88C | 3.03 | 1.60 | 1.67E-09 | 1.30E-07 |
| ENSG00000168918 | INPP5D | 3.03 | 1.60 | 6.47E-09 | 4.38E-07 |
| ENSG00000186470 | BTN3A2 | 3.03 | 1.60 | 7.74E-08 | 4.03E-06 |
| ENSG00000155465 | SLC7A7 | 3.03 | 1.60 | 1.05E-06 | 3.89E-05 |
| ENSG00000270659 | RP11-105N14.1 | 3.03 | 1.60 | 0.001663471 | 0.015400522 |
| ENSG00000200735 | RNY4P8 | 3.03 | 1.60 | 0.007036825 | 0.044935052 |
| ENSG00000130755 | GMFG | 3.02 | 1.59 | 2.87E-08 | 1.66E-06 |
| ENSG00000141968 | VAV1 | 3.02 | 1.59 | 8.27E-06 | 0.000227394 |
| ENSG00000271538 | RP11-326I11.4 | 3.02 | 1.60 | 0.000518005 | 0.006430403 |
| ENSG00000167613 | LAIR1 | 3.01 | 1.59 | 1.79E-07 | 8.28E-06 |
| ENSG00000117600 | LPPR4 | 3.01 | 1.59 | 6.93E-06 | 0.000196326 |
| ENSG00000238005 | RP11-443B7.1 | 3.01 | 1.59 | 0.004928402 | 0.034771925 |
| ENSG00000130775 | THEMIS2 | 3 | 1.58 | 7.05E-07 | 2.77E-05 |
| ENSG00000122420 | PTGFR | 3 | 1.59 | 7.75E-06 | 0.000215795 |
| ENSG00000257221 | RP11-689B22.2 | 3 | 1.59 | 0.000158813 | 0.00255252 |
| ENSG00000121898 | CPXM2 | 3 | 1.59 | 0.000179286 | 0.002806902 |
| ENSG00000256720 | RP11-436I9.6 | 3 | 1.58 | 0.000745917 | 0.008400018 |
| ENSG00000235532 | LINC00402 | 3 | 1.58 | 0.002408509 | 0.020460042 |
| ENSG00000204257 | HLA-DMA | 2.99 | 1.58 | 5.61E-09 | 3.88E-07 |
| ENSG00000158050 | DUSP2 | 2.99 | 1.58 | 2.78E-06 | 9.10E-05 |
| ENSG00000223946 | RP11-533O20.2 | 2.99 | 1.58 | 0.001543853 | 0.014564501 |
| ENSG00000179715 | PCED1B | 2.98 | 1.57 | 2.36E-06 | 7.90E-05 |
| ENSG00000027869 | SH2D2A | 2.97 | 1.57 | 5.81E-06 | 0.000168314 |
| ENSG00000226237 | RP11-276H19.1 | 2.97 | 1.57 | 0.000982087 | 0.010313806 |
| ENSG00000185155 | MIXL1 | 2.97 | 1.57 | 0.003605808 | 0.027644442 |
| ENSG00000064012 | CASP8 | 2.96 | 1.57 | 1.79E-09 | 1.38E-07 |
| ENSG00000235999 | RP11-403I13.8 | 2.96 | 1.56 | 1.23E-05 | 0.000313407 |
| ENSG00000148204 | CRB2 | 2.96 | 1.57 | 0.000192954 | 0.002966621 |
| ENSG00000100678 | SLC8A3 | 2.96 | 1.57 | 0.002416645 | 0.020500894 |
| ENSG00000130768 | SMPDL3B | 2.96 | 1.57 | 0.003242096 | 0.025646271 |
| ENSG00000011590 | ZBTB32 | 2.95 | 1.56 | 1.05E-07 | 5.21E-06 |
| ENSG00000115339 | GALNT3 | 2.95 | 1.56 | 1.46E-05 | 0.000361366 |
| ENSG00000104783 | KCNN4 | 2.95 | 1.56 | 0.00012918 | 0.002167076 |
| ENSG00000227218 | RP11-203J24.8 | 2.95 | 1.56 | 0.000330961 | 0.004552343 |
| ENSG00000267312 | RP11-1094M14.7 | 2.95 | 1.56 | 0.002600053 | 0.021668985 |
| ENSG00000266389 | CTB-41I6.1 | 2.95 | 1.56 | 0.003679285 | 0.028044633 |
| ENSG00000271913 | RP1-111C20.4 | 2.94 | 1.56 | 1.54E-09 | 1.21E-07 |
| ENSG00000173198 | CYSLTR1 | 2.94 | 1.56 | 1.21E-08 | 7.58E-07 |
| ENSG00000230322 | RP3-323N1.2 | 2.94 | 1.56 | 0.000136059 | 0.002257468 |
| ENSG00000135114 | OASL | 2.94 | 1.55 | 0.000391369 | 0.005190909 |
| ENSG00000080224 | EPHA6 | 2.94 | 1.55 | 0.000408209 | 0.005363469 |
| ENSG00000238528 | snoU13 | 2.94 | 1.55 | 0.00054103 | 0.006635877 |
| ENSG00000140678 | ITGAX | 2.94 | 1.56 | 0.001195603 | 0.012022754 |
| ENSG00000143196 | DPT | 2.93 | 1.55 | 7.04E-09 | 4.69E-07 |
| ENSG00000261971 | RP11-473M20.7 | 2.93 | 1.55 | 1.95E-05 | 0.000457402 |
| ENSG00000214851 | LINC00612 | 2.92 | 1.55 | 9.30E-05 | 0.001657195 |
| ENSG00000259236 | CTD-2611K5.5 | 2.92 | 1.55 | 0.00052161 | 0.00645953 |
| ENSG00000271151 | RP11-394I13.2 | 2.92 | 1.55 | 0.001867513 | 0.016913563 |
| ENSG00000179083 | FAM133A | 2.92 | 1.55 | 0.003162507 | 0.025171794 |
| ENSG00000128604 | IRF5 | 2.91 | 1.54 | 2.25E-08 | 1.35E-06 |
| ENSG00000138439 | FAM117B | 2.91 | 1.54 | 1.29E-07 | 6.30E-06 |
| ENSG00000088827 | SIGLEC1 | 2.91 | 1.54 | 2.21E-06 | 7.44E-05 |
| ENSG00000198417 | MT1F | 2.91 | 1.54 | 3.02E-06 | 9.70E-05 |
| ENSG00000165168 | CYBB | 2.91 | 1.54 | 3.04E-05 | 0.00065771 |
| ENSG00000242651 | RN7SL862P | 2.91 | 1.54 | 0.005141951 | 0.035706255 |
| ENSG00000176971 | FIBIN | 2.9 | 1.54 | 5.43E-05 | 0.001065282 |
| ENSG00000005381 | MPO | 2.9 | 1.54 | 0.001019624 | 0.01062775 |
| ENSG00000151023 | ENKUR | 2.89 | 1.53 | 0.003260966 | 0.025775686 |
| ENSG00000095970 | TREM2 | 2.88 | 1.53 | 9.80E-05 | 0.001724083 |
| ENSG00000225496 | AC104651.2 | 2.88 | 1.53 | 0.001477746 | 0.01412247 |
| ENSG00000214894 | LINC00243 | 2.88 | 1.53 | 0.002903852 | 0.02359247 |
| ENSG00000125144 | MT1G | 2.88 | 1.53 | 0.006322328 | 0.041587797 |
| ENSG00000110079 | MS4A4A | 2.87 | 1.52 | 4.75E-06 | 0.000141614 |
| ENSG00000258810 | RP11-219E7.1 | 2.87 | 1.52 | 0.000224924 | 0.003354367 |
| ENSG00000136449 | MYCBPAP | 2.87 | 1.52 | 0.000401842 | 0.005297889 |
| ENSG00000100629 | CEP128 | 2.86 | 1.52 | 2.54E-09 | 1.91E-07 |
| ENSG00000180549 | FUT7 | 2.86 | 1.52 | 0.00166354 | 0.015400522 |
| ENSG00000086288 | NME8 | 2.86 | 1.51 | 0.00211137 | 0.018602636 |
| ENSG00000244682 | FCGR2C | 2.85 | 1.51 | 1.26E-09 | 1.02E-07 |
| ENSG00000159189 | C1QC | 2.85 | 1.51 | 7.50E-06 | 0.000209778 |
| ENSG00000105383 | CD33 | 2.85 | 1.51 | 1.67E-05 | 0.000404423 |
| ENSG00000254362 | RP11-14I17.3 | 2.85 | 1.51 | 0.001537191 | 0.014523887 |
| ENSG00000198865 | CCDC152 | 2.84 | 1.51 | 1.54E-09 | 1.21E-07 |
| ENSG00000059377 | TBXAS1 | 2.84 | 1.51 | 5.99E-09 | 4.08E-07 |
| ENSG00000178075 | GRAMD1C | 2.84 | 1.51 | 3.21E-05 | 0.000687341 |
| ENSG00000104974 | LILRA1 | 2.84 | 1.51 | 0.001306774 | 0.012851261 |
| ENSG00000114268 | PFKFB4 | 2.83 | 1.50 | 1.39E-07 | 6.73E-06 |
| ENSG00000254503 | CTD-2521M24.4 | 2.83 | 1.50 | 0.000611667 | 0.007229759 |
| ENSG00000106178 | CCL24 | 2.83 | 1.50 | 0.005302746 | 0.036550987 |
| ENSG00000163435 | ELF3 | 0 | -8.37 | 5.32E-18 | 1.90E-15 |
| ENSG00000248713 | RP11-766F14.2 | 0 | -8.66 | 3.22E-16 | 8.28E-14 |
| ENSG00000268864 | CTB-167G5.5 | 0 | -9.11 | 3.75E-09 | 2.75E-07 |
| ENSG00000215506 | TPTE2P4 | -0.03 | -5.20 | 3.54E-06 | 0.000111442 |
| ENSG00000123977 | DAW1 | -0.03 | -4.98 | 2.12E-05 | 0.000488664 |
| ENSG00000254510 | RP11-867G23.10 | -0.04 | -4.61 | 8.89E-09 | 5.76E-07 |
| ENSG00000127129 | EDN2 | -0.04 | -4.58 | 2.51E-08 | 1.48E-06 |
| ENSG00000234680 | VN2R3P | -0.04 | -4.83 | 6.60E-05 | 0.001250784 |
| ENSG00000197616 | MYH6 | -0.06 | -4.17 | 2.54E-15 | 5.64E-13 |
| ENSG00000119508 | NR4A3 | -0.06 | -4.17 | 7.64E-12 | 9.53E-10 |
| ENSG00000166592 | RRAD | -0.07 | -3.92 | 9.17E-19 | 3.72E-16 |
| ENSG00000007908 | SELE | -0.07 | -3.94 | 4.27E-06 | 0.000129882 |
| ENSG00000161798 | AQP5 | -0.09 | -3.48 | 4.05E-06 | 0.00012408 |
| ENSG00000101144 | BMP7 | -0.09 | -3.48 | 5.12E-06 | 0.000151128 |
| ENSG00000185352 | HS6ST3 | -0.09 | -3.45 | 0.000327284 | 0.004507801 |
| ENSG00000164588 | HCN1 | -0.09 | -3.48 | 0.000615912 | 0.007270826 |
| ENSG00000177984 | LCN15 | -0.1 | -3.29 | 0.000226085 | 0.00336842 |
| ENSG00000176919 | C8G | -0.11 | -3.17 | 1.26E-12 | 1.81E-10 |
| ENSG00000204655 | MOG | -0.11 | -3.15 | 1.19E-11 | 1.44E-09 |
| ENSG00000235994 | RP3-470B24.5 | -0.11 | -3.12 | 5.12E-06 | 0.000151128 |
| ENSG00000259342 | RP11-519G16.5 | -0.11 | -3.13 | 6.30E-05 | 0.001204941 |
| ENSG00000124875 | CXCL6 | -0.11 | -3.16 | 0.001874238 | 0.016959517 |
| ENSG00000263155 | MYZAP | -0.12 | -3.05 | 1.27E-14 | 2.44E-12 |
| ENSG00000164089 | ETNPPL | -0.12 | -3.04 | 2.72E-11 | 3.00E-09 |
| ENSG00000013588 | GPRC5A | -0.12 | -3.02 | 5.44E-07 | 2.20E-05 |
| ENSG00000174429 | ABRA | -0.12 | -3.03 | 1.76E-06 | 6.09E-05 |
| ENSG00000261635 | RP11-618N24.1 | -0.12 | -3.11 | 3.59E-06 | 0.000112873 |
| ENSG00000223695 | RP4-633O19__A.1 | -0.12 | -3.03 | 1.49E-05 | 0.000367039 |
| ENSG00000186335 | SLC36A2 | -0.12 | -3.03 | 4.53E-05 | 0.00091941 |
| ENSG00000224904 | RP5-934G17.6 | -0.12 | -3.05 | 0.000871115 | 0.009437386 |
| ENSG00000163273 | NPPC | -0.13 | -2.91 | 5.66E-06 | 0.000164744 |
| ENSG00000118849 | RARRES1 | -0.13 | -2.93 | 2.00E-05 | 0.000466763 |
| ENSG00000108691 | CCL2 | -0.13 | -2.95 | 4.48E-05 | 0.000912289 |
| ENSG00000184557 | SOCS3 | -0.13 | -2.92 | 6.76E-05 | 0.001272995 |
| ENSG00000229367 | HMGN2P19 | -0.13 | -2.91 | 0.001747315 | 0.016008483 |
| ENSG00000198216 | CACNA1E | -0.14 | -2.81 | 1.34E-06 | 4.75E-05 |
| ENSG00000261315 | LARP4P | -0.14 | -2.84 | 1.24E-05 | 0.000315406 |
| ENSG00000166091 | CMTM5 | -0.14 | -2.88 | 4.02E-05 | 0.000828672 |
| ENSG00000197991 | PCDH20 | -0.15 | -2.74 | 3.91E-13 | 6.21E-11 |
| ENSG00000168334 | XIRP1 | -0.15 | -2.75 | 1.51E-11 | 1.76E-09 |
| ENSG00000185112 | FAM43A | -0.15 | -2.73 | 1.52E-10 | 1.46E-08 |
| ENSG00000104848 | KCNA7 | -0.15 | -2.69 | 0.000466916 | 0.005932563 |
| ENSG00000171476 | HOPX | -0.16 | -2.66 | 1.40E-09 | 1.11E-07 |
| ENSG00000223414 | LINC00473 | -0.16 | -2.65 | 7.21E-09 | 4.79E-07 |
| ENSG00000261191 | RP11-16L14.2 | -0.16 | -2.68 | 8.84E-06 | 0.00024048 |
| ENSG00000260582 | TPST2P1 | -0.16 | -2.66 | 3.55E-05 | 0.00074894 |
| ENSG00000148702 | HABP2 | -0.16 | -2.64 | 4.75E-05 | 0.000953103 |
| ENSG00000054938 | CHRDL2 | -0.16 | -2.62 | 8.33E-05 | 0.00152152 |
| ENSG00000253603 | CTA-397H3.3 | -0.16 | -2.66 | 0.003416093 | 0.02662617 |
| ENSG00000130176 | CNN1 | -0.17 | -2.52 | 8.68E-09 | 5.64E-07 |
| ENSG00000102802 | MEDAG | -0.17 | -2.58 | 2.41E-08 | 1.43E-06 |
| ENSG00000137801 | THBS1 | -0.17 | -2.54 | 1.27E-06 | 4.57E-05 |
| ENSG00000162772 | ATF3 | -0.17 | -2.56 | 1.75E-05 | 0.000418662 |
| ENSG00000272899 | RP11-309L24.9 | -0.17 | -2.55 | 3.82E-05 | 0.000796016 |
| ENSG00000250061 | RP11-541P9.3 | -0.17 | -2.55 | 0.000155324 | 0.002512168 |
| ENSG00000273259 | RP11-986E7.7 | -0.17 | -2.53 | 0.000192761 | 0.002965127 |
| ENSG00000221299 | Z83826.1 | -0.17 | -2.53 | 0.000220105 | 0.003292059 |
| ENSG00000189350 | FAM179A | -0.18 | -2.49 | 3.42E-05 | 0.00072618 |
| ENSG00000269641 | CTB-167G5.6 | -0.18 | -2.45 | 0.002706371 | 0.02232557 |
| ENSG00000184845 | DRD1 | -0.18 | -2.45 | 0.004931209 | 0.034781352 |
| ENSG00000165175 | MID1IP1 | -0.19 | -2.37 | 7.88E-09 | 5.20E-07 |
| ENSG00000175505 | CLCF1 | -0.19 | -2.37 | 3.60E-06 | 0.000112968 |
| ENSG00000254449 | SF3A3P2 | -0.19 | -2.37 | 6.06E-05 | 0.001167609 |
| ENSG00000165621 | OXGR1 | -0.19 | -2.36 | 0.000476656 | 0.006036273 |
| ENSG00000234965 | SHISA8 | -0.19 | -2.39 | 0.003951624 | 0.029616512 |
| ENSG00000139438 | FAM222A | -0.2 | -2.32 | 3.81E-09 | 2.79E-07 |
| ENSG00000187479 | C11orf96 | -0.2 | -2.33 | 1.06E-08 | 6.77E-07 |
| ENSG00000244998 | CTD-3064M3.4 | -0.2 | -2.34 | 1.30E-08 | 8.11E-07 |
| ENSG00000231933 | CTA-125H2.2 | -0.2 | -2.32 | 3.98E-07 | 1.67E-05 |
| ENSG00000261054 | RP11-6O2.4 | -0.2 | -2.30 | 4.70E-06 | 0.000140055 |
| ENSG00000075886 | TUBA3D | -0.2 | -2.35 | 1.11E-05 | 0.000291202 |
| ENSG00000156966 | B3GNT7 | -0.2 | -2.34 | 1.29E-05 | 0.000325676 |
| ENSG00000253669 | KB-1732A1.1 | -0.2 | -2.33 | 3.51E-05 | 0.000743052 |
| ENSG00000124935 | SCGB1D2 | -0.2 | -2.33 | 0.001291092 | 0.012729548 |
| ENSG00000123358 | NR4A1 | -0.21 | -2.25 | 2.04E-07 | 9.24E-06 |
| ENSG00000242052 | RP11-190C22.1 | -0.21 | -2.24 | 2.58E-07 | 1.13E-05 |
| ENSG00000124713 | GNMT | -0.21 | -2.23 | 4.56E-07 | 1.87E-05 |
| ENSG00000184661 | CDCA2 | -0.21 | -2.26 | 7.28E-07 | 2.84E-05 |
| ENSG00000264925 | Z98949.1 | -0.21 | -2.28 | 2.96E-05 | 0.000645132 |
| ENSG00000250603 | CTC-228N24.2 | -0.21 | -2.23 | 0.00060063 | 0.007138511 |
| ENSG00000267230 | RP11-376M2.2 | -0.21 | -2.25 | 0.000677618 | 0.007825368 |
| ENSG00000251891 | RNU7-79P | -0.21 | -2.28 | 0.000692395 | 0.007975098 |
| ENSG00000260401 | RP11-800A3.4 | -0.22 | -2.18 | 2.68E-13 | 4.42E-11 |
| ENSG00000175591 | P2RY2 | -0.22 | -2.15 | 4.44E-13 | 6.91E-11 |
| ENSG00000137571 | SLCO5A1 | -0.22 | -2.21 | 9.85E-11 | 9.86E-09 |
| ENSG00000133874 | RNF122 | -0.22 | -2.17 | 5.30E-09 | 3.72E-07 |
| ENSG00000164761 | TNFRSF11B | -0.22 | -2.19 | 1.38E-07 | 6.70E-06 |
| ENSG00000232867 | RP11-179D22.1 | -0.22 | -2.20 | 9.15E-05 | 0.001635707 |
| ENSG00000136928 | GABBR2 | -0.22 | -2.20 | 0.000164509 | 0.002622183 |
| ENSG00000222371 | RN7SKP202 | -0.22 | -2.16 | 0.000172618 | 0.002719088 |
| ENSG00000153234 | NR4A2 | -0.22 | -2.21 | 0.000416971 | 0.005434558 |
| ENSG00000166589 | CDH16 | -0.22 | -2.17 | 0.000446758 | 0.005741968 |
| ENSG00000256443 | RP11-794G24.1 | -0.22 | -2.19 | 0.0007825 | 0.008694418 |
| ENSG00000175592 | FOSL1 | -0.22 | -2.20 | 0.000928391 | 0.009898012 |
| ENSG00000229871 | RP4-710M16.1 | -0.22 | -2.17 | 0.001361605 | 0.013242554 |
| ENSG00000204936 | CD177 | -0.22 | -2.19 | 0.003701024 | 0.028163066 |
| ENSG00000124882 | EREG | -0.22 | -2.18 | 0.004267964 | 0.031264917 |
| ENSG00000168824 | NSG1 | -0.23 | -2.14 | 1.16E-08 | 7.32E-07 |
| ENSG00000261616 | RP11-6O2.3 | -0.23 | -2.13 | 1.29E-08 | 8.04E-07 |
| ENSG00000222389 | RNU2-28P | -0.23 | -2.14 | 0.000106282 | 0.001842798 |
| ENSG00000152292 | SH2D6 | -0.23 | -2.09 | 0.001680784 | 0.015495701 |
| ENSG00000145832 | SLC25A48 | -0.23 | -2.11 | 0.002905126 | 0.02359247 |
| ENSG00000265055 | AC145343.2 | -0.23 | -2.15 | 0.004012638 | 0.029939994 |
| ENSG00000272799 | RP11-474N24.6 | -0.23 | -2.10 | 0.004316407 | 0.031522356 |
| ENSG00000241211 | IQCJ-SCHIP1-AS1 | -0.24 | -2.09 | 2.56E-06 | 8.44E-05 |
| ENSG00000236423 | LINC01134 | -0.24 | -2.08 | 0.000173458 | 0.002728134 |
| ENSG00000224743 | TEX26-AS1 | -0.24 | -2.08 | 0.001190362 | 0.011974457 |
| ENSG00000139998 | RAB15 | -0.25 | -1.99 | 6.24E-11 | 6.43E-09 |
| ENSG00000099625 | C19orf26 | -0.25 | -1.98 | 1.14E-09 | 9.32E-08 |
| ENSG00000080031 | PTPRH | -0.25 | -1.99 | 2.72E-07 | 1.19E-05 |
| ENSG00000255308 | RP11-428C19.4 | -0.25 | -1.98 | 6.43E-06 | 0.00018425 |
| ENSG00000255414 | LINC01059 | -0.25 | -2.01 | 5.83E-05 | 0.001135759 |
| ENSG00000237133 | AC020594.5 | -0.25 | -1.98 | 5.93E-05 | 0.001150577 |
| ENSG00000114529 | C3orf52 | -0.25 | -1.99 | 0.000110642 | 0.001904465 |
| ENSG00000158859 | ADAMTS4 | -0.25 | -1.98 | 0.000292089 | 0.004118783 |
| ENSG00000214814 | FER1L6 | -0.25 | -1.99 | 0.001832384 | 0.016659069 |
| ENSG00000125740 | FOSB | -0.25 | -1.98 | 0.003845415 | 0.02898974 |
| ENSG00000173918 | C1QTNF1 | -0.26 | -1.94 | 6.60E-09 | 4.45E-07 |
| ENSG00000134107 | BHLHE40 | -0.26 | -1.93 | 1.48E-07 | 7.05E-06 |
| ENSG00000273237 | CTB-119C2.1 | -0.26 | -1.95 | 1.76E-07 | 8.14E-06 |
| ENSG00000258987 | RP11-131H24.4 | -0.26 | -1.93 | 1.06E-05 | 0.000281557 |
| ENSG00000204928 | GRXCR2 | -0.26 | -1.96 | 5.75E-05 | 0.001123581 |
| ENSG00000240404 | RP11-142L1.3 | -0.26 | -1.96 | 0.000251372 | 0.003675091 |
| ENSG00000259727 | RP11-1069G10.2 | -0.26 | -1.93 | 0.000723952 | 0.0081976 |
| ENSG00000140297 | GCNT3 | -0.26 | -1.94 | 0.000733201 | 0.00828103 |
| ENSG00000272275 | RP11-791G15.2 | -0.26 | -1.92 | 0.000985532 | 0.010345907 |
| ENSG00000103449 | SALL1 | -0.26 | -1.94 | 0.004975142 | 0.034929559 |
| ENSG00000239474 | KLHL41 | -0.27 | -1.89 | 8.76E-10 | 7.34E-08 |
| ENSG00000251628 | RP11-371M22.1 | -0.27 | -1.87 | 2.28E-07 | 1.02E-05 |
| ENSG00000142748 | FCN3 | -0.27 | -1.87 | 2.38E-07 | 1.06E-05 |
| ENSG00000102554 | KLF5 | -0.27 | -1.90 | 3.73E-07 | 1.58E-05 |
| ENSG00000091879 | ANGPT2 | -0.27 | -1.91 | 1.85E-06 | 6.36E-05 |
| ENSG00000256540 | RP11-598F7.6 | -0.27 | -1.87 | 3.32E-05 | 0.000709154 |
| ENSG00000163638 | ADAMTS9 | -0.27 | -1.86 | 0.000281006 | 0.004004662 |
| ENSG00000109321 | AREG | -0.27 | -1.87 | 0.003270984 | 0.025834989 |
| ENSG00000226337 | RP11-274B18.4 | -0.27 | -1.90 | 0.004473373 | 0.032331476 |
| ENSG00000143125 | PROK1 | -0.27 | -1.90 | 0.004487249 | 0.032401369 |
| ENSG00000254491 | RP11-145O15.2 | -0.27 | -1.87 | 0.008122171 | 0.049946814 |
| ENSG00000114737 | CISH | -0.28 | -1.82 | 5.60E-08 | 2.98E-06 |
| ENSG00000016391 | CHDH | -0.28 | -1.86 | 1.22E-07 | 6.00E-06 |
| ENSG00000272782 | RP4-607J23.2 | -0.28 | -1.85 | 1.49E-07 | 7.06E-06 |
| ENSG00000137331 | IER3 | -0.28 | -1.84 | 4.78E-06 | 0.000142273 |
| ENSG00000253925 | CTB-178M22.1 | -0.28 | -1.82 | 1.72E-05 | 0.000414472 |
| ENSG00000177452 | RP4-597J3.1 | -0.28 | -1.81 | 9.15E-05 | 0.001635707 |
| ENSG00000168143 | FAM83B | -0.28 | -1.82 | 0.000709842 | 0.008106348 |
| ENSG00000128342 | LIF | -0.28 | -1.83 | 0.00122799 | 0.012268541 |
| ENSG00000265768 | MIR4506 | -0.28 | -1.84 | 0.002258621 | 0.019509496 |
| ENSG00000124249 | KCNK15 | -0.28 | -1.82 | 0.002761685 | 0.022702849 |
| ENSG00000157856 | DRC1 | -0.28 | -1.85 | 0.003161889 | 0.025171794 |
| ENSG00000227158 | AC073621.2 | -0.28 | -1.86 | 0.004492554 | 0.032424476 |
| ENSG00000163735 | CXCL5 | -0.28 | -1.84 | 0.006176457 | 0.04090774 |
| ENSG00000180999 | C1orf105 | -0.29 | -1.77 | 5.62E-09 | 3.88E-07 |
| ENSG00000237512 | UNC5B-AS1 | -0.29 | -1.79 | 8.51E-08 | 4.41E-06 |
| ENSG00000137193 | PIM1 | -0.29 | -1.77 | 1.62E-07 | 7.63E-06 |
| ENSG00000122861 | PLAU | -0.29 | -1.80 | 4.20E-06 | 0.000127838 |
| ENSG00000069399 | BCL3 | -0.29 | -1.81 | 7.54E-06 | 0.000210834 |
| ENSG00000229644 | NAMPTL | -0.29 | -1.80 | 1.05E-05 | 0.000278939 |
| ENSG00000185022 | MAFF | -0.29 | -1.80 | 2.45E-05 | 0.000549232 |
| ENSG00000228056 | CFL1P3 | -0.29 | -1.77 | 8.60E-05 | 0.00155669 |
| ENSG00000167971 | CASKIN1 | -0.29 | -1.81 | 0.000102088 | 0.001785431 |
| ENSG00000253978 | CTB-178M22.2 | -0.29 | -1.79 | 0.000143735 | 0.002359411 |
| ENSG00000260018 | RP11-505K9.1 | -0.29 | -1.78 | 0.000152863 | 0.002477575 |
| ENSG00000260953 | RP11-426C22.6 | -0.29 | -1.77 | 0.000403411 | 0.005312897 |
| ENSG00000188015 | S100A3 | -0.29 | -1.80 | 0.000551669 | 0.006705261 |
| ENSG00000196361 | ELAVL3 | -0.29 | -1.79 | 0.000583644 | 0.0069917 |
| ENSG00000250137 | RP11-380P13.1 | -0.29 | -1.77 | 0.000719259 | 0.008168477 |
| ENSG00000241319 | SETP6 | -0.29 | -1.79 | 0.000904968 | 0.009728883 |
| ENSG00000243225 | RP11-7F17.1 | -0.29 | -1.76 | 0.001596824 | 0.014934983 |
| ENSG00000178125 | PPP1R42 | -0.29 | -1.77 | 0.001782396 | 0.016262188 |
| ENSG00000226807 | MROH5 | -0.29 | -1.79 | 0.002155544 | 0.018883844 |
| ENSG00000179242 | CDH4 | -0.29 | -1.81 | 0.002390761 | 0.020354168 |
| ENSG00000255650 | FAM222A-AS1 | -0.29 | -1.76 | 0.004253644 | 0.031197104 |
| ENSG00000199157 | MIR208A | -0.29 | -1.81 | 0.005583494 | 0.038051989 |
| ENSG00000104369 | JPH1 | -0.3 | -1.75 | 1.02E-09 | 8.52E-08 |
| ENSG00000250588 | IQCJ-SCHIP1 | -0.3 | -1.71 | 1.72E-07 | 7.99E-06 |
| ENSG00000177508 | IRX3 | -0.3 | -1.74 | 3.29E-06 | 0.000104656 |
| ENSG00000156500 | FAM122C | -0.3 | -1.74 | 2.38E-05 | 0.000536997 |
| ENSG00000259630 | CTD-2262B20.1 | -0.3 | -1.72 | 0.00011684 | 0.001997752 |
| ENSG00000170835 | CEL | -0.3 | -1.75 | 0.000275575 | 0.003947146 |
| ENSG00000186510 | CLCNKA | -0.3 | -1.76 | 0.000494778 | 0.006201025 |
| ENSG00000261787 | TCF24 | -0.3 | -1.72 | 0.000717184 | 0.008153927 |
| ENSG00000166407 | LMO1 | -0.3 | -1.75 | 0.00291474 | 0.023658079 |
| ENSG00000135248 | FAM71F1 | -0.3 | -1.73 | 0.006218971 | 0.041100966 |
| ENSG00000188176 | SMTNL2 | -0.31 | -1.69 | 4.40E-06 | 0.000133163 |
| ENSG00000071282 | LMCD1 | -0.31 | -1.70 | 8.03E-06 | 0.000222221 |
| ENSG00000249816 | LINC00964 | -0.31 | -1.70 | 3.21E-05 | 0.000687247 |
| ENSG00000130844 | ZNF331 | -0.31 | -1.67 | 4.24E-05 | 0.00086856 |
| ENSG00000239332 | LINC01119 | -0.31 | -1.70 | 0.00025272 | 0.003690256 |
| ENSG00000151164 | RAD9B | -0.31 | -1.68 | 0.002055351 | 0.018214688 |
| ENSG00000258676 | RP11-386M24.3 | -0.31 | -1.70 | 0.003344067 | 0.026230662 |
| ENSG00000109758 | HGFAC | -0.31 | -1.68 | 0.005671288 | 0.038412412 |
| ENSG00000124818 | OPN5 | -0.31 | -1.69 | 0.007267652 | 0.046046394 |
| ENSG00000129173 | E2F8 | -0.32 | -1.62 | 1.08E-08 | 6.87E-07 |
| ENSG00000105825 | TFPI2 | -0.32 | -1.65 | 1.46E-05 | 0.000361046 |
| ENSG00000261488 | RP11-757F18.5 | -0.32 | -1.63 | 0.000207151 | 0.003128688 |
| ENSG00000178752 | FAM132B | -0.32 | -1.63 | 0.000420496 | 0.00546933 |
| ENSG00000179388 | EGR3 | -0.32 | -1.63 | 0.002571131 | 0.021503563 |
| ENSG00000229854 | RP11-524G24.2 | -0.32 | -1.65 | 0.003527717 | 0.027275776 |
| ENSG00000251603 | RP11-164P12.4 | -0.33 | -1.61 | 1.15E-09 | 9.38E-08 |
| ENSG00000177464 | GPR4 | -0.33 | -1.62 | 4.99E-07 | 2.03E-05 |
| ENSG00000163132 | MSX1 | -0.33 | -1.60 | 2.61E-06 | 8.59E-05 |
| ENSG00000105835 | NAMPT | -0.33 | -1.61 | 3.16E-06 | 0.000101224 |
| ENSG00000130164 | LDLR | -0.33 | -1.60 | 3.58E-05 | 0.00075519 |
| ENSG00000138696 | BMPR1B | -0.33 | -1.59 | 0.000176809 | 0.002773756 |
| ENSG00000223949 | RP11-24J23.2 | -0.33 | -1.60 | 0.000507006 | 0.006321876 |
| ENSG00000232464 | CTA-125H2.1 | -0.33 | -1.60 | 0.000779814 | 0.008672577 |
| ENSG00000250994 | AC005355.1 | -0.33 | -1.62 | 0.001074398 | 0.011075037 |
| ENSG00000263293 | RP11-290H9.4 | -0.33 | -1.58 | 0.002128155 | 0.018702394 |
| ENSG00000241158 | ADAMTS9-AS1 | -0.33 | -1.58 | 0.002162492 | 0.018923189 |
| ENSG00000106236 | NPTX2 | -0.33 | -1.60 | 0.002451214 | 0.020739699 |
| ENSG00000163661 | PTX3 | -0.33 | -1.61 | 0.004245308 | 0.031165637 |
| ENSG00000162494 | LRRC38 | -0.33 | -1.60 | 0.005119661 | 0.035623048 |
| ENSG00000166831 | RBPMS2 | -0.34 | -1.56 | 8.45E-11 | 8.59E-09 |
| ENSG00000095794 | CREM | -0.34 | -1.57 | 4.11E-08 | 2.28E-06 |
| ENSG00000159388 | BTG2 | -0.34 | -1.54 | 1.30E-05 | 0.000327119 |
| ENSG00000099860 | GADD45B | -0.34 | -1.55 | 2.65E-05 | 0.000586593 |
| ENSG00000162174 | ASRGL1 | -0.34 | -1.56 | 0.000129204 | 0.002167076 |
| ENSG00000239482 | RP11-90K6.1 | -0.34 | -1.56 | 0.000250195 | 0.003662125 |
| ENSG00000251450 | CTC-459I6.1 | -0.34 | -1.56 | 0.005128443 | 0.035636926 |
| ENSG00000236039 | AC019117.2 | -0.34 | -1.55 | 0.005294016 | 0.036507148 |
| ENSG00000113889 | KNG1 | -0.34 | -1.55 | 0.005768856 | 0.038841217 |
| ENSG00000065320 | NTN1 | -0.35 | -1.51 | 1.06E-09 | 8.82E-08 |
| ENSG00000184545 | DUSP8 | -0.35 | -1.51 | 5.91E-09 | 4.04E-07 |
| ENSG00000137094 | DNAJB5 | -0.35 | -1.52 | 8.22E-09 | 5.41E-07 |
| ENSG00000167470 | MIDN | -0.35 | -1.52 | 9.70E-07 | 3.64E-05 |
| ENSG00000244921 | CTB-36O1.7 | -0.35 | -1.53 | 1.55E-06 | 5.39E-05 |
| ENSG00000131459 | GFPT2 | -0.35 | -1.53 | 7.38E-05 | 0.001368504 |
| ENSG00000248986 | RP11-774O3.1 | -0.35 | -1.51 | 0.000115381 | 0.001974995 |
| ENSG00000244020 | MT1HL1 | -0.35 | -1.50 | 0.000287353 | 0.004070637 |
| ENSG00000227630 | LINC01132 | -0.35 | -1.50 | 0.000526397 | 0.006508351 |
| ENSG00000260806 | RP11-872J21.3 | -0.35 | -1.51 | 0.001067326 | 0.011013192 |
| ENSG00000227227 | AC017101.10 | -0.35 | -1.51 | 0.001632426 | 0.015183423 |
| ENSG00000253519 | AC106801.1 | -0.35 | -1.52 | 0.002862387 | 0.023350015 |
