## Supplementary material for "Specific methylation marks in promoter regions are associated to the pathogenic process of Chronic Chagas disease Cardiomyopathy by modifying transcription factor binding patterns": Supplementary table 7.docx

**Supplementary table 7.** Features of the tested CpG sites and of the DMPs.

| **Feature** | **Number of**  **CpG** | **Percentage of CpG** | **Number of DMPs** | **Percentage of DMPs** |
| --- | --- | --- | --- | --- |
| **Body** | 270370 | 37.46 | 7023 | 41.60 |
| **IGR** | 198834 | 27.55 | 5281 | 31.28 |
| **TSS1500** | 90600 | 12.55 | 1364 | 8.08 |
| **5'UTR** | 62505 | 8.66 | 1687 | 9.99 |
| **TSS200** | 54591 | 7.56 | 738 | 4.37 |
| **1^st^ Exon** | 21837 | 3.03 | 420 | 2.49 |
| **3'UTR** | 18063 | 2.50 | 276 | 1.63 |
| **Exon Boundary** | 5002 | 0.69 | 94 | 0.56 |
