## Supplementary material for "Specific methylation marks in promoter regions are associated to the pathogenic process of Chronic Chagas disease Cardiomyopathy by modifying transcription factor binding patterns": Supplementary table 9.docx

**Supplementary table 9.** List of the regulatory DMRs associated to the 89 DEGs.

| **DMR**  **id** | **chromosome** | **start** | **end** | **Region covered by this DMR** | **Gene** |
| --- | --- | --- | --- | --- | --- |
| DMR82 | chr1 | 9714843 | 9714845 | TSS | C1orf200 |
| DMR2 | chr1 | 25291385 | 25292215 | 1stExon,TSS | RUNX3 |
| DMR41 | chr1 | 27668098 | 27668433 | TSS | SYTL1 |
| DMR51 | chr1 | 27952665 | 27953220 | TSS,1stExon | FGR |
| DMR52 | chr1 | 28521427 | 28521540 | TSS | PTAFR |
| DMR57 | chr1 | 40783063 | 40783234 | TSS | COL9A2 |
| DMR92 | chr1 | 111415781 | 111416181 | 1stExon,5'UTR | CD53 |
| DMR17 | chr1 | 111743038 | 111743411 | 1stExon,TSS | DENND2D |
| DMR20 | chr1 | 114414312 | 114414532 | 5'UTR,TSS | PTPN22 |
| DMR72 | chr1 | 151129062 | 151129298 | TSS,5'UTR | TNFAIP8L2 |
| DMR3 | chr1 | 159046391 | 159047177 | TSS,5'UTR | AIM2 |
| DMR74 | chr1 | 159770253 | 159770368 | 5'UTR,TSS | FCRL6 |
| DMR50 | chr1 | 160714299 | 160714382 | 5'UTR | SLAMF7 |
| DMR16 | chr1 | 161039601 | 161040044 | TSS,1stExon | ARHGAP30 |
| DMR55 | chr1 | 202128508 | 202128682 | 1stExon,5'UTR | PTPN7 |
| DMR28 | chr1 | 203733914 | 203733971 | TSS | LAX1 |
| DMR23 | chr1 | 209929437 | 209929622 | 1stExon,TSS | TRAF3IP3 |
| DMR56 | chr1 | 209931334 | 209931457 | 5'UTR | TRAF3IP3 |
| DMR9 | chr1 | 209941646 | 209941848 | TSS,1stExon | TRAF3IP3 |
| DMR8 | chr10 | 49892943 | 49893549 | TSS,5'UTR | WDFY4 |
| DMR7 | chr10 | 72362694 | 72362866 | TSS | PRF1 |
| DMR34 | chr10 | 125651034 | 125651370 | 1stExon | CPXM2 |
| DMR33 | chr11 | 58981043 | 58981095 | TSS | MPEG1 |
| DMR5 | chr11 | 60738971 | 60739183 | TSS,5'UTR | CD6 |
| DMR83 | chr11 | 60869910 | 60869960 | 5'UTR,TSS | CD5 |
| DMR29 | chr11 | 64107158 | 64107517 | TSS | CCDC88B |
| DMR81 | chr11 | 67171476 | 67171585 | 5'UTR | TBC1D10C |
| DMR63 | chr11 | 67205642 | 67205650 | TSS | PTPRCAP |
| DMR38 | chr11 | 118095405 | 118095739 | 5'UTR | AMICA1 |
| DMR54 | chr11 | 118213272 | 118213330 | 1stExon,5'UTR | CD3D |
| DMR30 | chr12 | 6881595 | 6881997 | 1stExon,TSS | LAG3 |
| DMR61 | chr12 | 7060263 | 7060386 | TSS | PTPN6 |
| DMR49 | chr12 | 12224246 | 12224360 | 5'UTR | BCL2L14 |
| DMR43 | chr12 | 47610257 | 47610418 | TSS | PCED1B-AS1 |
| DMR71 | chr12 | 51718155 | 51718251 | TSS | BIN2 |
| DMR15 | chr12 | 53496729 | 53497147 | TSS | SOAT2 |
| DMR22 | chr12 | 54891491 | 54891655 | 1stExon,TSS | NCKAP1L |
| DMR26 | chr12 | 68553577 | 68553980 | TSS | IFNG |
| DMR32 | chr12 | 109026944 | 109027086 | TSS | SELPLG |
| DMR87 | chr12 | 109027683 | 109027932 | TSS | SELPLG |
| DMR76 | chr12 | 109028385 | 109028610 | TSS | SELPLG |
| DMR14 | chr14 | 75988251 | 75988820 | TSS,5'UTR | BATF |
| DMR89 | chr15 | 38988533 | 38988755 | TSS | C15orf53 |
| DMR62 | chr15 | 44969244 | 44969481 | TSS | PATL2 |
| DMR10 | chr15 | 77286232 | 77287243 | TSS | PSTPIP1 |
| DMR69 | chr15 | 81590933 | 81591058 | 5'UTR | IL16 |
| DMR18 | chr16 | 27414210 | 27414536 | 1stExon,TSS,5'UTR | IL21R |
| DMR80 | chr16 | 29673933 | 29674184 | TSS | SPN |
| DMR21 | chr16 | 29757318 | 29757565 | 1stExon,TSS | C16orf54 |
| DMR11 | chr16 | 50715260 | 50715700 | TSS,1stExon | SNX20 |
| DMR94 | chr17 | 4487099 | 4487125 | TSS | SMTNL2 |
| DMR37 | chr17 | 56408688 | 56409028 | TSS | MIR142 |
| DMR40 | chr18 | 43652592 | 43652594 | TSS | PSTPIP2 |
| DMR45 | chr19 | 3179364 | 3179741 | 1stExon | S1PR4 |
| DMR60 | chr19 | 17862017 | 17862104 | 5'UTR | FCHO1 |
| DMR46 | chr19 | 36204551 | 36204918 | 5'UTR | ZBTB32 |
| DMR48 | chr19 | 44285940 | 44285954 | TSS | KCNN4 |
| DMR64 | chr19 | 49838478 | 49838777 | TSS,1stExon | CD37 |
| DMR25 | chr2 | 10261684 | 10262019 | TSS | RRM2 |
| DMR79 | chr2 | 143886326 | 143886567 | TSS | ARHGAP15 |
| DMR27 | chr2 | 158300475 | 158300811 | 1stExon,TSS | CYTIP |
| DMR6 | chr2 | 202125088 | 202125310 | TSS,5'UTR,1stExon | CASP8 |
| DMR66 | chr2 | 204732461 | 204732474 | TSS | CTLA4 |
| DMR35 | chr2 | 225811610 | 225811669 | 1stExon | DOCK10 |
| DMR39 | chr20 | 35273933 | 35274281 | 5'UTR | SLA2 |
| DMR42 | chr20 | 56195541 | 56195574 | 1stExon | ZBP1 |
| DMR68 | chr21 | 46332181 | 46332291 | 5'UTR | ITGB2 |
| DMR77 | chr21 | 46334192 | 46334214 | 5'UTR | ITGB2 |
| DMR44 | chr22 | 50524032 | 50524541 | TSS,5'UTR | MLC1 |
| DMR84 | chr22 | 50985797 | 50986031 | TSS | KLHDC7B |
| DMR19 | chr3 | 45984838 | 45985168 | TSS,5'UTR | CXCR6 |
| DMR24 | chr3 | 46411369 | 46411474 | TSS | CCR5 |
| DMR67 | chr4 | 40201884 | 40201943 | 5'UTR | RHOH |
| DMR85 | chr4 | 100737691 | 100738011 | TSS,1stExon | DAPP1 |
| DMR65 | chr5 | 149792783 | 149792840 | TSS | CD74 |
| DMR58 | chr5 | 156607793 | 156608118 | 1stExon,TSS | ITK |
| DMR4 | chr5 | 169407439 | 169407941 | 1stExon,5'UTR,TSS | FAM196B |
| DMR88 | chr6 | 26367571 | 26367580 | 5'UTR | BTN3A2 |
| DMR53 | chr6 | 29527870 | 29527885 | TSS | UBD |
| DMR1 | chr6 | 31539539 | 31540461 | TSS,5'UTR | LTA |
| DMR70 | chr6 | 32909282 | 32909523 | TSS | HLA-DMB |
| DMR78 | chr6 | 33041343 | 33041697 | 1stExon,TSS | HLA-DPA1 |
| DMR59 | chr6 | 42391208 | 42391254 | 5'UTR | TRERF1 |
| DMR13 | chr6 | 108145374 | 108145601 | 1stExon,TSS | SCML4 |
| DMR47 | chr6 | 149805292 | 149805596 | 5'UTR | ZC3H12D |
| DMR91 | chr7 | 3067279 | 3067293 | 5'UTR | CARD11 |
| DMR86 | chr7 | 36764019 | 36764082 | 1stExon | AOAH |
| DMR73 | chr7 | 45018658 | 45019005 | 5'UTR,TSS | MYO1G |
| DMR36 | chr7 | 50358075 | 50358218 | 5'UTR | IKZF1 |
| DMR31 | chr8 | 21771446 | 21771668 | TSS | DOK2 |
| DMR75 | chr8 | 134072526 | 134072706 | 5'UTR,TSS | SLA |
| DMR90 | chr9 | 95726377 | 95726447 | 1stExon | FGD3 |
| DMR93 | chr9 | 117692745 | 117692759 | 1stExon | TNFSF8 |
| DMR12 | chr9 | 123688715 | 123689193 | 5'UTR,TSS | TRAF1 |
