## Supplementary material for "Specific methylation marks in promoter regions are associated to the pathogenic process of Chronic Chagas disease Cardiomyopathy by modifying transcription factor binding patterns": Supplementary table 16.docx

**Supplementary table 16.** Features of the DMPs on blood samples.

| **Feature** | **Number of**  **CpG** | **Percentage of**  **CpG** | **Number of**  **DMPs**  **(asymptomatic vs severe CCC)** | **Number of**  **DMPs**  **(asymptomatic vs severe CCC)** | **Number of**  **DMPs**  **(moderate CCC vs severe CCC)** | **Percentage of DMPs**  **(moderate CCC vs severe CCC)** |
| --- | --- | --- | --- | --- | --- | --- |
| **Body** | 275214 | 37.36 | 4833 | 38.28 | 2599 | 38.59 |
| **IGR** | 201739 | 27.39 | 3421 | 27.1 | 1851 | 27.48 |
| **TSS1500** | 92486 | 12.55 | 1472 | 11.66 | 837 | 12.43 |
| **5'UTR** | 63918 | 8.68 | 1119 | 8.86 | 537 | 7.97 |
| **TSS200** | 57108 | 7.75 | 929 | 7.36 | 506 | 7.51 |
| **1stExon** | 22688 | 3.08 | 427 | 3.38 | 197 | 2.93 |
| **3'UTR** | 18381 | 2.5 | 316 | 2.5 | 150 | 2.23 |
| **Exon Borders** | 5127 | 0.7 | 107 | 0.85 | 58 | 0.86 |
