## Supplementary material for "Specific methylation marks in promoter regions are associated to the pathogenic process of Chronic Chagas disease Cardiomyopathy by modifying transcription factor binding patterns": Supplementary table 19.docx

**Supplementary table 19.** List of CpG site biomarkers useful for diagnosis and prognosis.

| **Biomarkers indicating the presence of the disease (asymptomatic vs CCC)**  **198 CpG site set**  **Sensitivity: 1 specificity: 0.87** | | | | | | |
| --- | --- | --- | --- | --- | --- | --- |
| **ID** | **Chr** | **Pos** | **Gene** | **Location** | **Corrected pvalue** | **delta beta** |
| cg20703997 | 1 | 4087676 |  | IGR | 9.9E-03 | -0.105 |
| cg17667591 | 1 | 17719944 | PADI6 | Body | 5.1E-03 | -0.117 |
| cg14378564 | 1 | 28417750 |  | IGR | 3.5E-02 | -0.082 |
| cg26113488 | 1 | 39249433 |  | IGR | 4.3E-02 | -0.074 |
| cg14667685 | 1 | 39249604 |  | IGR | 4.7E-02 | -0.074 |
| cg22128527 | 1 | 39510999 |  | IGR | 1.8E-02 | -0.077 |
| cg20245116 | 1 | 55522013 | PCSK9 | Body | 5.9E-03 | -0.119 |
| cg13462158 | 1 | 55522104 | PCSK9 | Body | 1.4E-02 | -0.107 |
| cg15821589 | 1 | 78444904 | FUBP1 | TSS200 | 1.8E-02 | 0.083 |
| cg23656322 | 1 | 153533922 | S100A2 | Body | 2.7E-02 | 0.072 |
| cg11569930 | 1 | 155113381 | DPM3 | TSS1500 | 3.4E-05 | -0.126 |
| cg03441844 | 1 | 161368947 |  | IGR | 6.5E-03 | 0.089 |
| cg00864916 | 1 | 180886483 | KIAA1614 | Body | 4.1E-02 | -0.079 |
| cg02586212 | 1 | 192544902 | RGS1 | 1stExon | 7.8E-03 | -0.105 |
| cg17360849 | 1 | 201688419 | MIR5191 | TSS1500 | 2.5E-02 | 0.082 |
| cg13615839 | 1 | 218094953 |  | IGR | 2.2E-04 | 0.076 |
| cg12880874 | 2 | 2942535 | LINC01250 | Body | 1.7E-02 | 0.140 |
| cg12135269 | 2 | 3642586 | COLEC11 | 1stExon | 1.6E-02 | -0.072 |
| cg19867917 | 2 | 3642629 | COLEC11 | TSS200 | 1.3E-02 | -0.080 |
| cg00835279 | 2 | 3642710 | COLEC11 | 1stExon | 9.4E-03 | -0.082 |
| cg17872886 | 2 | 3642732 | COLEC11 | 1stExon | 5.0E-03 | -0.073 |
| cg17872867 | 2 | 3885178 | DCDC2C | Body | 3.3E-03 | 0.074 |
| cg22708768 | 2 | 7813295 |  | IGR | 2.9E-02 | 0.088 |
| cg16842187 | 2 | 26951473 | KCNK3 | 3'UTR | 3.5E-03 | -0.086 |
| cg02148796 | 2 | 29548577 | ALK | Body | 5.7E-03 | 0.112 |
| cg11765362 | 2 | 31806551 | SRD5A2 | TSS1500 | 1.7E-03 | -0.091 |
| cg00712943 | 2 | 31806781 | SRD5A2 | TSS1500 | 5.7E-03 | -0.082 |
| cg13968390 | 2 | 108904812 | SULT1C2 | TSS1500 | 2.1E-02 | -0.113 |
| cg25838818 | 2 | 108905173 | SULT1C2 | 5'UTR | 2.6E-02 | -0.087 |
| cg11942181 | 2 | 129659946 |  | IGR | 1.8E-03 | -0.075 |
| cg20416831 | 2 | 137521943 |  | IGR | 3.0E-03 | -0.073 |
| cg08444004 | 2 | 240086178 | HDAC4 | Body | 4.1E-02 | 0.110 |
| cg00935887 | 2 | 242844000 |  | IGR | 1.9E-02 | -0.097 |
| cg22334681 | 3 | 14257893 |  | IGR | 3.9E-02 | 0.124 |
| cg27084712 | 3 | 42977845 |  | IGR | 9.1E-03 | -0.074 |
| cg21104412 | 3 | 42978026 |  | IGR | 1.1E-02 | -0.084 |
| cg23275390 | 3 | 105601367 |  | IGR | 4.8E-02 | 0.102 |
| cg16391801 | 3 | 135794793 | PPP2R3A | Body | 1.0E-02 | 0.096 |
| cg10599156 | 3 | 141145231 | ZBTB38 | 5'UTR | 1.3E-02 | 0.115 |
| cg10725542 | 3 | 149094653 | TM4SF1 | Body | 2.7E-02 | 0.098 |
| cg26306106 | 3 | 149094816 | TM4SF1 | Body | 2.9E-02 | 0.096 |
| cg23379566 | 3 | 149094892 | TM4SF1 | Body | 3.4E-02 | 0.105 |
| cg26584465 | 3 | 149095006 | TM4SF1 | Body | 2.5E-02 | 0.080 |
| cg08124030 | 3 | 149095283 | TM4SF1 | 1stExon | 5.6E-03 | 0.130 |
| cg02909206 | 3 | 194574434 |  | IGR | 4.4E-04 | 0.099 |
| cg02294690 | 4 | 17581984 | LAP3 | Body | 1.5E-02 | -0.098 |
| cg06113917 | 4 | 66809014 |  | IGR | 7.7E-03 | 0.082 |
| cg14493094 | 4 | 132896579 |  | IGR | 3.3E-02 | -0.083 |
| cg06070696 | 4 | 166662284 | LINC01179 | Body | 1.2E-02 | -0.073 |
| cg21259215 | 4 | 174834360 |  | IGR | 1.3E-02 | -0.079 |
| cg21873524 | 4 | 190942744 |  | IGR | 1.1E-02 | -0.146 |
| cg21795442 | 5 | 34517523 |  | IGR | 4.4E-02 | -0.083 |
| cg10606776 | 5 | 55610640 |  | IGR | 6.1E-03 | -0.092 |
| cg23500924 | 5 | 140465235 |  | IGR | 1.0E-02 | -0.082 |
| cg08626876 | 5 | 140501540 | PCDHB4 | TSS200 | 4.1E-04 | -0.075 |
| cg10935175 | 5 | 140614727 | PCDHB18 | Body | 1.0E-02 | -0.076 |
| cg01828067 | 5 | 140628308 |  | IGR | 4.1E-04 | -0.105 |
| cg16831002 | 5 | 140749358 | PCDHGB3 | TSS1500 | 1.5E-03 | -0.086 |
| cg07524997 | 5 | 140796892 | PCDHGA4 | Body | 1.5E-03 | -0.082 |
| cg10094930 | 5 | 178194658 | AACSP1 | Body | 3.8E-03 | -0.078 |
| cg23329272 | 5 | 180085924 |  | IGR | 4.0E-02 | 0.076 |
| cg06588529 | 6 | 4154091 | LOC100507506 | Body | 4.4E-02 | -0.100 |
| cg13746813 | 6 | 14911904 |  | IGR | 5.5E-03 | 0.112 |
| cg08818610 | 6 | 24910720 | FAM65B | 5'UTR | 8.4E-03 | -0.073 |
| cg25133685 | 6 | 29013336 | OR2W1 | TSS1500 | 1.9E-02 | 0.072 |
| cg08269402 | 6 | 32549631 | HLA-DRB1 | Body | 2.8E-03 | -0.206 |
| cg00650978 | 6 | 108684632 | LACE1 | Body | 8.6E-07 | 0.091 |
| cg02872426 | 6 | 110736772 | DDO | TSS200 | 4.2E-02 | 0.073 |
| cg07164639 | 6 | 110736958 | DDO | TSS1500 | 4.7E-03 | 0.074 |
| cg06696596 | 6 | 122391246 |  | IGR | 4.0E-02 | -0.079 |
| cg24246628 | 6 | 168435914 | KIF25 | Body | 3.4E-02 | -0.101 |
| cg24642844 | 7 | 1081250 | C7orf50 | Body | 2.6E-02 | 0.076 |
| cg21728101 | 7 | 1979360 | MAD1L1 | Body | 3.8E-03 | -0.086 |
| cg22772380 | 7 | 6747037 | ZNF12 | TSS1500 | 1.9E-02 | -0.117 |
| cg10871071 | 7 | 31553412 | CCDC129 | TSS1500 | 1.7E-02 | -0.079 |
| cg18850127 | 7 | 39170497 | POU6F2 | Body | 7.0E-03 | -0.117 |
| cg15212455 | 7 | 39170539 | POU6F2 | Body | 5.0E-03 | -0.137 |
| cg20302533 | 7 | 39170763 | POU6F2 | Body | 1.2E-02 | -0.081 |
| cg08835956 | 7 | 39171034 | POU6F2 | Body | 1.6E-03 | -0.101 |
| cg10621924 | 7 | 39171070 | POU6F2 | Body | 4.4E-03 | -0.080 |
| cg21665744 | 7 | 39171113 | POU6F2 | Body | 3.5E-03 | -0.104 |
| cg00050402 | 7 | 55073022 |  | IGR | 5.3E-03 | -0.094 |
| cg13881108 | 7 | 55073432 |  | IGR | 9.8E-03 | -0.091 |
| cg13335691 | 7 | 56242142 |  | IGR | 3.3E-03 | -0.078 |
| cg11460282 | 7 | 56297661 |  | IGR | 3.4E-02 | -0.091 |
| cg00538212 | 7 | 158751591 |  | IGR | 1.8E-02 | -0.083 |
| cg18002480 | 8 | 672007 | ERICH1 | Body | 3.6E-02 | -0.072 |
| cg17179911 | 8 | 674434 | ERICH1 | Body | 2.2E-02 | -0.097 |
| cg21650737 | 8 | 674525 | ERICH1 | Body | 3.6E-02 | -0.105 |
| cg27048067 | 8 | 674560 | ERICH1 | Body | 1.2E-02 | -0.114 |
| cg25569760 | 8 | 6518284 | MCPH1-AS1 | Body | 7.2E-03 | 0.075 |
| cg02269453 | 8 | 9009499 | PPP1R3B | TSS1500 | 6.5E-03 | -0.083 |
| cg04520078 | 8 | 43101622 |  | IGR | 9.9E-03 | -0.079 |
| cg18607583 | 8 | 63387061 | NKAIN3 | Body | 5.0E-04 | 0.084 |
| cg06928823 | 8 | 73533533 | KCNB2 | Body | 5.6E-03 | -0.088 |
| cg08270491 | 8 | 126100743 | KIAA0196 | 5'UTR | 2.8E-02 | 0.101 |
| cg07830218 | 9 | 6714365 |  | IGR | 1.7E-02 | 0.073 |
| cg02300840 | 9 | 23829435 |  | IGR | 1.7E-02 | -0.074 |
| cg19097407 | 9 | 36154750 | GLIPR2 | Body | 5.0E-02 | 0.120 |
| cg09466719 | 9 | 36277154 | GNE | TSS200 | 4.9E-02 | 0.072 |
| cg03363289 | 9 | 124990165 | LHX6 | Body | 3.7E-02 | -0.144 |
| cg16733545 | 9 | 133026410 |  | IGR | 2.0E-04 | -0.077 |
| cg08046642 | 10 | 3939393 |  | IGR | 1.8E-03 | 0.075 |
| cg24591913 | 10 | 47062881 |  | IGR | 1.4E-02 | 0.127 |
| cg05119883 | 10 | 80752407 | ZMIZ1-AS1 | Body | 4.3E-02 | 0.088 |
| cg17828988 | 10 | 81951504 | ANXA11 | 5'UTR | 5.1E-03 | 0.071 |
| cg12793466 | 10 | 109673222 | LINC01435 | Body | 7.2E-03 | 0.079 |
| cg24536568 | 10 | 134746366 | CFAP46 | Body | 4.6E-02 | -0.078 |
| cg13507983 | 11 | 5128882 |  | IGR | 1.8E-03 | -0.074 |
| cg04585209 | 11 | 6292311 | CCKBR | Body | 3.2E-02 | -0.099 |
| cg18706028 | 11 | 6292490 | CCKBR | Body | 4.6E-02 | -0.112 |
| cg26700447 | 11 | 6292511 | CCKBR | Body | 4.2E-02 | -0.100 |
| cg08449403 | 11 | 8632315 |  | IGR | 3.1E-02 | 0.125 |
| cg01058253 | 11 | 9748095 | SWAP70 | Body | 4.8E-04 | 0.076 |
| cg05226335 | 11 | 70253499 | CTTN | Body | 1.6E-03 | 0.102 |
| cg07136909 | 11 | 71278894 |  | IGR | 4.4E-02 | -0.102 |
| cg06978117 | 11 | 111155014 | C11orf53 | Body | 9.2E-03 | -0.074 |
| cg03554573 | 11 | 111155021 | C11orf53 | Body | 6.3E-03 | -0.078 |
| cg11805729 | 11 | 114633677 |  | IGR | 6.6E-03 | -0.072 |
| cg07109603 | 12 | 24049014 | SOX5 | Body | 1.4E-02 | -0.086 |
| cg13281312 | 12 | 77719842 |  | IGR | 1.7E-02 | -0.098 |
| cg07120026 | 12 | 122287892 | HPD | Body | 1.8E-02 | 0.077 |
| cg24288706 | 12 | 122287927 | HPD | Body | 2.9E-02 | 0.085 |
| cg24704312 | 12 | 123310691 | CCDC62 | 3'UTR | 1.8E-02 | -0.105 |
| cg15509177 | 13 | 19919208 | LOC100101938 | TSS200 | 3.4E-02 | -0.087 |
| cg19692240 | 13 | 21748982 | MRPL57 | TSS1500 | 1.7E-03 | 0.087 |
| cg09460553 | 13 | 110521956 |  | IGR | 8.9E-03 | -0.095 |
| cg02315096 | 13 | 110522020 |  | IGR | 6.0E-03 | -0.115 |
| cg18865445 | 13 | 110522265 |  | IGR | 3.8E-03 | -0.095 |
| cg17797229 | 13 | 110522297 |  | IGR | 4.3E-03 | -0.135 |
| cg20256375 | 13 | 112730681 |  | IGR | 2.7E-02 | -0.082 |
| cg06470855 | 13 | 112997365 |  | IGR | 1.5E-02 | -0.081 |
| cg25787588 | 14 | 50784952 | ATP5S | Body | 1.6E-02 | -0.101 |
| cg02291164 | 14 | 59296302 |  | IGR | 4.2E-02 | -0.078 |
| cg06898279 | 14 | 60629192 | DHRS7 | Body | 1.9E-02 | 0.077 |
| cg24000535 | 14 | 91110600 | LOC101928909 | Body | 1.2E-02 | -0.107 |
| cg17973977 | 14 | 91687281 | C14orf159 | Body | 2.4E-04 | 0.078 |
| cg18561199 | 14 | 95027379 | SERPINA4 | TSS1500 | 1.8E-03 | -0.263 |
| cg15451504 | 14 | 106267680 |  | IGR | 1.2E-02 | -0.097 |
| cg23088318 | 15 | 25093985 | SNRPN | 5'UTR | 2.3E-03 | 0.111 |
| cg18457085 | 15 | 33602455 | LOC101928134 | Body | 2.3E-02 | -0.073 |
| cg06684395 | 15 | 35351277 |  | IGR | 1.3E-02 | 0.073 |
| cg23849826 | 15 | 39872186 | THBS1 | TSS1500 | 2.7E-02 | -0.073 |
| cg11036485 | 15 | 43924717 | CATSPER2 | Body | 5.7E-03 | -0.092 |
| cg17582782 | 15 | 57592438 | LOC283663 | TSS200 | 5.0E-02 | 0.080 |
| cg11193064 | 15 | 67008444 | SMAD6 | Body | 3.3E-02 | 0.113 |
| cg19684296 | 15 | 67008514 | SMAD6 | Body | 1.4E-02 | 0.079 |
| cg06680906 | 15 | 69325560 | MIR548H4 | Body | 1.1E-02 | -0.072 |
| cg18128914 | 15 | 74244249 | LOXL1 | 3'UTR | 1.5E-03 | 0.117 |
| cg27398640 | 15 | 77910606 | LINGO1 | Body | 7.3E-03 | 0.104 |
| cg06211550 | 16 | 3230726 |  | IGR | 2.8E-02 | 0.082 |
| cg11322254 | 16 | 12659506 | SNX29 | Body | 8.5E-03 | 0.072 |
| cg17391820 | 16 | 34256336 |  | IGR | 4.6E-02 | -0.072 |
| cg03998338 | 16 | 55794910 | CES4 | Body | 2.8E-02 | -0.079 |
| cg04773618 | 16 | 58708863 | SLC38A7 | Body | 2.4E-02 | 0.075 |
| cg08457029 | 16 | 75313404 |  | IGR | 3.2E-02 | -0.075 |
| cg16582469 | 16 | 88910141 | GALNS | Body | 1.9E-02 | 0.073 |
| cg24367698 | 17 | 10453091 | MYH2 | TSS200 | 1.9E-03 | 0.091 |
| cg03641585 | 17 | 10761400 |  | IGR | 3.2E-02 | -0.098 |
| cg23280506 | 17 | 14201938 |  | IGR | 9.7E-03 | 0.072 |
| cg25393494 | 17 | 17109936 | PLD6 | TSS1500 | 1.8E-04 | -0.121 |
| cg16340422 | 17 | 17110120 | PLD6 | TSS1500 | 1.9E-03 | -0.095 |
| cg24578857 | 17 | 17110207 | PLD6 | TSS1500 | 6.6E-03 | -0.083 |
| cg14560110 | 17 | 20799470 | CCDC144NL | TSS200 | 1.7E-02 | -0.090 |
| cg08288433 | 17 | 20799511 | CCDC144NL | TSS200 | 5.8E-03 | -0.091 |
| cg06809326 | 17 | 20799526 | CCDC144NL | TSS200 | 5.1E-03 | -0.099 |
| cg22570042 | 17 | 20799532 | CCDC144NL | TSS200 | 8.7E-03 | -0.106 |
| cg03168497 | 17 | 48586147 | MYCBPAP | Body | 7.8E-03 | 0.084 |
| cg27366766 | 17 | 56565286 | HSF5 | 1stExon | 2.3E-02 | -0.074 |
| cg22674497 | 17 | 72916000 | USH1G | Body | 5.8E-03 | -0.078 |
| cg04585822 | 17 | 72916509 | USH1G | Body | 1.2E-02 | -0.072 |
| cg27176264 | 17 | 81060149 |  | IGR | 3.8E-02 | -0.089 |
| cg20097520 | 18 | 8835912 |  | IGR | 6.5E-04 | 0.087 |
| cg21894287 | 18 | 72837531 |  | IGR | 3.9E-03 | -0.086 |
| cg18709881 | 18 | 72837627 |  | IGR | 1.9E-03 | -0.072 |
| cg14395744 | 18 | 72837693 |  | IGR | 6.3E-03 | -0.085 |
| cg04756515 | 18 | 72837700 |  | IGR | 8.2E-03 | -0.079 |
| cg05170020 | 18 | 74874153 |  | IGR | 1.7E-02 | 0.073 |
| cg03511628 | 18 | 77280586 | NFATC1 | Body | 6.0E-03 | 0.078 |
| cg12648671 | 18 | 77377538 |  | IGR | 2.1E-02 | -0.087 |
| cg20697926 | 18 | 77377589 |  | IGR | 1.6E-02 | -0.098 |
| cg12577633 | 18 | 77623199 | KCNG2 | TSS1500 | 6.0E-04 | -0.078 |
| cg23825213 | 18 | 77623475 | KCNG2 | TSS200 | 7.0E-03 | -0.087 |
| cg03682112 | 18 | 77623598 | KCNG2 | TSS200 | 1.1E-03 | -0.078 |
| cg04173586 | 19 | 2167496 | DOT1L | Body | 2.0E-02 | 0.078 |
| cg26455718 | 19 | 36076992 |  | IGR | 2.8E-03 | -0.074 |
| cg14061069 | 19 | 46274453 | DMPK | Body | 2.6E-02 | 0.080 |
| cg23899409 | 19 | 54850582 | LILRA4 | TSS200 | 4.2E-02 | 0.075 |
| cg26411441 | 20 | 3733040 | HSPA12B | 3'UTR | 1.8E-02 | 0.076 |
| cg24047921 | 20 | 43169720 | PKIG | 5'UTR | 2.0E-02 | 0.087 |
| cg25301532 | 20 | 43378953 | KCNK15 | Body | 2.3E-02 | -0.098 |
| cg16667508 | 20 | 43936853 | MATN4 | 1stExon | 4.9E-02 | -0.092 |
| cg14102530 | 20 | 52680924 | BCAS1 | 5'UTR | 1.5E-02 | -0.116 |
| cg12058843 | 20 | 57844021 |  | IGR | 3.6E-02 | -0.076 |
| cg12080266 | 21 | 36060640 | CLIC6 | Body | 4.9E-02 | 0.155 |
| cg13270055 | 22 | 36960499 | CACNG2 | Body | 6.4E-03 | -0.083 |
| cg10491108 | 22 | 37409263 | TST | Body | 1.2E-02 | -0.110 |
| cg22343728 | 22 | 46287028 |  | IGR | 7.0E-03 | -0.102 |

| **Biomarkers indicating the presence of the disease (moderate CCC vs severe CCC)**  **61 CpG site set**  **Sensitivity: 0.92 specificity: 1** | | | | | | |
| --- | --- | --- | --- | --- | --- | --- |
| **ID** | **Chr** | **Pos** | **Gene** | **Location** | **Corrected p value** | **delta beta** |
| cg25880573 | 1 | 24073129 | TCEB3 | Body | 1.5E-02 | 0.033 |
| cg13704271 | 1 | 32666264 | CCDC28B | 1stExon | 1.0E-02 | 0.009 |
| cg26687031 | 1 | 38269629 | YRDC | Body | 4.4E-02 | 0.006 |
| cg24591770 | 1 | 45082704 | RNF220 | Body | 1.6E-02 | 0.013 |
| cg18015640 | 1 | 93310900 | FAM69A | Body | 1.5E-02 | 0.012 |
| cg15998505 | 1 | 153479701 |  | IGR | 4.4E-02 | 0.018 |
| cg25905016 | 2 | 24267482 | C2orf44 | 5'UTR | 4.4E-02 | 0.020 |
| cg22088495 | 2 | 85804653 | VAMP8 | 5'UTR | 4.6E-02 | 0.007 |
| cg05894754 | 2 | 210675440 | UNC80 | Body | 2.4E-02 | 0.114 |
| cg06750177 | 3 | 172341525 |  | IGR | 1.5E-02 | 0.069 |
| cg17104149 | 4 | 26492421 | CCKAR | TSS1500 | 3.8E-02 | 0.046 |
| cg26916851 | 4 | 141117172 |  | IGR | 1.3E-02 | 0.060 |
| cg20506240 | 4 | 142023809 | RNF150 | Body | 4.9E-02 | 0.087 |
| cg12140212 | 4 | 188477987 | LOC100506272 | Body | 4.1E-02 | 0.062 |
| cg08295865 | 5 | 1790944 |  | IGR | 4.1E-02 | 0.042 |
| cg04225172 | 5 | 31767552 |  | IGR | 1.6E-02 | 0.049 |
| cg11499025 | 5 | 140739655 | PCDHGA4 | Body | 1.0E-02 | -0.060 |
| cg20815819 | 6 | 2396932 | GMDS-AS1 | Body | 4.4E-02 | -0.009 |
| cg20198768 | 6 | 29635579 | MOG | 3'UTR | 5.0E-02 | 0.090 |
| cg14801238 | 6 | 31275664 |  | IGR | 4.4E-02 | -0.078 |
| cg26938522 | 6 | 158497675 | SYNJ2 | ExonBnd | 4.6E-02 | 0.010 |
| cg00271210 | 6 | 167070053 | RPS6KA2 | Body | 3.8E-02 | -0.085 |
| cg07361047 | 6 | 167533317 | CCR6 | 5'UTR | 4.1E-02 | 0.060 |
| cg16622899 | 7 | 1577003 | MAFK | 5'UTR | 1.6E-02 | -0.007 |
| cg06280368 | 7 | 12663282 | SCIN | Body | 3.8E-02 | 0.056 |
| cg09433113 | 7 | 28701898 | CREB5 | Body | 4.4E-02 | 0.076 |
| cg04541854 | 7 | 42147789 | GLI3 | Body | 4.1E-02 | 0.012 |
| cg11180069 | 7 | 131422872 |  | IGR | 3.6E-02 | -0.077 |
| cg20031843 | 8 | 17656574 | MTUS1 | 5'UTR | 4.4E-02 | 0.025 |
| cg17835356 | 8 | 41387960 | GINS4 | Body | 1.8E-02 | 0.047 |
| cg14645880 | 9 | 134501604 | RAPGEF1 | Body | 2.1E-02 | 0.008 |
| cg22499994 | 10 | 17071728 | CUBN | Body | 4.4E-02 | 0.008 |
| cg23477281 | 10 | 89263190 | MIR4678 | TSS1500 | 2.4E-02 | -0.008 |
| cg09144424 | 10 | 97050675 | PDLIM1 | 1stExon | 1.8E-02 | 0.007 |
| cg19209225 | 10 | 102756912 | LZTS2 | TSS200 | 1.4E-05 | 0.031 |
| cg21150100 | 10 | 103132349 | BTRC | Body | 1.5E-02 | 0.017 |
| cg22792014 | 10 | 134267908 |  | IGR | 4.1E-02 | -0.018 |
| cg12887863 | 11 | 13325247 | ARNTL | 5'UTR | 4.4E-02 | 0.014 |
| cg22992837 | 11 | 113643339 | ZW10 | Body | 4.4E-02 | 0.028 |
| cg20141578 | 12 | 12225262 | BCL2L14 | 5'UTR | 1.3E-02 | 0.133 |
| cg09005886 | 12 | 15517124 | PTPRO | Body | 3.8E-02 | 0.049 |
| cg13211181 | 12 | 25801455 | IFLTD1 | 1stExon | 1.0E-02 | 0.053 |
| cg01691592 | 12 | 25801556 | LMNTD1 | TSS200 | 4.4E-02 | 0.050 |
| cg18832388 | 12 | 73524285 |  | IGR | 5.0E-02 | -0.036 |
| cg18797859 | 12 | 96253319 | SNRPF | Body | 2.1E-02 | -0.018 |
| cg01257889 | 12 | 113230065 | RPH3A | 5'UTR | 4.8E-02 | -0.085 |
| cg19769395 | 12 | 114680340 |  | IGR | 4.9E-02 | -0.008 |
| cg14648877 | 12 | 124434035 | CCDC92 | 5'UTR | 2.4E-02 | -0.020 |
| cg13114549 | 14 | 20917403 | OSGEP | Body | 1.5E-02 | 0.014 |
| cg22432580 | 14 | 54955579 | GMFB | Body | 1.8E-02 | -0.006 |
| cg15530474 | 14 | 99696161 | BCL11B | Body | 4.4E-02 | 0.023 |
| cg15140996 | 15 | 95038079 |  | IGR | 1.6E-02 | 0.014 |
| cg07870479 | 16 | 1813987 | MAPK8IP3 | Body | 4.4E-02 | -0.013 |
| cg00251271 | 16 | 1832493 | SPSB3 | 5'UTR | 3.3E-02 | -0.007 |
| cg22477428 | 16 | 2837938 | PRSS33 | TSS1500 | 1.5E-02 | 0.042 |
| cg02910054 | 16 | 12241554 | SNX29 | Body | 1.6E-02 | 0.104 |
| cg06903478 | 17 | 76183632 | TK1 | TSS1500 | 4.4E-02 | -0.034 |
| cg22194807 | 18 | 32957683 | ZNF396 | TSS1500 | 4.4E-02 | -0.010 |
| cg23195373 | 19 | 45954891 |  | IGR | 5.0E-02 | 0.038 |
| cg01454134 | 19 | 56686707 | GALP | TSS1500 | 4.9E-02 | 0.039 |
| cg24045875 | 20 | 61425320 |  | IGR | 1.0E-02 | 0.014 |
