## Supplementary figures and images for "Specific methylation marks in promoter regions are associated to the pathogenic process of Chronic Chagas disease Cardiomyopathy by modifying transcription factor binding patterns"

### Supplementary figure 1.TIF

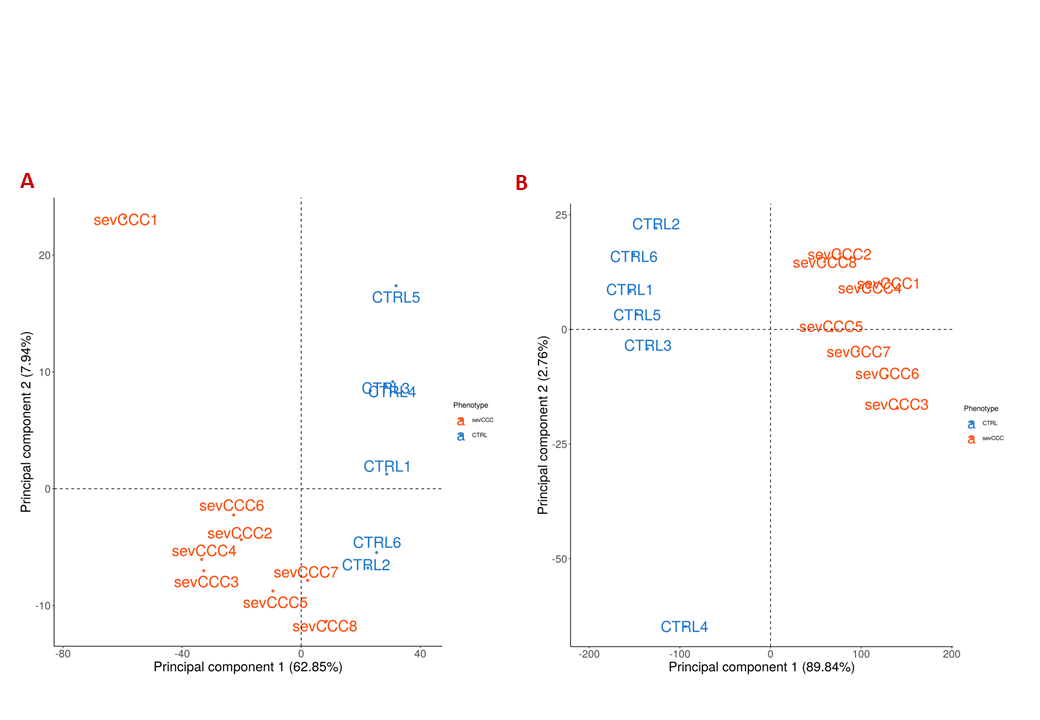

### Supplementary figure 2.TIF

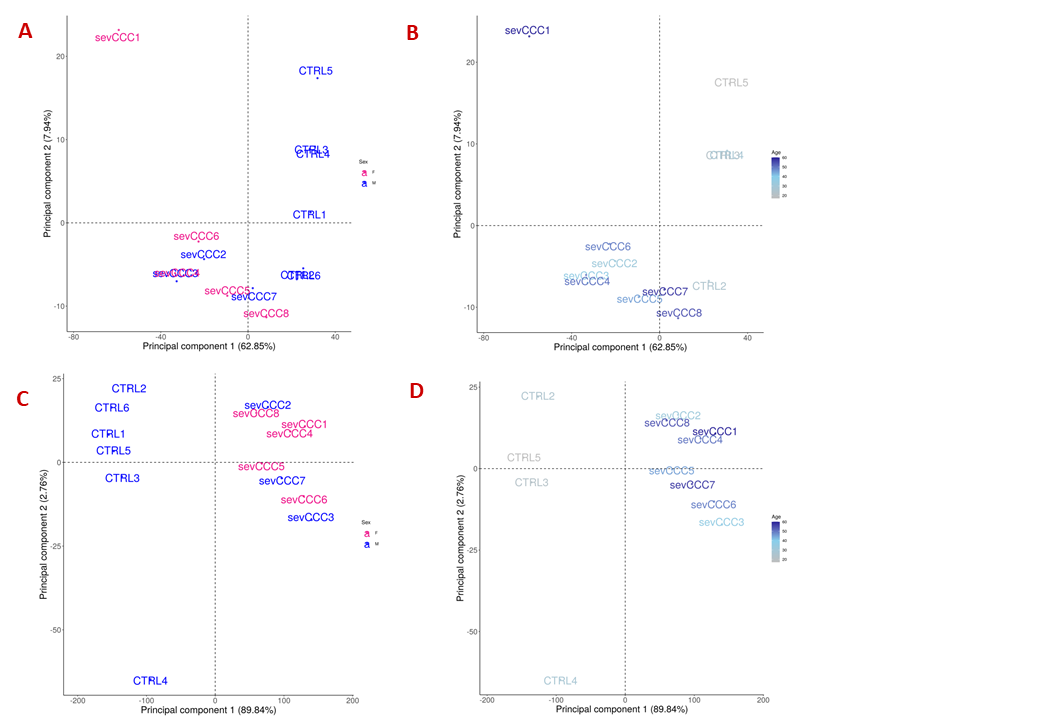

### Supplementary figure 3.TIF

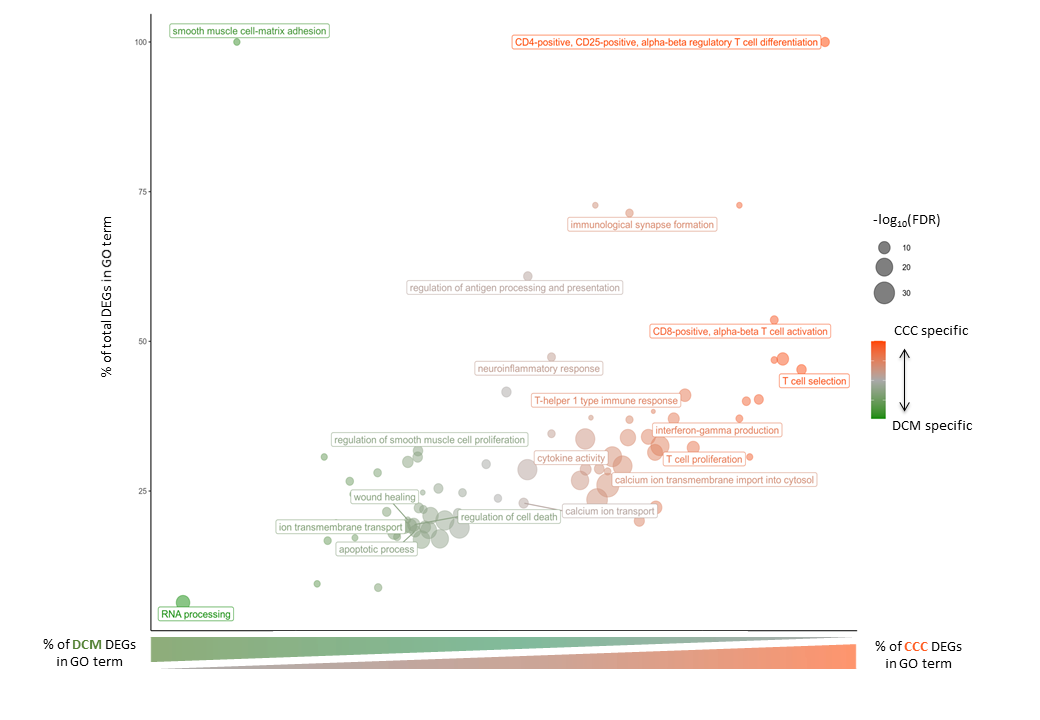

### Supplementary figure 4.TIF

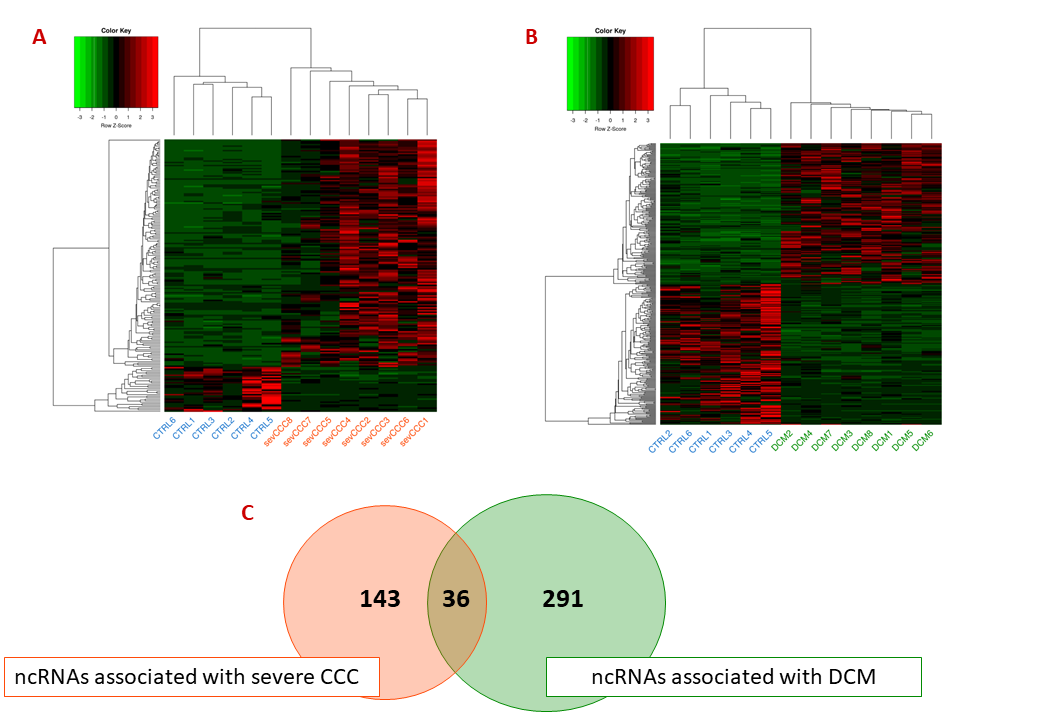

### Supplementary figure 5.TIF

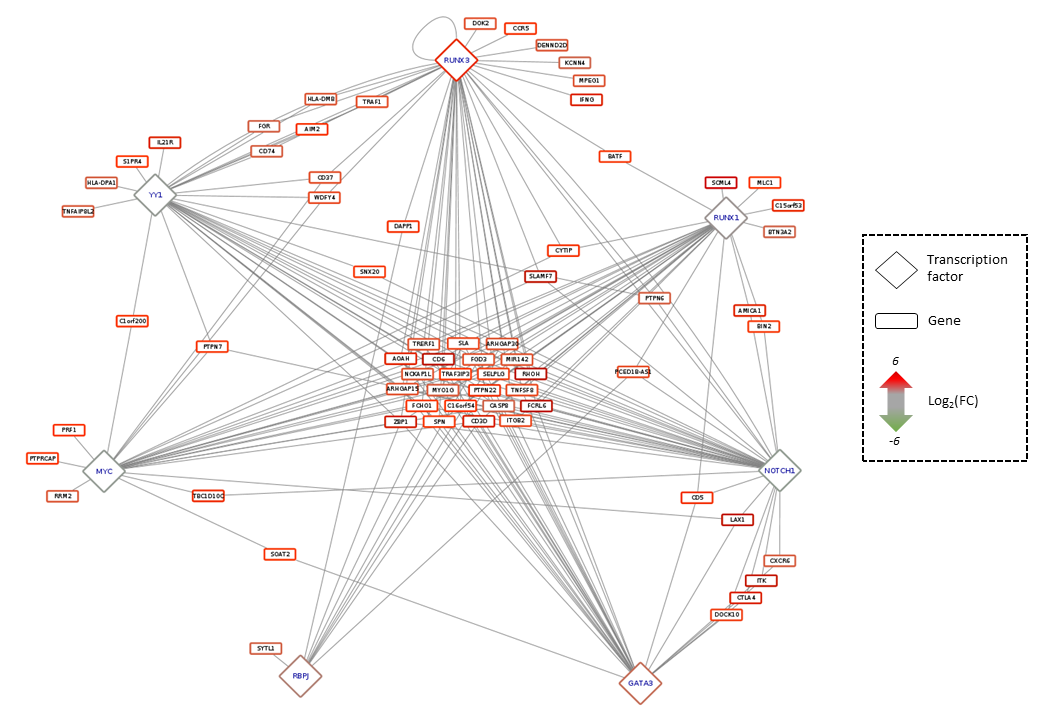

### Supplementary figure 6.TIF

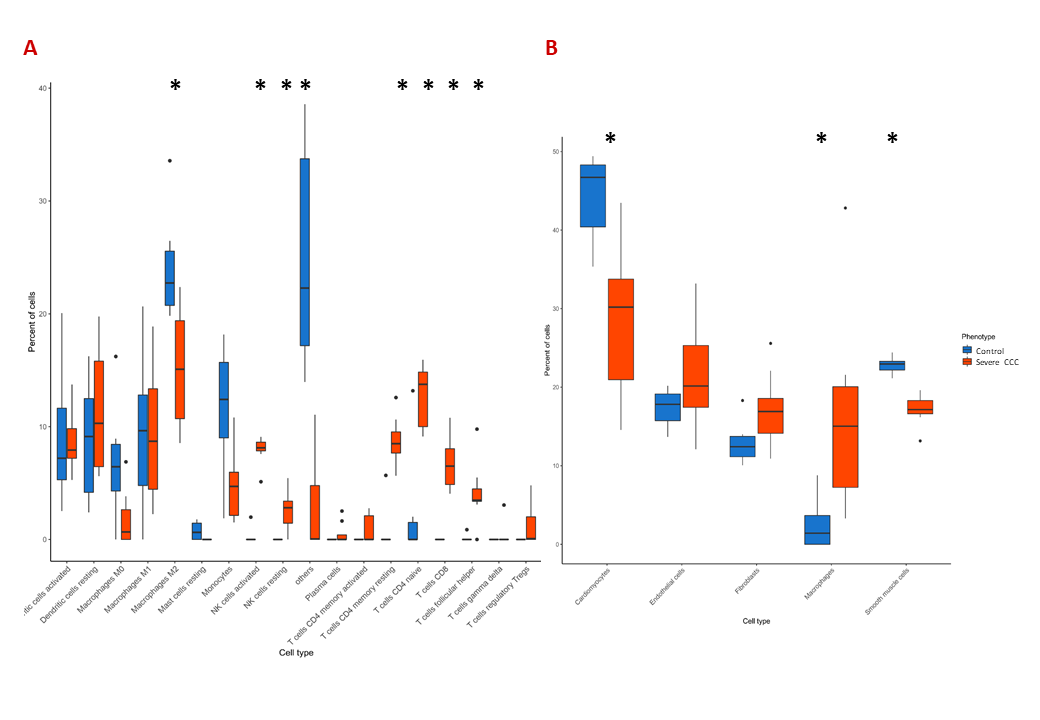

### Supplementary figure 7.TIF

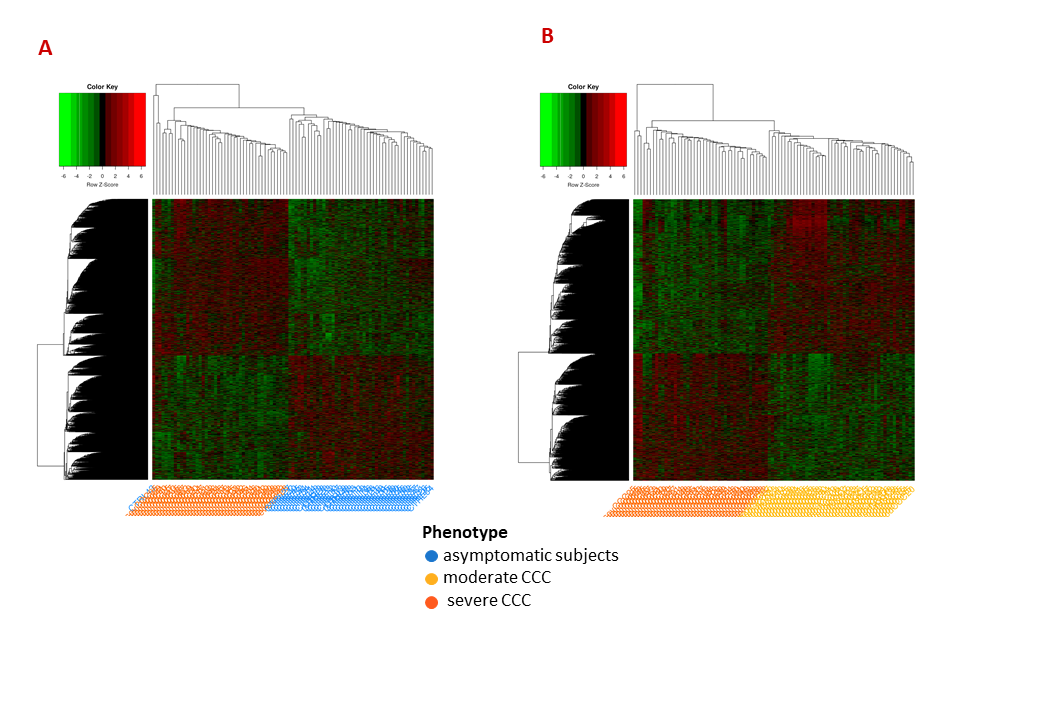
